# Sex-specific genetic loci linked to early and late onset type 2 diabetes

**DOI:** 10.1101/2022.10.27.22281587

**Authors:** Jaime Berumen, Lorena Orozco, Héctor Gallardo-Rincón, Rosa Elba Benuto, Espiridión Ramos-Martinez, Fernando Rivas, Humberto García-Ortiz, Melissa Marin-Medina, Elizabeth Barrera, Eligia Juárez-Torres, Anabel Alvarado Silva, Luis Alberto MartÍnez-Juárez, Julieta Lomelín-Gascón, Alejandra Montoya, Janinne Ortega-Montiel, Diego-Abelardo Alvarez-Hernández, Roberto Tapia-Conyer

## Abstract

**Purpose:** To investigate the effect of sex and age on the timing of a type 2 diabetes (T2D) diagnosis and the influence T2D-related genes, parental history of T2D, and obesity on T2D development.

**Methods:** In this case-control study, 1012 T2D cases and 1008 healthy subjects were selected from the Diabetes in Mexico Study database. Participants were stratified by sex and age at T2D diagnosis (early, ≤45 years; late, ≥46 years). Seventy T2D-associated SNPs were explored and the percentage contribution (R^2^) of T2D-related genes, parental history of T2D, and obesity (body mass index [BMI] and waist-hip ratio [WHR]) on T2D development was calculated using univariate and multivariate logistic regression models.

**Results:** T2D-related genes influenced T2D development most in males who were diagnosed early (R^2^ = 23.5%; females diagnosed early, R^2^ = 13.5%; males and females diagnosed late, R^2^ = 11.9% and R^2^ = 7.3%, respectively). With an early diagnosis, insulin production genes were more influential in males (76.0% of R^2^) whilst peripheral insulin resistance genes were more influential in females (52.3% of R^2^). With a late diagnosis, insulin production genes from chromosome region 11p15.5 notably influenced males while peripheral insulin resistance and inflammation genes notably influenced females. Influence of parental history was higher among those diagnosed early (males, 19.9%; females, 17.5%) versus late (males, 6.4%; females, 5,3%). Unilateral maternal T2D history was more influential than paternal T2D history. BMI influenced T2D development for all, while WHR exclusively influenced males.

**Conclusions:** The influence of T2D-related genes, maternal T2D history, and fat distribution on T2D development was greater in males than females.

## Introduction

The etiological parameters of type 2 diabetes (T2D) have not been fully elucidated; however, T2D is clearly associated with obesity, parental history, genes, ethnicity, poor eating habits, dyslipidemia, lack of physical activity, and aging, among others (1). Of these, obesity, parental history, and genes appear most important. Obesity, a modifiable factor, is clearly associated with T2D development and is considered the main risk factor; T2D risk increases linearly with increased body mass index (BMI) (2-4). However, the relationship between T2D and obesity may not be as direct as thought. For example, in countries such as China, India, and Japan, where the prevalence of T2D is high, the prevalence of obesity is relatively low. Currently, we know that approximately two-thirds of people with T2D are overweight or obese; however, only 2%–13% of obese people develop T2D. When using univariate logistic regression (ULR) models, we recently reported that the variability (R^2^) of T2D attributed to obesity would be 12.6%–14.9% (5). However, these values vary substantially in multivariate logistic regression (MLR) models that include parental history of T2D and genes as variables (5).

The importance of parental history on T2D development has been well established in both family (6-8) and association studies (9). The heritability of T2D is variable, reported to be around 25% in family studies (9-11) and 20% in association studies (6, 7, 12). In a case– control study conducted in Mexico, the R^2^ of T2D attributed to parental history of diabetes was 12.1%. There are currently over 400 genes with an identified association with T2D (13). However, based on their low individual odds ratios (OR), most genes have very little influence on the development of the disease, which can vary with different study populations. Most studies suggest that genetic involvement contributes to no more than 10% of T2D variability (13, 14). Recently, a study of 16 single nucleotide polymorphisms (SNPs) with the highest predictive power for T2D in a Latin American population reported that only nine were associated with T2D and that this contributed to only 6.5% of T2D variability (5). However, as previously reported (15), this study found that parental history and genes have a greater influence on T2D when it occurs before the age of 46 and that the influence of these factors decreases substantially when the disease is diagnosed later in life (≥46 years). Furthermore, this study also found an important difference in the association of genes, but not of parental history, with T2D between males and females (15). Here, the R^2^ value was much higher in males than females (11.2% versus 4.1%), and given that the differences in gene association between males and females have not been previously reported or have only been reported for individual genes (16, 17), the differences are likely specific to the ancestry of populations in Mexico, which include a mixture of different Amerindian and European ancestries.

In fact, in recent high-throughput studies, as part of the Slim Initiative in Genomic Medicine for the Americas (SIGMA), polymorphisms strongly associated with T2D have been discovered in Mexican and Latin populations that are either non-existent or very rare in European populations. These polymorphisms are located in *SLC16A11* (18), *INS-IGF2* (19), and *HNF1A* (20). Here, the difference between the R^2^ values of males and females may be partially explained by the difference in the effect size of these genes between sexes. We note that polymorphisms in well-established genes associated with T2D in multiple populations, such as *KCNQ1* and *TCF7L2*, are also important contributors to this difference (5). Because of the small number of genes analyzed in a previous study, we were unable to investigate which biological processes associated with T2D (e.g., insulin production, peripheral insulin resistance, inflammation) were most relevant for the genetic differences between men and women and between early- and late-onset T2D. Given the difference in the degree of association of genes between the two sexes and between the late and early presentation of T2D, it is likely that the biological processes associated with T2D in each group are markedly different.

In this work, a case–control study (n = 2020) was performed, including 69 SNPs associated with T2D located in genes involved in different biological processes. The aim was to solve the following research questions: 1) to confirm whether the influence of genes on T2D is different between men and women, 2) to investigate which biological processes have different weights of contribution in T2D between sexes and age at T2D presentation, and 3) to understand how the contribution of each biological process changes in each group when obesity and parental history of diabetes are included in the MLR models. To identify the factors associated with T2D in each group, univariate logistic regression (ULR) models were used. Then, MLR models were used to calculate the percentage contribution (R^2^) of each factor to the variability of T2D. In the latter, the genes (grouped by biological processes) and the remaining factors were entered in successive blocks to calculate the contribution of each factor (R^2^) in the global model. The variation in the order of entry of each factor allowed us to identify how obesity and parental history affect the importance of gene biological processes linked to insulin production and resistance in the variability of T2D between sexes and age of disease onset.

## Material and Methods

### Sample selection and study design

Study participants were part of the Diabetes in Mexico Study (DMS) (21). The DMS study design was previously described as part of the SIGMA Type 2 Diabetes Consortium. Briefly, participants were recruited from two tertiary-level hospitals in Mexico City (Centro Médico Nacional siglo XXI del Seguro Social Mexicano [IMSS], Centro Médico Nacional 20 de Noviembre del Seguro Social para los Trabajadores del Estado [ISSSTE]). T2D was diagnosed according to the criteria of the American Diabetes Association (22). A total of 1012 cases (unrelated individuals, >20 years old, with a previous diagnosis of T2D or fasting glucose levels >125 mg/dL) and 1008 controls (healthy subjects, >50 years old, fasting glucose levels <100 mg/dL) were selected from the DMS database, which consisted of 1500 controls and 1500 cases of the Latin American mestizo population. Participants and same-sex, same-hospital controls were recruited between November 2009 and August 2013. Clinical information including weight, waist and hip circumference, and parental history of T2D was collected during the initial interview. For fasting glucose measurements and DNA extraction, 10 mL of intravenous blood was collected. The association of the 69 selected SNPs, parental history of T2D, BMI, and WHR with T2D was investigated using ULR and MLR models.

This study was approved by the Ethics and Research Committees of the National Institute of Genomic Medicine and the Federal Commission for the Protection against Health Risks (COFEPRIS; approval number CAS/OR/CMN/113300410D0027-0577/2012). The study protocols were carried out in accordance with the ethical principles outlined in the 1964 Declaration of Helsinki. Written informed consent was obtained from all participants prior to their inclusion in the study. This study was not registered in a clinical trial registry.

### SNP selection and genotyping

Sixty-nine SNPs were selected; nine associated with T2D in a previous study (5) and 60 from different GWAS databases and published articles (**Table S1**). SNPs with a minor allele frequency (MAF) ≥10% and OR ≥ 1.2 or ≤ 0.83 in the Latin American mestizo population and those localized in genes with a role in cellular processes involved in T2D development were prioritized. All DNA samples were genotyped for the 69 SNPs listed in **Table S1**. Assays were designed using Applied Biosystems TaqMan SNP assay design technology (Foster City, CA, USA). Genotyping was performed by the allelic discrimination assay-by-design TaqMan® method on OpenArray® plates. The plates were analyzed on the QuantStudio™ 12 K Flex Real-Time PCR System (ThermoFisher Scientific, Waltham, MA, USA). The genotypes were analyzed by Genotyper™ Software v1.3 (ThermoFisher Scientific, Waltham, MA, USA).

A total of 23 SNPs that had a difference of distribution between cases and controls of *P* < 0.2 in the ULR models were ultimately included in the MLR models. The list of SNPs and gene symbols are presented in **Table 1**, and the names of genes and their function in **Table S2**. The SNPs were classified into four groups according to the function and location of the gene in which they occur: Group 1, insulin production located in chromosome region 11p15.5 (n = 5); Group 2, insulin production located in other regions (n = 7); Group 3, peripheral insulin resistance (n = 6); and Group 4, inflammation and other functions (n = 5) (**Table 1, Table S2**).

**Table 1.**
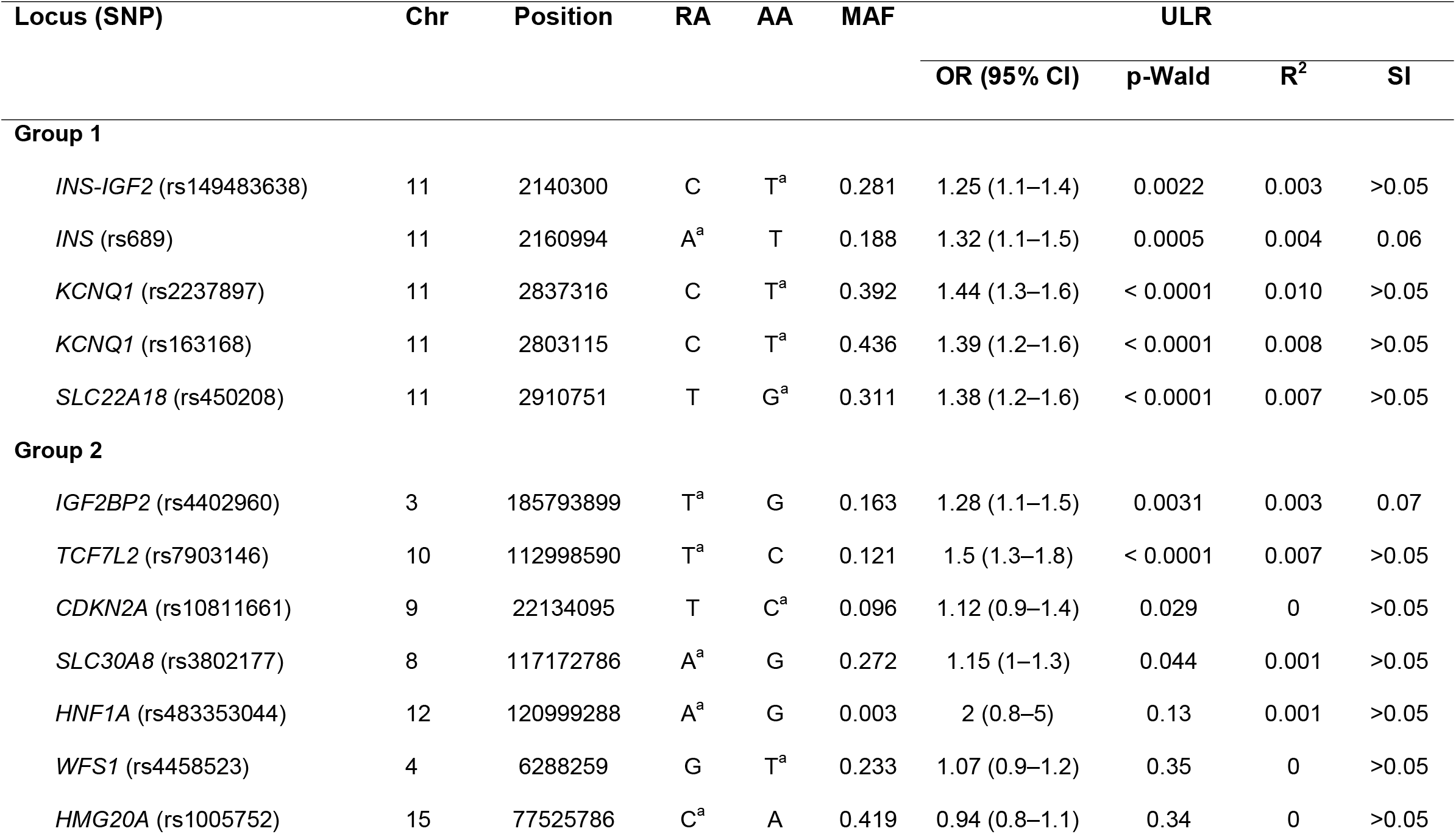
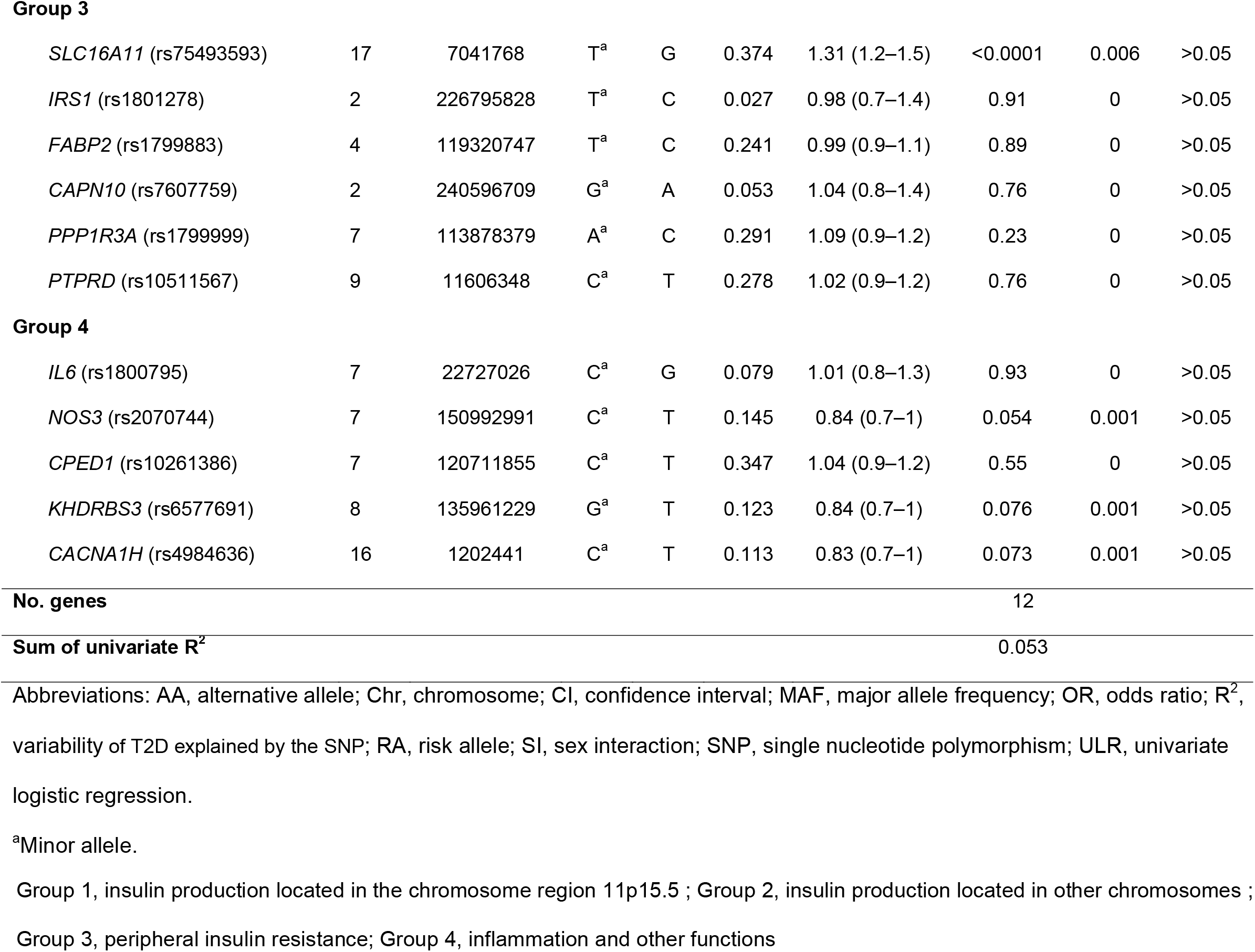
Association of 23 SNPs with type 2 diabetes among the whole sample (N = 4040 chromosomes)

### Statistical analyses

When determining the sample size, we considered MLR models to have good performance when there was a baseline of 50 cases (as in the ULR models) and 10–15 additional cases for each variable introduced in the model (23). Given that we planned to introduce 15–20 variables in the MLR models, we calculated a minimum number of 200–350 cases in the comparison groups.

Participant age, age at T2D diagnosis, BMI, and WHR were expressed as mean ± standard deviation (SD). Distribution of the frequency of genotypes according to the Hardy– Weinberg law based on the allelic frequency and the formula: (*a* + *b*)^2^ = *a*^2^ + 2ab + *b*^2^, where *a* and *b* are the allelic frequencies in the control group, was investigated. Differences in the distribution of genotypes between the observed and expected results were calculated using the chi-square test. The BMI was adjusted (BMIadj) for participants who had been diagnosed with T2D for ≥3 years using data from patients who had been diagnosed with T2D for ≤2 years (5). In the ULR and MLR models, case (diagnosis of T2D) or control was considered a dependent variable; values of the alleles and genotypes of the SNPs, parental history, BMI, and WHR were considered explanatory variables. Interactions between each SNP and both sex and age of T2D presentation were assessed in the ULR models. All variables (BMIadj, WHR, parental history of T2D, and SNPs) were analyzed with stratification by sex (male and female) and age at T2D diagnosis (≤45 years and ≥46 years). The risk conferred by each factor (explanatory variables) was calculated by comparing cases and controls using ULR models. Association was expressed as an OR with 95% confidence intervals (CIs); the contribution to the variability of T2D was expressed as Nagelkerke’s R^2^, which represents the percentage of T2D variability that can be explained by a named factor (24). Confounders were identified using a theoretical strategy based on a backstep, stepwise MLR model and the change-in-estimate criterion. Variables with *P* < 0.20 in the ULR analysis were considered for entry in the MLR model. Confounders were defined as those variables for which the percentage difference between the values of the regression β between the adjusted and non-adjusted variables in the stepwise MLR model was larger than 10% (*P* > 0.1). Therefore, the total variability and the contribution of each factor on T2D was calculated using this MLR model. These factors were included successively in the model in different blocks, and the contribution of each factor to the model was assessed by the increase of R^2^ and the decrease in the −2 log likelihood ratio value from one block to the next; the Omnibus test was used to determine whether the differences between the successive blocks were statistically significant. A post-hoc power analysis was performed for each logistic regression model using the software G * Power 3.1.9.2, considering the sample size, the OR, the probability of the event in the control group, and an α = 0.05 (25). In addition, for MLR models, the value of the total R^2^ obtained at the end of the model was introduced for power calculation.

All statistical tests were two-sided, and differences were considered significant when *P* < 0.05. Statistical analyses were conducted using SPSS version 20 software (IBM Corp., Armonk, NY, USA).

## Results

### Participant characteristics

The 2020 participants (cases, 1012; controls, 1008) were included in the SNP and BMI analyses; given missing data for some participants, 1690 were included in the parental history analysis of T2D and 1335 in the WHR analysis. WHR was not included in some MLR models; a total of 1690 participants were included in models where parental history was introduced. The demographic characteristics of patients with T2D and non-diabetic controls are presented in **Table S3**.

Among the participants, 53.9% were female and 46.1% were male. The mean ± SD age of non-diabetic controls (59.1 ± 11.3 years) was higher than that of cases (55.5 ± 11.7 years) at the time of enrollment. The mean ± SD age at T2D diagnosis was 45.8 ± 10.7 years, and the number of years with T2D varied widely (range, 0–46 years; mean ± SD, 9.5 ± 8.7 years). At the time of study recruitment, most patients (89.5%) were taking medication for their T2D, and few were smokers (cases, 20.2%; controls, 19.8%).

### Identification of genes associated with T2D using ULR models

The allelic frequencies of only 23 (**Table 1**) of the 69 SNPs explored (**Table S1**) were significantly different (*P* < 0.1) between cases and controls in all participants (**Table S4**), stratified by age at T2D diagnosis (≤45 years and ≥46 years) (**Table S4**), sex (male and female) (**Table S5**), or both (**Table S6**).

In the allelic ULR analysis, 12 of 23 SNPs showed a significant association with T2D for the entire study population (**Table 1**). The R^2^ sum of these 12 SNPs only explains 5.3% of the variability of T2D etiology.

When analyzed according to age of T2D diagnosis (≤45 years versus ≥46 years), 8 of the 12 SNPs associated with T2D in the whole sample were associated with T2D in both age groups (**Table S7**). SNPs in *SLC30A8* (rs3802177), *NOS3* (rs2070744), and *KHDRBS3* (rs6577691) were associated only with early T2D diagnosis and SNPs in *CACNA1H* (rs4984636) were only associated with late diagnosis. Two additional SNPs, not associated with T2D in the analysis of the entire study population, were also identified: *WFS1* (rs4458523) (associated with early T2D diagnosis) and *HMG20A* (rs1005752) (associated with late T2D diagnosis) (**Table S7**). No major differences were observed in the association of gene groups between early or late T2D diagnosis. However, all SNPs except *INS-IGF2* (rs149483638) had a greater association with early than late T2D diagnosis. Compared with the overall analysis, the R^2^ sum increased in SNPs associated with early T2D diagnosis (R^2^ sum = 7.37%) and decreased in those associated with late T2D diagnosis (R^2^ sum = 3.32%).

There was a significant interaction (*P* < 0.1) between sex and some SNPs, both in the entire study population (n = 8) and when stratified by early (n = 9) and late (n = 6) T2D diagnosis (**Table 1, Table S7**). **Table S5** shows allelic frequency and **Table S7** shows the ULR results stratified by sex. Twelve and eight of 23 SNPs were associated with T2D exclusively in males and females, respectively; five were associated with both sexes. A large difference was observed in the association of different groups of genes between males and females. For example, all five SNPs on the chromosome region 11p15.5 were associated with T2D in males but only three SNPs were in females. The two SNPs that were not associated with T2D in females were located at the upstream end of the region where *INS* and *IGF2* are located. Those shared with males, however, were located in the downstream region (*KCNQ1* and *SLC22A18*) (**Figure 1**). The two SNPs located in *KCNQ1* had a stronger association with T2D in males than females (OR >1.6 versus OR = 1.2; *P* < 0.0001 versus *P* < 0.05), while the SNP in *SLC22A18* had a slightly higher association in females versus males (OR >1.43 versus OR = 1.32; *P* < 0.0002 versus *P* < 0.007). None of the seven SNPs in genes involved with insulin production located in other genomic regions (Group 2) were associated with T2D in females; however, three SNPs (*IGF2BP2* [rs4402960], *TCF7L2* [rs7903146], *SLC30A8* [rs3802177]) were associated in males. Peripheral resistance appears more important than insulin production for T2D development in females. The SNP in *SLC16A11* had a greater effect in females than males (OR >1.37 versus OR = 1.24; *P* < 0.0004 versus *P* < 0.021) and the SNP in *PPP1R3A* was only associated with T2D in females.

**Figure 1.**
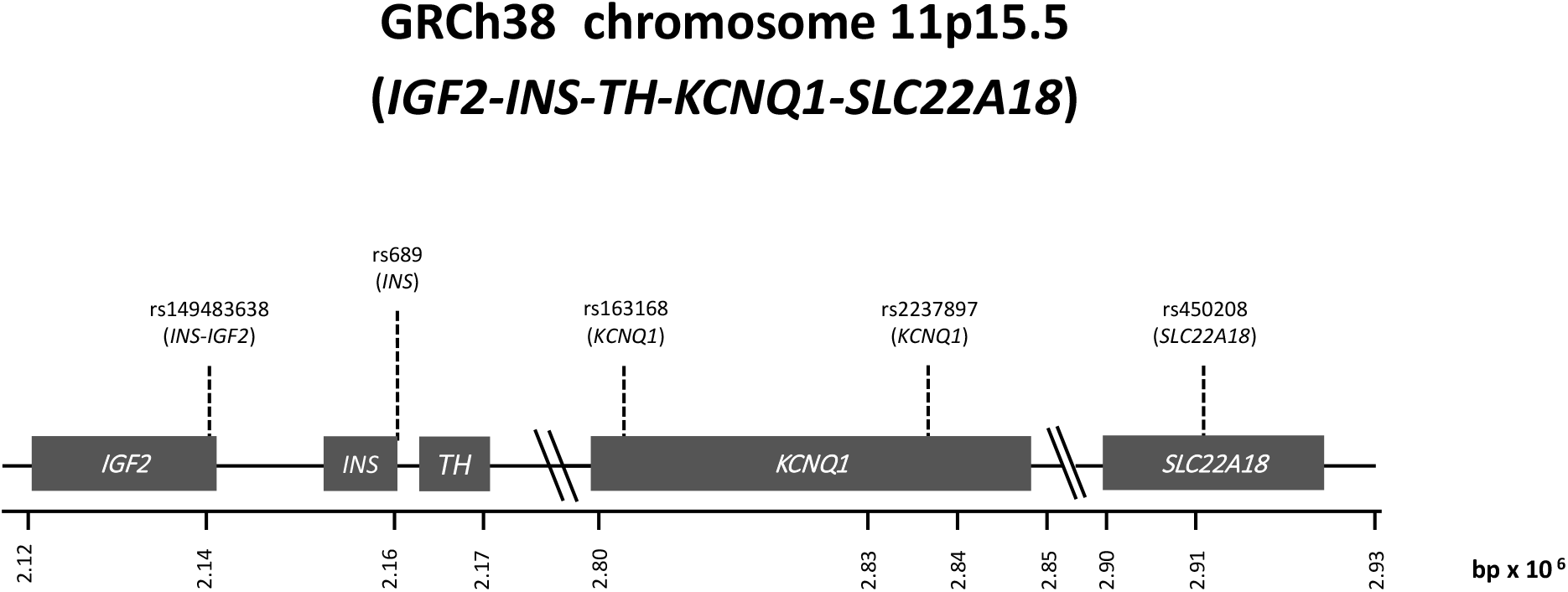
Graphic representation of the chromosome 11p15.5 region.

Overall, ORs were higher and *P* values lower in genes associated with T2D in males versus females. In females, the highest ORs were reported for SNPs in *SLC16A11* (OR, 1.37) and *SLC22A18* (OR, 1.43), while there were five SNPs with an OR > 1.5 (located in *IGF2BP2, INS, KCNQ1*, and *TCF7L2*) for males. The R^2^ sum was 3.5% for females and 10.7% for males. When the analysis was stratified by sex and age of T2D detection some differences in the associations of groups of genes were seen between the two groups of males and females (**Table 2** and see below in the MLR analysis).

**Table 2.**
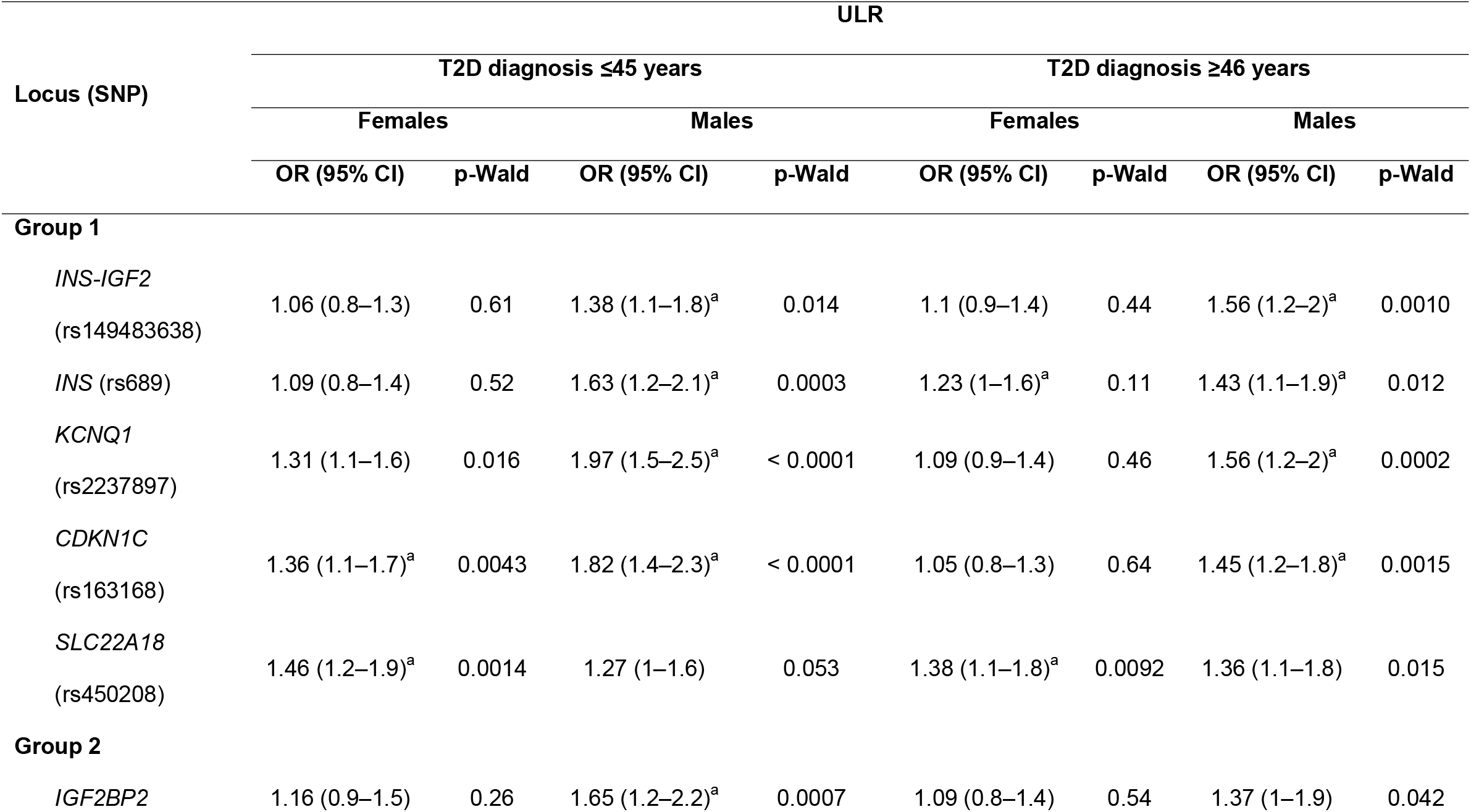
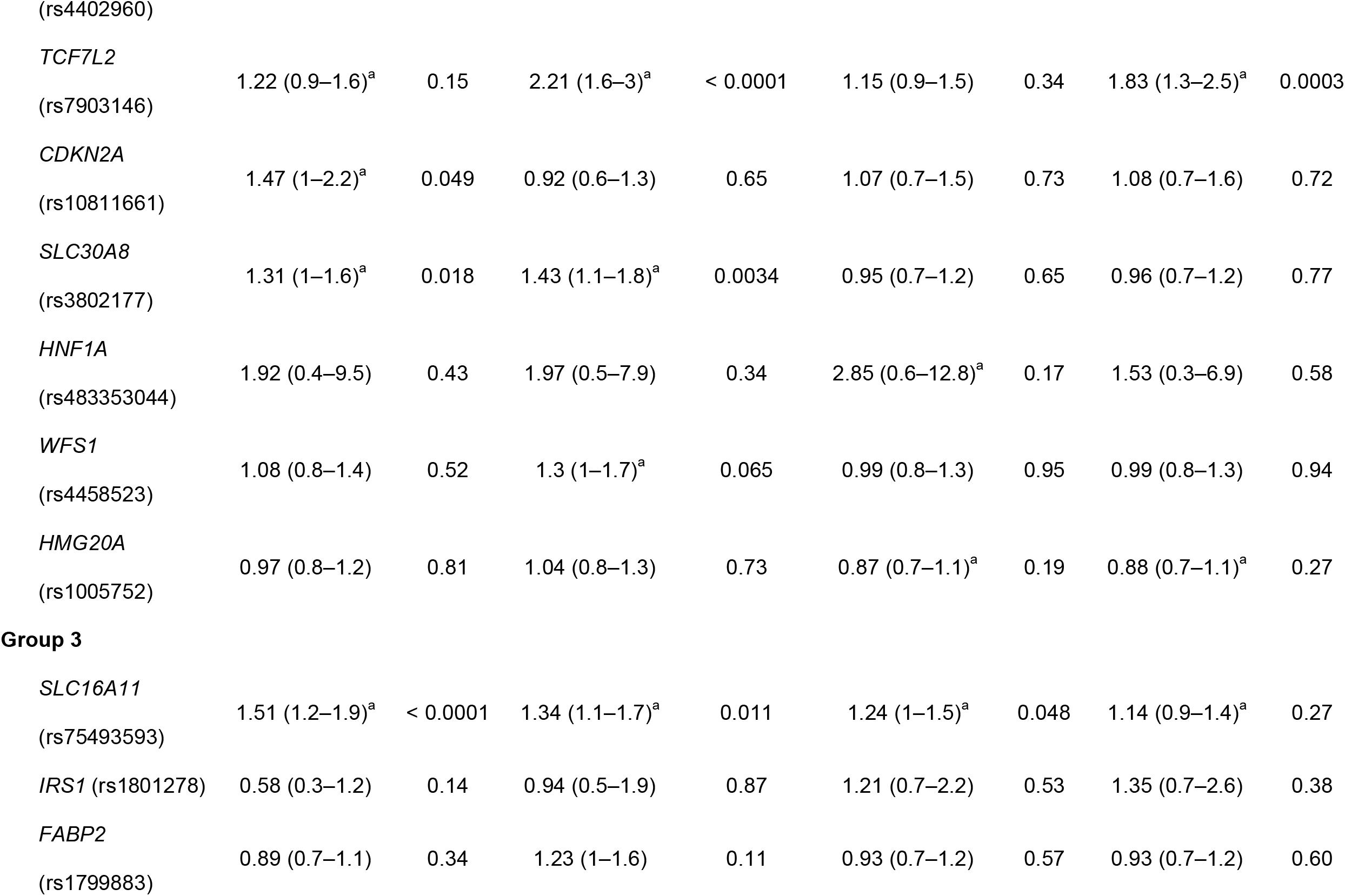
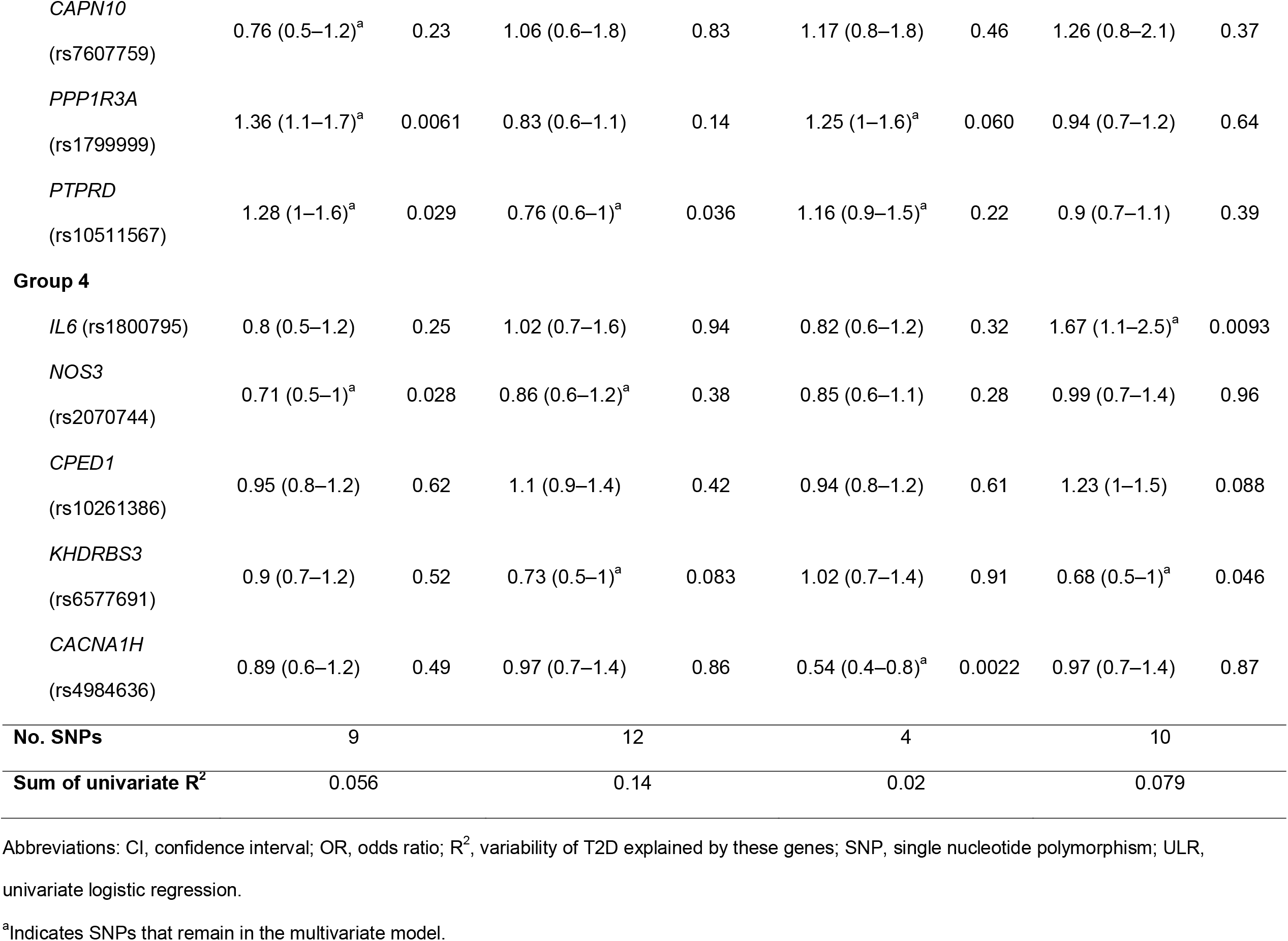
Allele association of 23 SNPs with type 2 diabetes stratified by age of type 2 diabetes diagnosis and sex among the total population (N = 4040 chromosomes).

The observed genotypic frequency was distributed according to the Hardy–Weinberg law in all 23 SNPs studied (**Table S8**). Genotypic frequency is shown in **Table S9** (the total sample stratified by age at T2D diagnosis), **Table S10** (stratified by sex), and **Table S11** (stratified by age at T2D diagnosis and sex). Overall, SNPs associated with T2D in the allelic ULR analysis also showed a significant association in the ULR genotype analysis (**Table S12**). However, in all ULR genotypic models, R^2^ sum values were much higher than those observed in the univariate allelic models, clearly indicating that genotypes rather than alleles should be used in MLR models.

### Degree of participation of each group of genes in the variability of T2D using MLR models

The MLR analyses were performed with the SNP groups (Group 1, Group 2, Group 3, and Group 4) introduced in successive blocks. Most SNPs identified by ULR, except the SNP rs450208 (SLC22A18) in males and SNP rs2237897 (KCNQ1) in females, which could be in partial linkage disequilibrium with some of the other three SNPs located in chromosome region 11p15.5 (**Table S13**), remained in the MLR models (**Table S14**).

The influence of genes on T2D was greatest in males diagnosed early (23.5%), followed by females diagnosed early (13.5%), males diagnosed late (11.9%), and females diagnosed late (7.3%). Genes on the chromosome region 11p15.5 (40.3% of the R^2^; **Figure 2A**) and others involved in insulin production (35.7% of the R^2^; **Figure 2A**) had the greatest effect in males diagnosed early, which contrasts with the major influence of genes involved in peripheral insulin resistance in females diagnosed early (52.3% of the R^2^; **Figure 2A**). Also notable, males diagnosed late had a predominance of associated SNPs in genes of the chromosome 11p15.5 region, which contrasts with the lower R^2^ values of the same SNPs in females diagnosed late. Although an increase in the influence of genes involved with inflammation was observed in males and females diagnosed late, it accounted for nearly one quarter of the genes’ influence in females.

**Figure 2.**
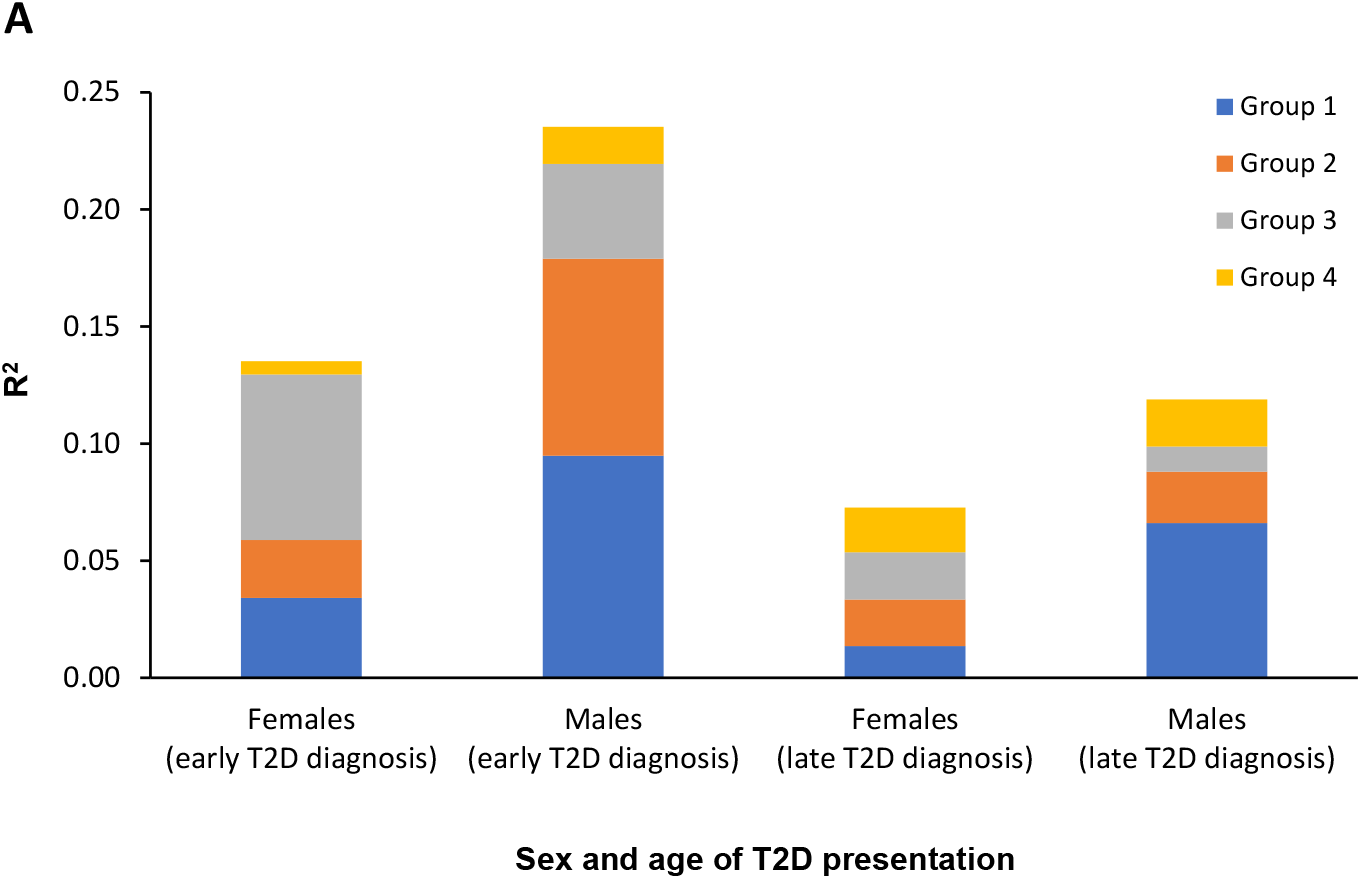
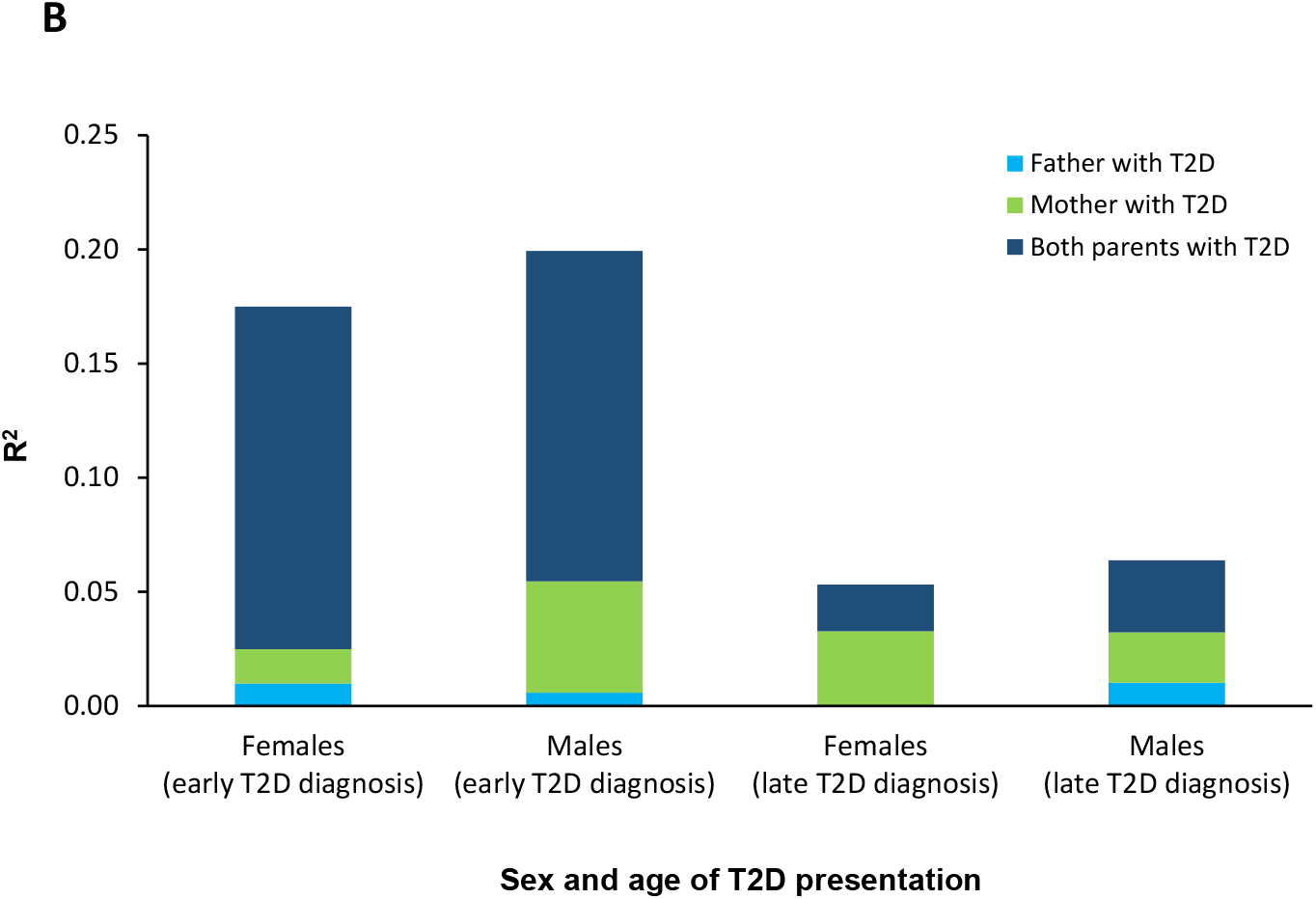
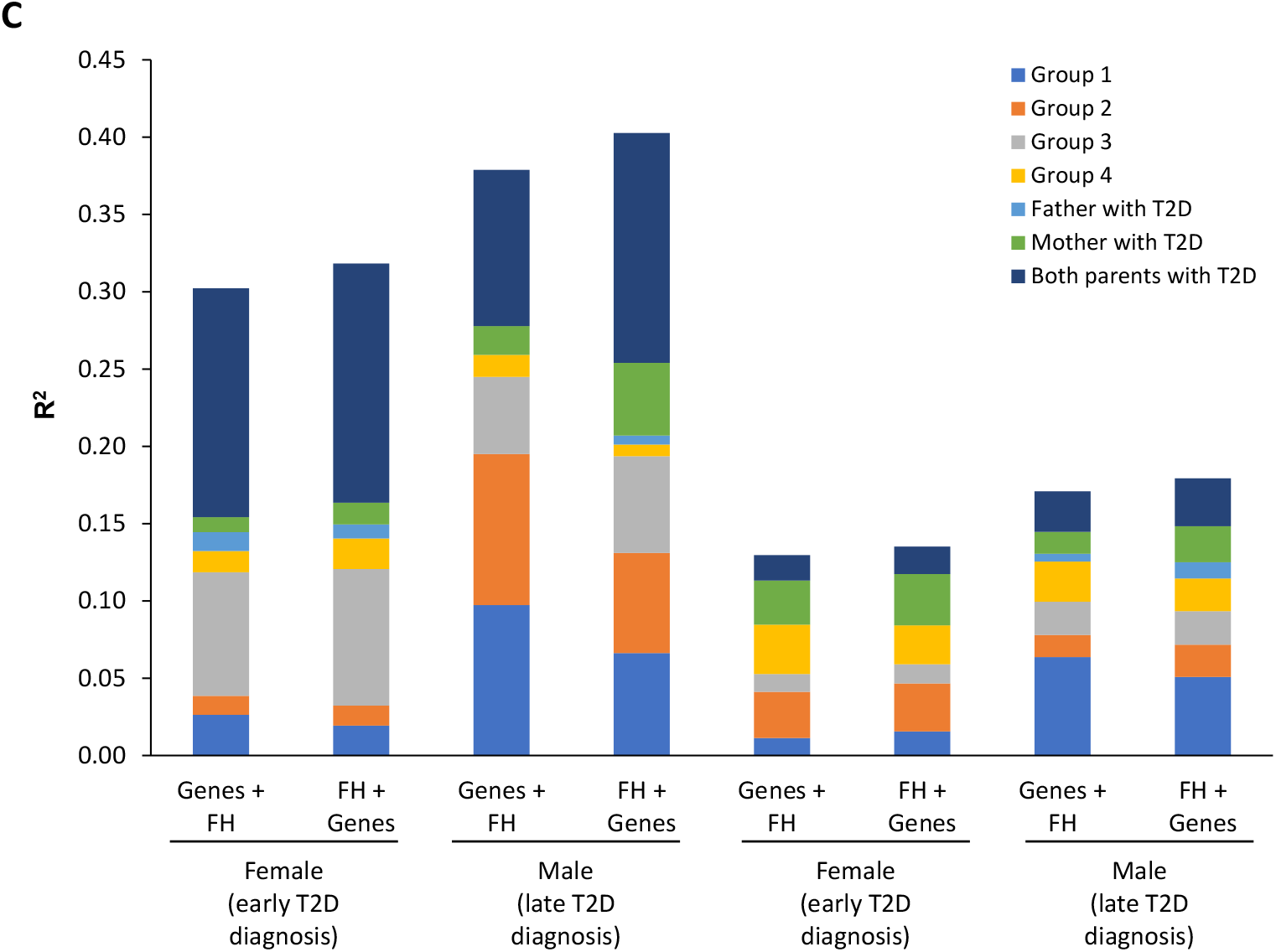
Influence (R^2^) of genes and parental history of T2D on T2D development. MLR analysis stratified by age at T2D diagnosis and sex was conducted to demonstrate (A) the effect of groups of genes, (B) parental history, and (C) both factors on T2D development (early diagnosis was ≤45 years whereas late diagnosis was ≥46 years). Gene groups and parental history as factors were introduced into the models as successive blocks. The gene groups were as follows: Group 1, five SNPs located in genes on chromosome region 11p15.5 that are in involved in insulin production (*INS, IGF2*, and *KCNQ1*); Group 2, seven SNPs located in genes outside of chromosome 11 and involved in insulin production; Group 3, six SNPs located in genes involved in peripheral insulin resistance; Group 4, five SNPs located in genes involved in inflammation and other functions. The influence of parental history of T2D was investigated according to having a mother with T2D, a father with T2D, or both parents with T2D. In panel C, the effects of genes and parental history were investigated with genes as the first block and parental history as the second block and vice versa. Abbreviations: MLR, multivariate logistic regression; SNP, single nucleotide polymorphism; T2D, type 2 diabetes.

### Influence of parental history of T2D

Parental history had the strongest influence in those cases diagnosed with T2D at ≤45 years of age (males, 19.9%; females, 17.5%) and the least influence in those cases diagnosed with T2D later in life (≤6.4%) (**Figure 2B**). History of T2D in both parents had the most influence in both males and females diagnosed early. In contrast, having only a father with T2D had little or no influence on T2D development whereas having a mother with T2D influenced T2D diagnosis, especially among males diagnosed early (25%) and females diagnosed late (50%). The risk of developing T2D early was approximately 10 times higher when both parents and three times higher when one parent had a history of T2D versus neither. In contrast, a history of T2D in both parents conferred a lower risk of T2D in those diagnosed late compared with an early diagnosis (males, 3.6 times higher; females, 2.7 times higher versus neither parent with T2D) (data not shown).

In the four groups, the total value of R^2^ was higher when parental history was the first block and genes was the second block, particularly for males diagnosed early (R^2^, 0.403) (**Figure 2C**). In this group, gene effects decreased from 0.251 to 0.201 (**Figure 2C**) as compared with genes in the first block. In contrast, the effect of parental history decreased when analyzing in the opposite order (R^2^ decreased from 0.202 to 0.119), suggesting a common effect on 5%–8% of T2D variance linked to genes and parental history of T2D, and that much of the effect of parental history is not directly due to the assessed genetic polymorphisms. Interestingly, when the effect of genes is reduced, there is not a uniform reduction for the four groups of genes. Rather, the influence (R^2^) of genes related to insulin production is decreased (chromosome region 11p15.5 [Group 1], 32%; other genomic regions [Group 2], 34%) and in genes related to peripheral insulin resistance (Group 3), the influence is increased by 25%. In the reverse model (first block, genes; second block, parental history) the decreased influence of parental history was not uniform for a history in the father (decreased from a small effect to no effect), mother (60% decrease), or both (32% decrease) in males diagnosed early (**Figure 2C**). These data may suggest a relationship between SNPs on genes in the chromosome region 11p15.5 and a maternal history of T2D, especially given that genes in this region have genetic imprinting on the maternal side (26). When stratified by the type of parental inheritance, the distribution of risk alleles compared with alternative alleles for several 11p15.5 genes and *TCF7L2* only differs between cases and controls with unilateral maternal inheritance in males diagnosed early (**Figure 3**).

**Figure 3.**
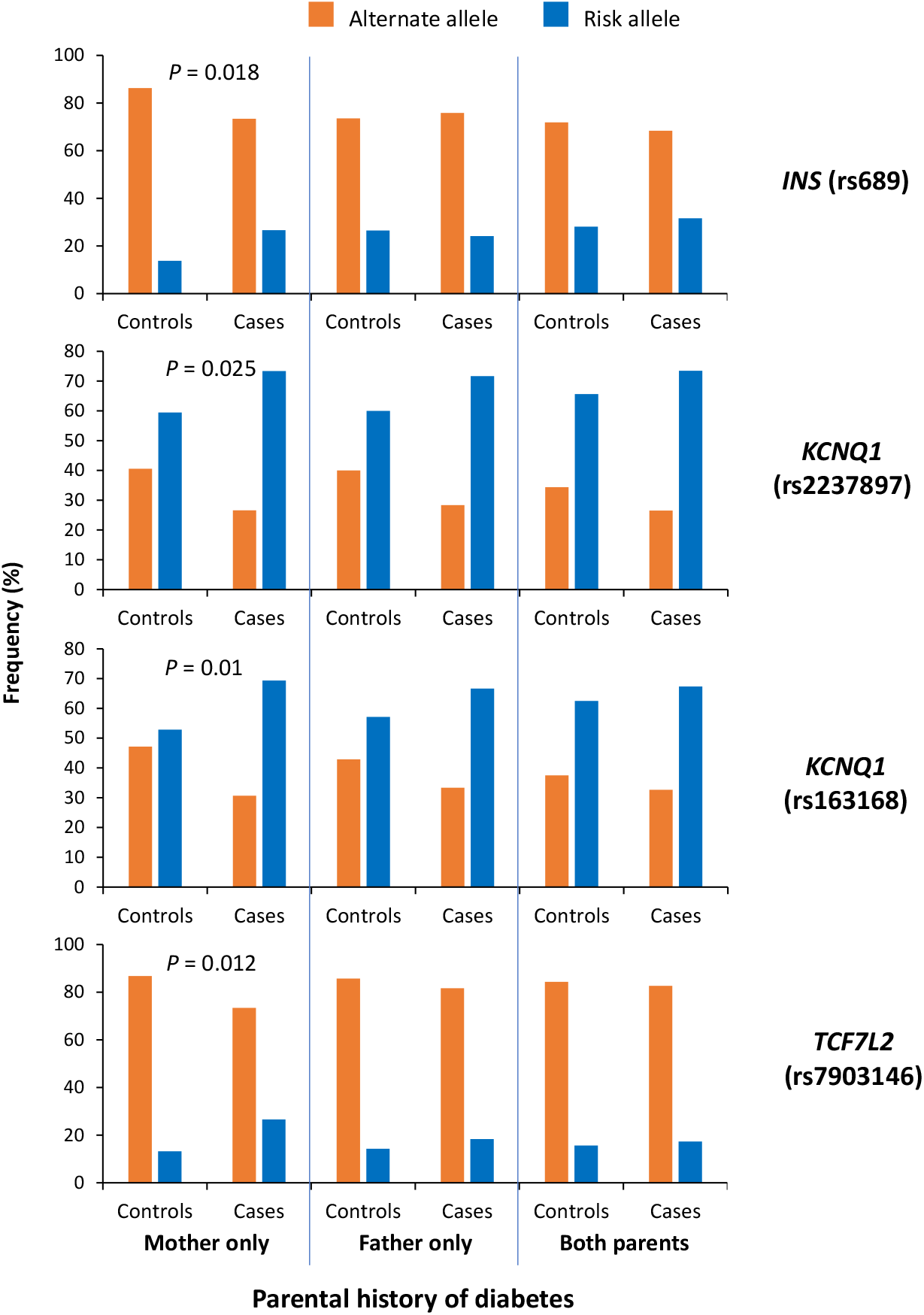
Analysis of allelic frequency stratified by type of parental history of T2D in males diagnosed early. A comparison of frequencies of risk and alternative alleles for SNPs in genes located in the chromosome region 11p15.5 (*INS* and *KCNQ1*) and *TCF7L2* between cases and controls stratified by the type of parental history of T2D (mother only, father only, both parents). *P* values were calculated using a chi-square test. Abbreviations: SNP, single nuclear polymorphism; T2D, type 2 diabetes.

### Influence of obesity and fat distribution

ULR revealed that BMI was important in all four groups of T2D (males diagnosed early, R^2^ = 0.198; males diagnosed late, R^2^ = 0.147; females diagnosed early, R^2^ = 0.132; females diagnosed late, R^2^ = 0.106) (**Figure 4A**). WHR was most important for both male groups (**Figure 4A**). BMI results were similar in MLR models when BMI was the first block (**Figure 5A** and **Figure 5B**). However, if BMI was the third block (following parental history and genetic influence), the effect decreases substantially (males diagnosed early, 54.6%; males diagnosed late, 34.3%; women diagnosed early, 40.7%; females diagnosed late, 19.9%) (**Figure 4B**).

**Figure 4.**
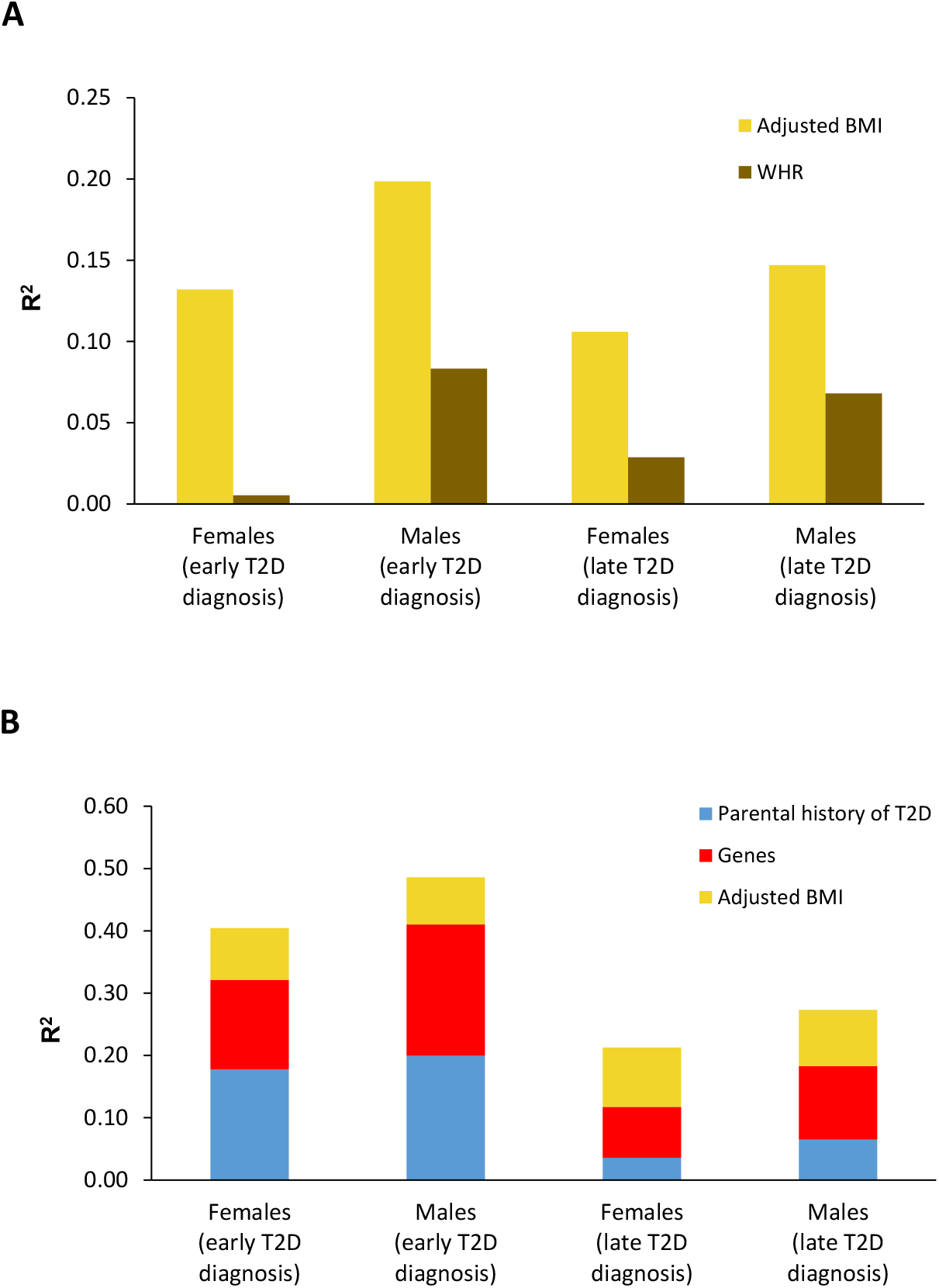
Influence (R^2^) of genes, parental history of T2D, BMI, and WHR on T2D development. (A) MLR analysis was conducted to determine the involvement of BMI and WHR in the four models of T2D (stratified by age of T2D diagnosis and sex). (B) MLR analysis of the participation of parental history, genes, and adjusted BMI in the four models. Abbreviations: BMI, body mass index; MLR, multivariate logistic regression; T2D, type 2 diabetes; WHR, waist-hip ratio.

**Figure 5.**
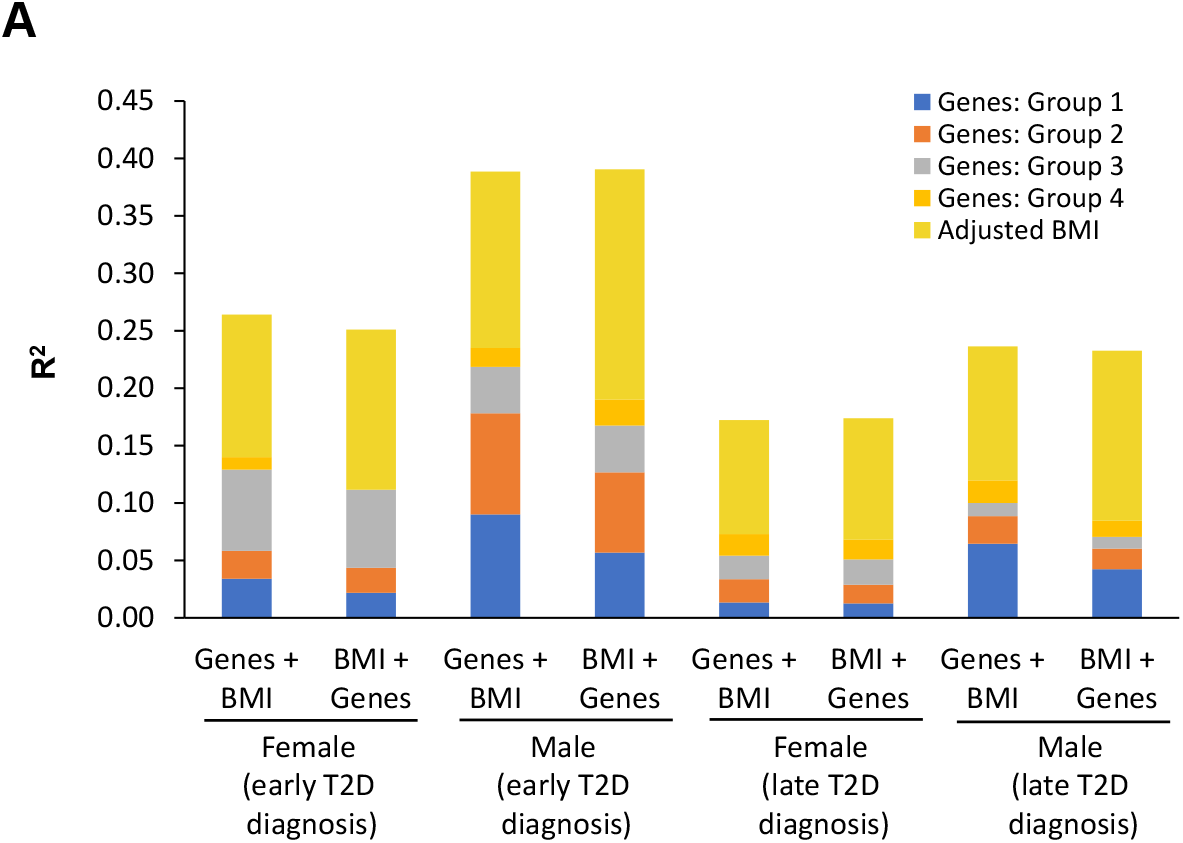
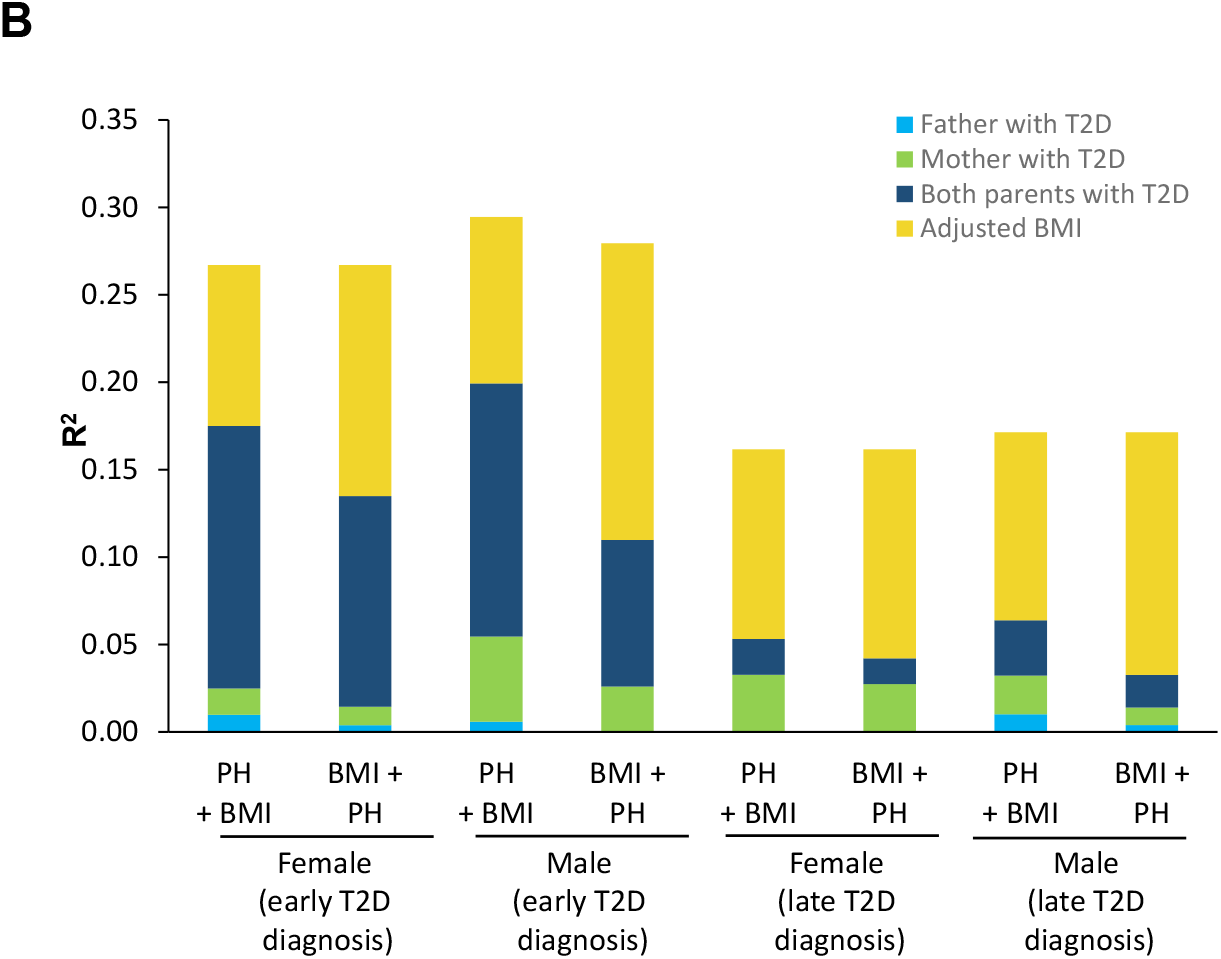
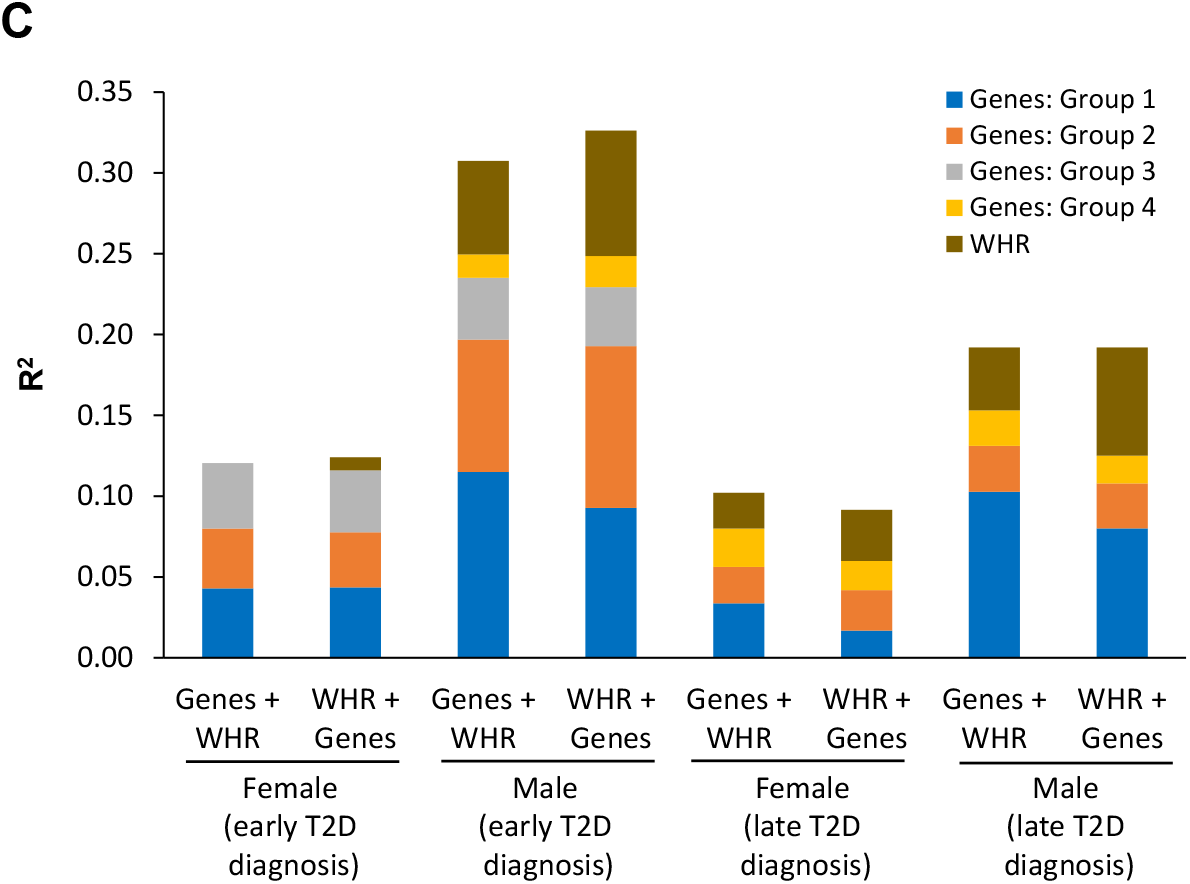
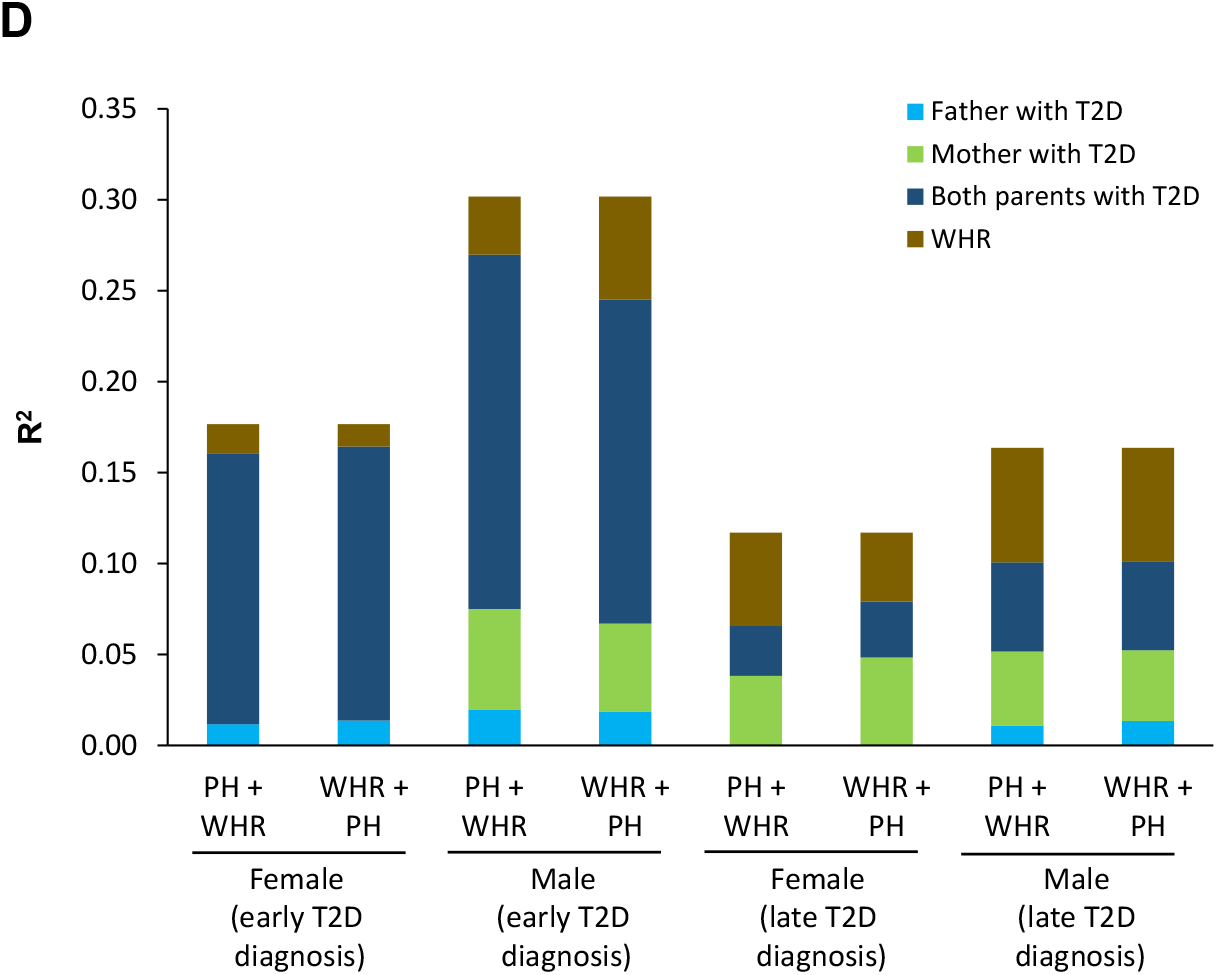
Influence (R^2^) of obesity and fat distribution and genes and parental history on T2D development overlap. Overlapping analysis was performed with stratification by age of T2D diagnosis and sex. Overlapping analysis of (A) BMI with genes, (B) BMI with parental history, (C) WHR with genes, and (D) WHR with parental history are shown. The overlapping effect of R^2^ between genes and BMI and between parental history and BMI was observed by introducing BMI as the first block and either genes or parental history as the second block and then conducting the analysis with the blocks reversed. Abbreviations: BMI, body mass index; PH, parental history; T2D, type 2 diabetes; WHR, weight distribution.

Using MLR, the effects of genes and parental history were analyzed separately to determine how much each individually absorbs the effect of BMI (first block, genes or parental history, second block, BMI; first block, BMI, second block, genes or parental history). In males diagnosed early, parental history or genes decreased the effect of BMI by 43.6% or 23.3%, respectively. With parental history as the first block, the effect was mainly absorbed by having a mother with T2D (87.2% increase) or both parents with T2D (72% increase); the effect of a father with T2D was minor (**Figure 5B**). With genes as the first block, the effect of BMI was mainly absorbed by genes located in the chromosome region 11p15.5 (58.5% increase) (**Figure 5A**). Similar increases with parental history or genes as the first block were observed in males diagnosed late and females diagnosed early (**Figure 5A** and **Figure 5B**). A similar analysis was performed using WHR rather than BMI. The findings in males diagnosed early were similar to those found for BMI (**Figure 5C** and **Figure 5D**) and together these results suggest that some components of parental history, and to a lesser extent genes on the chromosome 11p15.5 region, are linked to obesity.

Using the Pearson correlation, we found a positive correlation between the number of risk alleles in *KCNQ1* (chromosome 11p15.5 region) and both BMI and WHR in males diagnosed early (r = 0.125, *P* = 0.001 and r = 0.130, *P* < 0.009, respectively) and late (r = 0.103, *P* < 0.007 and r = 0.140, *P* < 0.003, respectively) (**Figure 6**). A positive correlation was also observed between SNP in *INS* and BMI in males diagnosed early (r = 0.09, *P* = 0.019; data not shown).

**Figure 6.**
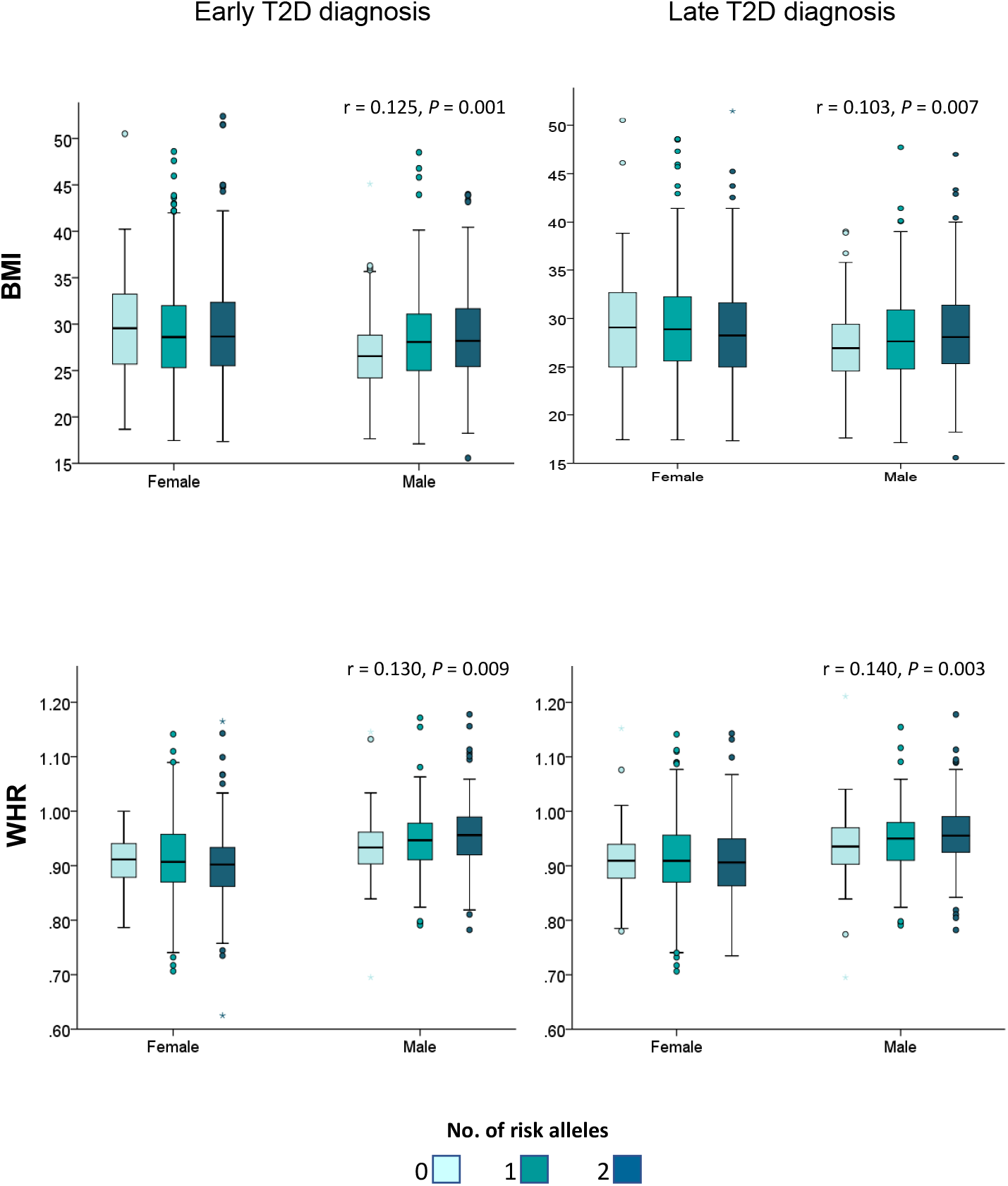
Correlation analysis between the number of risk alleles in *KCNQ1* in relation to obesity and fat distribution. (A) Correlation between the number of risk alleles in *KCNQ1* and BMI in males and females with a T2D diagnosis ≤45 years and ≥46 years of age. (B) Correlation between the number of risk alleles in *KCNQ1* and WHR in males and females with a T2D diagnosis ≤45 years and ≥46 years of age. Correlations and *P* values were calculated using the Pearson correlation test. Abbreviations: BMI, body mass index; T2D, type 2 diabetes; WHR, weight distribution.

When BMI was introduced as the last block, the influence of BMI (R^2^) was similar among the four groups of T2D (R^2^ range: 0.078–0.094). However, the strength of the BMI effect related to parental history and genes appeared stronger for a late diagnosis compared with an early diagnosis (**Figure 4B**), indicating that obesity is more important than genetic influence and parental history for males and females diagnosed with late-onset T2D.

## Discussion

This study identified notable differences in the association of T2D-related genes, parental history of T2D, BMI, and WHR with T2D development between males and females of Latin American mestizo origin. When participants were stratified by age of T2D presentation, the differences observed between males and females remained regardless of whether they were diagnosed with T2D early (≤45 years) or late (≥46 years). However, in males and females diagnosed late, the influence of genes and family history decreased drastically, while the influence of obesity increased. The reduced effect of T2D-related genes and parental history in late-onset T2D has previously been reported in other studies on White populations (27, 28). The three factors (genetic loci, parental history, and BMI) appear related to each other, given that they each compete for a percentage of R^2^ (percent of T2D variability explained by each factor). In fact, the relationship of these three factors seems to be much more important in this studied population than in previously analyzed European populations (29) The differences between males and females were most evident in those diagnosed with T2D early: 1) the genetic contribution involved in insulin production was predominant in males while genes involved in peripheral resistance were more important in females, 2) the influence of obesity, especially WHR, was much greater in males than females, and 3) the influence of a mother (unilateral) with a history of T2D greatly influenced males. Given genes that are involved in insulin production are located in autosomes, differences in T2D association should not be related to a differential distribution of risk alleles between males and females. Rather, this is most likely associated with external factors that differentially influence these genes in males and females. Our findings suggest that obesity, hormones, and maternal inheritance may be involved in the selection of males, but not females, who have risk alleles involved in insulin production for the early development of T2D. The association of BMI and especially WHR was much higher in males than females in the group that was diagnosed with T2D early (WHR 16× higher). Also, in males there was a linear correlation between BMI and WHR and the number of risk alleles in several genes located in the chromosome region 11p15.5 (*INS* and *KCNQ1*) that are associated with insulin production. There is clear evidence that sex hormones play an important role in fat distribution, measured as WHR (30). WHR is a measurement of visceral fat (waist circumference) and gluteal fat (hip circumference). Gluteal-femoral fat predominates in females prior to menopause whereas abdominal fat predominates in postmenopausal females and males of all ages. Abdominal fat has been associated with an increased risk of developing T2D and cardiovascular disease (31). This may be contributing to the lower risks of developing T2D in premenopausal females compared with males of the same age (32).

The overall prevalence of T2D is slightly higher in males than females (33-35); in the 2021 International Diabetes Federation report, the prevalence of T2D in males and females 20–79 years of age was 10.8% and 10.2%, respectively (17.7 million more males than females with T2D). However, this difference is not uniform across age ranges; the difference is greatest among those ≤55 years of age (approximately 16.5% versus 13.5%) compared with those who are older, for whom the difference disappears with increasing age (17, 34-36). In Mexico, this difference appears even greater. For example, in 2019 the incidence of T2D was 17.2% higher in males than females aged 15–49 years (524 versus 447 cases/100,000 persons) (37). This lower prevalence in younger females coincides with the premenopausal period, suggesting a protective effect for estrogens. This protective role has been demonstrated in studies of estradiol (38). Hormone replacement therapy is reported to reduce the incidence of T2D by 35% versus placebo in postmenopausal females (39) and early menopause is associated with an increased risk of T2D (40). Fasting glucose levels in premenopausal females are lower than those in age- and weight-matched males (41), while rat models have shown that estrogens protect against T2D by increasing insulin sensitivity (42, 43) and insulin secretion (43), and decreasing oxidative stress (42). This may suggest that a hormonal effect may protect or delay the impact of genes on the development of T2D in females. Furthermore, overweight or obese males may have low concentrations of serum testosterone, which is associated with an increased risk of T2D (44). A study of males aged 50-74 years, with a waist circumference ≥95 cm, serum testosterone concentration ≤14.0 nmol/L, and impaired glucose tolerance or newly diagnosed T2D investigated the effects of participation in a lifestyle program with or without testosterone treatment on T2D (45). The study found that those who received testosterone treatment for 2 years had a greater reduction in 2-hour glucose versus baseline than those who received placebo (mean difference −0.75 mmol/L; p<0.0001). Pancreatic islets have also been shown to be more susceptible to oxidative stress in males than females (46).

It has previously been shown that the influence of maternal history of diabetes is more important than paternal influence, at least for the early development of T2D (47). In the present study, we observed that the parental history of T2D through the mother (unilateral) may play an important role in differences in the association of risk alleles between the sexes in early-diagnosed T2D for two reasons: 1) MLR analysis revealed a substantial reduction in the influence of a mother with T2D when the groups of genes involved in insulin secretion were included in the first block and parental inheritance in the second, and 2) the distribution of risk alleles in *INS* and *KCNQ1* (chromosome 11p15.5 region) and *TCF7L2* was significantly different between cases and controls only when the mother had a history of T2D (not with the father or both parents). At least in the case of the chromosome 11p15.5 region, there is evidence of genetic imprinting on the maternal side; a previous study reported that risk alleles in *KCNQ1* conferred a risk for T2D only when are inherited by the mother (26) and that risk alleles influenced methylation levels of a regulatory sequence in fetal human pancreas, suggesting that diabetes risk effects may be mediated in early development (48). Interestingly, the effect of maternal inheritance on genes from that region was only observed in males with an early T2D diagnosis. Such an association has not been previously reported.

There were also important differences in the T2D models of males and females with a late T2D diagnosis. The effect size of T2D-related genes was larger in males than females, primarily at the expense of the genes involved with insulin production from the chromosome 11p15.5 region. It is also noteworthy that in both groups with a late diagnosis, the proportion of the participation of genes involved with inflammation and other processes, relative to the total variance due to genes, was higher than in those with an early diagnosis. This could be related to increased inflammatory processes with aging (49).

This study had some limitations. As noted, the number of participants with missing data for WHR was notable. However, the important difference in the median WHR and the degree of association with T2D between men and women, as well as the significant correlation between WHR and the number of risk alleles of the *KCNQ1* gene, are very notable. These data could be considered valid because they were explored in ULR analyses. However, it is clear that the interpretation of the effect of WHR in MLR models should be confirmed in studies with a larger sample of cases and controls. Furthermore, this study is also limited by the inclusion of only Mexican participants and therefore the study findings are not necessarily generalizable to other populations.

The previous results allow us to conclude that the influence of T2D-related genes, maternal T2D history, and fat distribution on T2D development was greater in males than females.

## Supporting information

STROBE Checklist

## Data Availability

All data generated or analyzed during this study are included in this published article and its supplementary information files.

## Acknowledgements

The authors thank Sarah Bubeck, PhD, of Edanz (www.edanz.com) for providing medical writing support, which was funded by Fundación Carlos Slim, in accordance with Good Publication Practice (GPP3) guidelines (http://www.ismpp.org/gpp3).

## Author contributions

J.B. and R.T.C. made substantial contributions to the conceptualization of the study. J.B., L.O., H.G.R., A.M., and R.T.C. contributed to the study methodology. F.R. utilized software. J.B. performed the formal analysis. H.G.O., M.M.M., E.B., E.J.T., A.A.S, L.A.M.J., and D.A.A.H. were involved in the study investigation. L.O., R.E.B., and H.G.O. provided resources for the study. J.B., H.G.R., L.A.M.J., A.M., D.A.A.H., and R.T.C were involved in visualization of the study data. J.B., H.G.R., and R.T.C. supervised the study. R.E.B. was involved in project administration and J.B. and R.T.C. acquired funding for the study. J.B. wrote the original draft and all authors reviewed and edited the manuscript.

## References

1. Zheng Y, Ley SH, Hu FB. Global aetiology and epidemiology of type 2 diabetes mellitus and its complications. Nat Rev Endocrinol. 2018;14(2):88–98.

2. Carey VJ, Walters EE, Colditz GA, Solomon CG, Willett WC, Rosner BA, Speizer FE, Manson JE. Body fat distribution and risk of non-insulin-dependent diabetes mellitus in women. The Nurses’ Health Study. Am J Epidemiol. 1997;145(7):614–619.

3. Hu FB. Obesity epidemiology. Oxford: Oxford University Press; 2008.

4. Zheng Y, Manson JE, Yuan C, Liang MH, Grodstein F, Stampfer MJ, Willett WC, Hu FB. Associations of weight gain from early to middle adulthood with major health outcomes later in life. JAMA. 2017;318(3):255–269.

5. Berumen J, Orozco L, Betancourt-Cravioto M, Gallardo H, Zulueta M, Mendizabal L, Simon L, Benuto RE, Ramírez-Campos E, Marin M, Juárez E, García-Ortiz H, Martínez-Hernández A, Venegas-Vega C, Peralta-Romero J, Cruz M, Tapia-Conyer R. Influence of obesity, parental history of diabetes, and genes in type 2 diabetes: A case-control study. Sci Rep. 2019;9(1):2748.

6. Almgren P, Lehtovirta M, Isomaa B, Sarelin L, Taskinen MR, Lyssenko V, Tuomi T, Groop L; Botnia Study Group. Heritability and familiality of type 2 diabetes and related quantitative traits in the Botnia Study. Diabetologia. 2011;54(11):2811–2819.

7. Jang HM, Hwang MY, Kim BJ, Kim YJ. Validation and genetic heritability estimation of known type 2 diabetes related variants in the Korean population. Genomics Inform. 2021;19(4):e37.

8. Chi Y, Wang X, Jia J, Huang T. Smoking status and type 2 diabetes, and cardiovascular dsease: A comprehensive analysis of shared genetic etiology and causal relationship. Front Endocrinol (Lausanne). 2022;13:809445.

9. Fuchsberger C, Flannick J, Teslovich TM, Mahajan A, Agarwala V, Gaulton KJ, Ma C, Fontanillas P, Moutsianas L, McCarthy DJ, Rivas MA, Perry JRB, Sim X, Blackwell TW, Robertson NR, Rayner NW, Cingolani P, Locke AE, Tajes JF, Highland HM, Dupuis J, Chines PS, Lindgren CM, Hartl C, Jackson AU, Chen H, Huyghe JR, van de Bunt M, Pearson RD, Kumar A, Müller-Nurasyid M, Grarup N, Stringham HM, Gamazon ER, Lee J, Chen Y, Scott RA, Below JE, Chen P, Huang J, Go MJ, Stitzel ML, Pasko D, Parker SCJ, Varga TV, Green T, Beer NL, Day-Williams AG, Ferreira T, Fingerlin T, Horikoshi M, Hu C, Huh I, Ikram MK, Kim BJ, Kim Y, Kim YJ, Kwon MS, Lee J, Lee S, Lin KH, Maxwell TJ, Nagai Y, Wang X, Welch RP, Yoon J, Zhang W, Barzilai N, Voight BF, Han BG, Jenkinson CP, Kuulasmaa T, Kuusisto J, Manning A, Ng MCY, Palmer ND, Balkau B, Stančáková A, Abboud HE, Boeing H, Giedraitis V, Prabhakaran D, Gottesman O, Scott J, Carey J, Kwan P, Grant G, Smith JD, Neale BM, Purcell S, Butterworth AS, Howson JMM, Lee HM, Lu Y, Kwak SH, Zhao W, Danesh J, Lam VKL, Park KS, Saleheen D, So WY, Tam CHT, Afzal U, Aguilar D, Arya R, Aung T, Chan E, Navarro C, Cheng CY, Palli D, Correa A, Curran JE, Rybin D, Farook VS, Fowler SP, Freedman BI, Griswold M, Hale DE, Hicks PJ, Khor CC, Kumar S, Lehne B, Thuillier D, Lim WY, Liu J, van der Schouw YT, Loh M, Musani SK, Puppala S, Scott WR, Yengo L, Tan ST, Taylor HA Jr, Thameem F, Wilson G Sr, Wong TY, Njølstad PR, Levy JC, Mangino M, Bonnycastle LL, Schwarzmayr T, Fadista J, Surdulescu GL, Herder C, Groves CJ, Wieland T, Bork-Jensen J, Brandslund I, Christensen C, Koistinen HA, Doney ASF, Kinnunen L, Esko T, Farmer AJ, Hakaste L, Hodgkiss D, Kravic J, Lyssenko V, Hollensted M, Jørgensen ME, Jørgensen T, Ladenvall C, Justesen JM, Käräjämäki A, Kriebel J, Rathmann W, Lannfelt L, Lauritzen T, Narisu N, Linneberg A, Melander O, Milani L, Neville M, Orho-Melander M, Qi L, Qi Q, Roden M, Rolandsson O, Swift A, Rosengren AH, Stirrups K, Wood AR, Mihailov E, Blancher C, Carneiro MO, Maguire J, Poplin R, Shakir K, Fennell T, DePristo M, de Angelis MH, Deloukas P, Gjesing AP, Jun G, Nilsson P, Murphy J, Onofrio R, Thorand B, Hansen T, Meisinger C, Hu FB, Isomaa B, Karpe F, Liang L, Peters A, Huth C, O’Rahilly SP, Palmer CNA, Pedersen O, Rauramaa R, Tuomilehto J, Salomaa V, Watanabe RM, Syvänen AC, Bergman RN, Bharadwaj D, Bottinger EP, Cho YS, Chandak GR, Chan JCN, Chia KS, Daly MJ, Ebrahim SB, Langenberg C, Elliott P, Jablonski KA, Lehman DM, Jia W, Ma RCW, Pollin TI, Sandhu M, Tandon N, Froguel P, Barroso I, Teo YY, Zeggini E, Loos RJF, Small KS, Ried JS, DeFronzo RA, Grallert H, Glaser B, Metspalu A, Wareham NJ, Walker M, Banks E, Gieger C, Ingelsson E, Im HK, Illig T, Franks PW, Buck G, Trakalo J, Buck D, Prokopenko I, Mägi R, Lind L, Farjoun Y, Owen KR, Gloyn AL, Strauch K, Tuomi T, Kooner JS, Lee JY, Park T, Donnelly P, Morris AD, Hattersley AT, Bowden DW, Collins FS, Atzmon G, Chambers JC, Spector TD, Laakso M, Strom TM, Bell GI, Blangero J, Duggirala R, Tai ES, McVean G, Hanis CL, Wilson JG, Seielstad M, Frayling TM, Meigs JB, Cox NJ, Sladek R, Lander ES, Gabriel S, Burtt NP, Mohlke KL, Meitinger T, Groop L, Abecasis G, Florez JC, Scott LJ, Morris AP, Kang HM, Boehnke M, Altshuler D, McCarthy MI. The genetic architecture of type 2 diabetes. Nature. 2016;536(7614):41–47.

10. Manolio TA, Collins FS, Cox NJ, Goldstein DB, Hindorff LA, Hunter DJ, McCarthy MI, Ramos EM, Cardon LR, Chakravarti A, Cho JH, Guttmacher AE, Kong A, Kruglyak L, Mardis E, Rotimi CN, Slatkin M, Valle D, Whittemore AS, Boehnke M, Clark AG, Eichler EE, Gibson G, Haines JL, Mackay TF, McCarroll SA, Visscher PM. Finding the missing heritability of complex diseases. Nature. 2009;461(7265):747–753.

11. Wang L, Lee S, Gim J, Qiao D, Cho M, Elston RC, Silverman EK, Won S. Family-based rare variant association analysis: A fast and efficient method of multivariate phenotype association analysis. Genet Epidemiol. 2016;40(6):502–511.

12. Ma Y, Zhou Z, Li X, Yan Z, Ding K, Xiao H, Wu Y, Wu T, Chen D. Integrative identification of genetic loci jointly influencing diabetes-related traits and sleep traits of insomnia, sleep duration, and chronotypes. Biomedicines. 2022;10(2):368.

13. Mahajan A, Taliun D, Thurner M, Robertson NR, Torres JM, Rayner NW, Payne AJ, Steinthorsdottir V, Scott RA, Grarup N, Cook JP, Schmidt EM, Wuttke M, Sarnowski C, Mägi R, Nano J, Gieger C, Trompet S, Lecoeur C, Preuss MH, Prins BP, Guo X, Bielak LF, Below JE, Bowden DW, Chambers JC, Kim YJ, Ng MCY, Petty LE, Sim X, Zhang W, Bennett AJ, Bork-Jensen J, Brummett CM, Canouil M, Ec Kardt KU, Fischer K, Kardia SLR, Kronenberg F, Läll K, Liu CT, Locke AE, Luan J, Ntalla I, Nylander V, Schönherr S, Schurmann C, Yengo L, Bottinger EP, Brandslund I, Christensen C, Dedoussis G, Florez JC, Ford I, Franco OH, Frayling TM, Giedraitis V, Hackinger S, Hattersley AT, Herder C, Ikram MA, Ingelsson M, Jørgensen ME, Jørgensen T, Kriebel J, Kuusisto J, Ligthart S, Lindgren CM, Linneberg A, Lyssenko V, Mamakou V, Meitinger T, Mohlke KL, Morris AD, Nadkarni G, Pankow JS, Peters A, Sattar N, Stančáková A, Strauch K, Taylor KD, Thorand B, Thorleifsson G, Thorsteinsdottir U, Tuomilehto J, Witte DR, Dupuis J, Peyser PA, Zeggini E, Loos RJF, Froguel P, Ingelsson E, Lind L, Groop L, Laakso M, Collins FS, Jukema JW, Palmer CNA, Grallert H, Metspalu A, Dehghan A, Köttgen A, Abecasis GR, Meigs JB, Rotter JI, Marchini J, Pedersen O, Hansen T, Langenberg C, Wareham NJ, Stefansson K, Gloyn AL, Morris AP, Boehnke M, McCarthy MI. Fine-mapping type 2 diabetes loci to single-variant resolution using high-density imputation and islet-specific epigenome maps. Nat Genet. 2018;50(11):1505–1513.

14. Teslovich TM, Musunuru K, Smith AV, Edmondson AC, Stylianou IM, Koseki M, Pirruccello JP, Ripatti S, Chasman DI, Willer CJ, Johansen CT, Fouchier SW, Isaacs A, Peloso GM, Barbalic M, Ricketts SL, Bis JC, Aulchenko YS, Thorleifsson G, Feitosa MF, Chambers J, Orho-Melander M, Melander O, Johnson T, Li X, Guo X, Li M, Shin Cho Y, Jin Go M, Jin Kim Y, Lee JY, Park T, Kim K, Sim X, Twee-Hee Ong R, Croteau-Chonka DC, Lange LA, Smith JD, Song K, Hua Zhao J, Yuan X, Luan J, Lamina C, Ziegler A, Zhang W, Zee RY, Wright AF, Witteman JC, Wilson JF, Willemsen G, Wichmann HE, Whitfield JB, Waterworth DM, Wareham NJ, Waeber G, Vollenweider P, Voight BF, Vitart V, Uitterlinden AG, Uda M, Tuomilehto J, Thompson JR, Tanaka T, Surakka I, Stringham HM, Spector TD, Soranzo N, Smit JH, Sinisalo J, Silander K, Sijbrands EJ, Scuteri A, Scott J, Schlessinger D, Sanna S, Salomaa V, Saharinen J, Sabatti C, Ruokonen A, Rudan I, Rose LM, Roberts R, Rieder M, Psaty BM, Pramstaller PP, Pichler I, Perola M, Penninx BW, Pedersen NL, Pattaro C, Parker AN, Pare G, Oostra BA, O’Donnell CJ, Nieminen MS, Nickerson DA, Montgomery GW, Meitinger T, McPherson R, McCarthy MI, McArdle W, Masson D, Martin NG, Marroni F, Mangino M, Magnusson PK, Lucas G, Luben R, Loos RJ, Lokki ML, Lettre G, Langenberg C, Launer LJ, Lakatta EG, Laaksonen R, Kyvik KO, Kronenberg F, König IR, Khaw KT, Kaprio J, Kaplan LM, Johansson A, Jarvelin MR, Janssens AC, Ingelsson E, Igl W, Kees Hovingh G, Hottenga JJ, Hofman A, Hicks AA, Hengstenberg C, Heid IM, Hayward C, Havulinna AS, Hastie ND, Harris TB, Haritunians T, Hall AS, Gyllensten U, Guiducci C, Groop LC, Gonzalez E, Gieger C, Freimer NB, Ferrucci L, Erdmann J, Elliott P, Ejebe KG, Döring A, Dominiczak AF, Demissie S, Deloukas P, de Geus EJ, de Faire U, Crawford G, Collins FS, Chen YD, Caulfield MJ, Campbell H, Burtt NP, Bonnycastle LL, Boomsma DI, Boekholdt SM, Bergman RN, Barroso I, Bandinelli S, Ballantyne CM, Assimes TL, Quertermous T, Altshuler D, Seielstad M, Wong TY, Tai ES, Feranil AB, Kuzawa CW, Adair LS, Taylor HA Jr, Borecki IB, Gabriel SB, Wilson JG, Holm H, Thorsteinsdottir U, Gudnason V, Krauss RM, Mohlke KL, Ordovas JM, Munroe PB, Kooner JS, Tall AR, Hegele RA, Kastelein JJ, Schadt EE, Rotter JI, Boerwinkle E, Strachan DP, Mooser V, Stefansson K, Reilly MP, Samani NJ, Schunkert H, Cupples LA, Sandhu MS, Ridker PM, Rader DJ, van Duijn CM, Peltonen L, Abecasis GR, Boehnke M, Kathiresan S. Biological, clinical and population relevance of 95 loci for blood lipids. Nature. 2010;466(7307):707–713.

15. Piko P, Werissa NA, Fiatal S, Sandor J, Adany R. Impact of genetic factors on the age of onset for type 2 diabetes mellitus in addition to the conventional risk factors. J Pers Med. 2020;11(1):6.

16. Humphries SE, Gable D, Cooper JA, Ireland H, Stephens JW, Hurel SJ, Li KW, Palmen J, Miller MA, Cappuccio FP, Elkeles R, Godsland I, Miller GJ, Talmud PJ. Common variants in the TCF7L2 gene and predisposition to type 2 diabetes in UK European Whites, Indian Asians and Afro-Caribbean men and women. J Mol Med (Berl). 2006;84(12)1005–1014.

17. Kautzky-Willer A, Harreiter J, Pacini G. Sex and gender differences in risk, pathophysiology and complications of type 2 diabetes mellitus. Endocr Rev. 2016;37(3):278–316.

18. SIGMA Type 2 Diabetes Consortium, Williams AL, Jacobs SB, Moreno-Macías H, Huerta-Chagoya A, Churchhouse C, Márquez-Luna C, García-Ortíz H, Gómez-Vázquez MJ, Burtt NP, Aguilar-Salinas CA, González-Villalpando C, Florez JC, Orozco L, Haiman CA, Tusié-Luna T, Altshuler D. Sequence variants in SLC16A11 are a common risk factor for type 2 diabetes in Mexico. Nature. 2014;506(7486)97–101.

19. Mercader JM, Florez JC. The genetic basis of type 2 diabetes in Hispanics and Latin Americans: Challenges and opportunities. Front Public Health. 2017;5:329.

20. SIGMA Type 2 Diabetes Consortium, Estrada K, Aukrust I, Bjørkhaug L, Burtt NP, Mercader JM, García-Ortiz H, Huerta-Chagoya A, Moreno-Macías H, Walford G, Flannick J, Williams AL, Gómez-Vázquez MJ, Fernandez-Lopez JC, Martínez-Hernández A, Jiménez-Morales S, Centeno-Cruz F, Mendoza-Caamal E, Revilla-Monsalve C, Islas-Andrade S, Córdova EJ, Soberón X, González-Villalpando ME, Henderson E, Wilkens LR, Le Marchand L, Arellano-Campos O, Ordóñez-Sánchez ML, Rodríguez-Torres M, Rodríguez-Guillén R, Riba L, Najmi LA, Jacobs SB, Fennell T, Gabriel S, Fontanillas P, Hanis CL, Lehman DM, Jenkinson CP, Abboud HE, Bell GI, Cortes ML, Boehnke M, González-Villalpando C, Orozco L, Haiman CA, Tusié-Luna T, Aguilar-Salinas CA, Altshuler D, Njølstad PR, Florez JC, MacArthur DG. Association of a low-frequency variant in HNF1A with type 2 diabetes in a Latino population. JAMA. 2014;311(22):2305–2314.

21. Mercader JM, Liao RG, Bell AD, Dymek Z, Estrada K, Tukiainen T, Huerta-Chagoya A, Moreno-Macías H, Jablonski KA, Hanson RL, Walford GA, Moran I, Chen L, Agarwala V, Ordoñez-Sánchez ML, Rodríguez-Guillen R, Rodríguez-Torres M, Segura-Kato Y, García-Ortiz H, Centeno-Cruz F, Barajas-Olmos F, Caulkins L, Puppala S, Fontanillas P, Williams AL, Bonàs-Guarch S Hartl c, Ripke S; Diabetes Prevention Program Research Group, Tooley K, Lane J, Zerrweck C, Martínez-Hernández A, Córdova EJ, Mendoza-Caamal E, Contreras-Cubas C, González-Villalpando Me, Cruz-Bautista I, Muñoz-Hernández L, Gómez-Velasco D, Alvirde U, Henderson BE, Wilkens LR, Le Marchand L, Arellano-Campos O, Riba L, Harden M; Broad Genomics Platform Gabriel S; T2D-GENES Consortium, Abboud HE, Cortes ML, Revilla-Monsalve C, Islas-Andrade S, Soberon X, Curran JE, Jenkinson CP, DeFronzo RA, Lehman DM, Hanis CL, Bell GI, Boehnke M, Blangero J, Duggirala R, Saxena R, MacArthur D, Ferrer J, McCarroll SA, Torrents D, Knowler WC, Baier LJ, Burtt N, González-Villalpando C, Haiman CA, Aguilar-Salinas CA, Tusié-Luna T, Flannick J, Jacobs SBR, Orozco L, Altshuler D, Florez JC; SIGMA T2D Genetics Consortium. A loss-of-function splice acceptor variant in IGF2 is protective for type 2 diabetes. Diabetes. 2017;66(11): 2903–2914.

22. American Diabetes Association. Standards of Medical Care in Diabetes-2022 Abridged for Primary Care Providers. Clin Diabetes. 2022;40(1):10–38.

23. Babyak MA. What you see may not be what you get: a brief, nontechnical introduction to overfitting in regression-type models. Psychosom Med. 2004;66(3):411–421.

24. Nagelkerke NJD. A note on a general definition of the coefficient of determination. Biometrika. 1991;78(3):691–692.

25. Faul F, Erdfelder E, Lang AG, Buchner A. G*Power 3: a flexible statistical power analysis program for the social, behavioral, and biomedical sciences. Behav Res Methods. 2007;39(2):175–191.

26. Kong A, Steinthorsdottir V, Masson G, Thorleifsson G, Sulem P, Besenbacher S, Jonasdottir A, Sigurdsson A, Kristinsson KT, Jonasdottir A, Frigge ML, Gylfason A, Olason PI, Gudjonsson SA, Sverrisson S, Stacey SN, Sigurgeirsson B, Benediktsdottir KR, Sigurdsson H, Jonsson T, Benediktsson R, Olafsson JH, Johannsson OT, Hreidarsson AB, Sigurdsson G; DIAGRAM Consortium, Ferguson-Smith AC, Gudbjartsson DF, Thorsteinsdottir U, Stefansson K. Parental origin of sequence variants associated with complex diseases. Nature. 2009;462(7275):868–874.

27. Ding M, Ahmad S, Qi L, Hu Y, Bhupathiraju SN, Guasch-Ferré M, Jensen MK, Chavarro JE, Ridker PM, Willett WC, Chasman DI, Hu FB, Kraft P. Additive and multiplicative interactions between genetic risk score and family history and lifestyle in relation to risk of type 2 diabetes. Am J Epidemiol. 2020;189(5):445–460.

28. Amuta AO, Mkuu R, Jacobs W, Barry AE. Number and severity of type 2 diabetes among family members are associated with nutrition and physical activity behaviors. Front Public Health. 2017;5:157.

29. InterAct Consortium, Scott RA, Langenberg C, Sharp SJ, Franks PW, Rolandsson O, Drogan D, van der Schouw YT, Ekelund U, Kerrison ND, Ardanaz E, Arriola L, Balkau B, Barricarte A, Barroso I, Bendinelli B, Beulens JW, Boeing H, de Lauzon-Guillain B, Deloukas P, Fagherazzi G, Gonzalez C, Griffin SJ, Groop LC, Halkjaer J, Huerta JM, Kaaks R, Khaw KT, Krogh V, Nilsson PM, Norat T, Overvad K, Panico S, Rodriguez-Suarez L, Romaguera D, Romieu I, Sacerdote C, Sánchez MJ, Spijkerman AM, Teucher B, Tjonneland A, Tumino R, van der A DL, Wark PA, McCarthy MI, Riboli E, Wareham NJ. The link between family history and risk of type 2 diabetes is not explained by anthropometric, lifestyle or genetic risk factors: the EPIC-InterAct study. Diabetologia. 2013;56(1):60–69.

30. Watts EL, Appleby PN, Albanes D, Black A, Chan JM, Chen C, Cirillo PM, Cohn BA, Cook MB, Donovan JL, Ferrucci L, Garland CF, Giles GG, Goodman PJ, Habel LA, Haiman CA, Holly JMP, Hoover RN, Kaaks R, Knekt P, Kolonel LN, Kubo T, Le Marchand L, Luostarinen T, MacInnis RJ, Mäenpää HO, Männistö S, Metter EJ, Milne RL, Nomura AMY, Oliver SE, Parsons JK, Peeters PH, Platz EA, Riboli E, Ricceri F, Rinaldi S, Rissanen H, Sawada N, Schaefer CA, Schenk JM, Stanczyk FZ, Stampfer M, Stattin P, Stenman UH, Tjønneland A, Trichopoulou A, Thompson IM, Tsugane S, Vatten L, Whittemore AS, Ziegler RG, Allen NE, Key TJ, Travis RC. Circulating sex hormones in relation to anthropometric, sociodemographic and behavioural factors in an international dataset of 12,300 men. PLoS One. 2017;12(12): e0187741.

31. Benz V, Kintscher U, Foryst-Ludwig A. Sex-specific differences in Type 2 Diabetes Mellitus and dyslipidemia therapy: PPAR agonists. Handb Exp Pharmacol. 2012;(214):387–410.

32. Zhang H, Sairam, MR. Sex hormone imbalances and adipose tissue dysfunction impacting on metabolic syndrome; a paradigm for the discovery of novel adipokines. Horm Mol Biol Clin Investig. 2014;17(2):89–97.

33. Wild S, Roglic G, Green A, Sicree R, King H. Global prevalence of diabetes: estimates for the year 2000 and projections for 2030. Diabetes Care. 2004;27(5):1047–1053.

34. Khan MAB, Hashim MJ, King JK, Govender RD, Mustafa H, Al Kaabi J. Epidemiology of type 2 diabetes - Global burden of disease and forecasted trends. J Epidemiol Glob Health. 2020;10(1):107–111.

35. International Diabetes Federation. IDF Diabetes Atlas 2021: IDF Atlas. 10th Edition. Available from: https://diabetesatlas.org/atlas/tenth-edition/.

36. Wändell PE, Carlsson AC. Gender differences and time trends in incidence and prevalence of type 2 diabetes in Sweden--a model explaining the diabetes epidemic worldwide today? Diabetes Res Clin Pract. 2014;106(3):e90–92.

37. Institute for Health Metrics and Evaluation. GBD 2019. Available from: http://ihmeuw.org/5pb2.

38. Fappi A, Mittendorfer B. Different physiological mechanisms underlie an adverse cardiovascular disease risk profile in men and women. Proc Nutr Soc. 2020;79(2):210–218.

39. Kanaya AM, Herrington D, Vittinghoff E, Lin F, Grady D, Bittner V, Cauley JA, Barrett-Connor E; Heart and Estrogen/progestin Replacement Study. Glycemic effects of postmenopausal hormone therapy: the Heart and Estrogen/progestin Replacement Study. A randomized, double-blind, placebo-controlled trial. Ann Intern Med. 2003;138(1):1–9.

40. Brand JS, van der Schouw YT, Onland-Moret NC, Sharp SJ, Ong KK, Khaw KT, Ardanaz E, Amiano P, Boeing H, Chirlaque MD, Clavel-Chapelon F, Crowe FL, de Lauzon-Guillain B, Duell EJ, Fagherazzi G, Franks PW, Grioni S, Groop LC, Kaaks R, Key TJ, Nilsson PM, Overvad K, Palli D, Panico S, Quirós JR, Rolandsson O, Sacerdote C, Sánchez MJ, Slimani N, Teucher B, Tjonneland A, Tumino R, van der A DL, Feskens EJ, Langenberg C, Forouhi NG, Riboli E, Wareham NJ; InterAct Consortium. Age at menopause, reproductive life span, and type 2 diabetes risk: results from the EPIC-InterAct study. Diabetes Care. 2013;36(4):1012–1019.

41. Anderwald C, Gastaldelli A, Tura A, Krebs M, Promintzer-Schifferl M, Kautzky-Willer A, Stadler M, DeFronzo RA, Pacini G, Bischof MG. Mechanism and effects of glucose absorption during an oral glucose tolerance test among females and males. J Clin Endocrinol Metab. 2011;96(2):515–524.

42. Díaz A, López-Grueso R, Gambini J, Monleón D, Mas-Bargues C, Abdelaziz KM, Viña J, Borrás C. Sex differences in age-associated type 2 diabetes in rats-Role of estrogens and oxidative stress. Oxid Med Cell Longev. 2019;2019: 6734836.

43. Bian C, Bai B, Gao Q, Li S, Zhao Y. 17beta-estradiol regulates glucose metabolism and insulin secretion in rat islet beta cells through GPER and Akt/mTOR/GLUT2 pathway. Front Endocrinol (Lausanne). 2019;10:531.

44. Fui MN, Dupuis P, Grossmann M. Lowered testosterone in male obesity: mechanisms, morbidity and management. Asian J Androl. 2014;16(2):223–231.

45. Wittert G, Bracken K, Robledo KP, Grossmann M, Yeap BB, Handelsman DJ, Stuckey B, Conway A, Inder W, McLachlan R, Allan C, Jesudason D, Fui MNT, Hague W, Jenkins A, Daniel M, Gebski V, Keech A. Testosterone treatment to prevent or revert type 2 diabetes in men enrolled in a lifestyle programme (T4DM): a randomised, double-blind, placebo-controlled, 2-year, phase 3b trial. Lancet Diabetes Endocrinol. 2021;9(1):32–45.

46. Yokomizo H, Inoguchi T, Sonoda N, Sakaki Y, Maeda Y, Inoue T, Hirata E, Takei R, Ikeda N, Fujii M, Fukuda K, Sasaki H, Takayanagi R. Maternal high-fat diet induces insulin resistance and deterioration of pancreatic beta-cell function in adult offspring with sex differences in mice. Am J Physiol Endocrinol Metab. 2014;306(10):E1163–1175.

47. Meigs JB, Cupples LA, Wilson PW. Parental transmission of type 2 diabetes: the Framingham Offspring Study. Diabetes. 2000;49(12):2201–2207.

48. Travers ME, Mackay DJ, Dekker Nitert M, Morris AP, Lindgren CM, Berry A, Johnson PR, Hanley N, Groop LC, McCarthy MI, Gloyn AL. Insights into the molecular mechanism for type 2 diabetes susceptibility at the KCNQ1 locus from temporal changes in imprinting status in human islets. Diabetes. 2013;62(3):987–992.

49. Thakur N, Tiwari VK, Thomassin H, Pandey RR, Kanduri M, Göndör A, Grange T, Ohlsson R, Kanduri C. An antisense RNA regulates the bidirectional silencing property of the Kcnq1 imprinting control region. Mol Cell Biol. 2004;24(18):7855–7862.

