## Supplementary Tables S1-14 for "Sex-specific genetic loci linked to early and late onset type 2 diabetes"

**Supplementary Materials**

**Table S1.** List of 69 SNPs explored, organized by selection group

| **SNP** | **Chr** | **Position** | **Gene** | **Alt allele** | **Minor allele** | **MAF MEX** | **OR** | **P-WALD** | **Annotations** |
| --- | --- | --- | --- | --- | --- | --- | --- | --- | --- |
| **Selected from the Broad Institute database: https://t2d.hugeamp.org/** | | | | | | | | | |
| rs36113334 | 4 | 54937937 | *KDR* | C | G | 0.088 | 1.09 (0.8-1.5) | 0.584 | 5'-79 Kb-LINC02358/3'-6 Kb-AC111194.1 |
| rs42522 | 7 | 94411651 | *COL1A2* | A | G | 0.171 | 1.05 (0.8-1.3) | 0.685 | intronic,5upstream |
| **rs10511567** | 9 | 11606348 | *PTPRD* | T | C | 0.272 | 1.08 (0.9-1.3) | 0.455 | 5'-330 Kb-AL451129.1/3'-12 Kb-AL592227.1 |
| rs10961158 | 9 | 13592180 | *MPDZ* | G | A | 0.086 | 0.98 (0.7-1.4) | 0.901 | 5'-105 Kb-AL583785.1/3'- 336 Kb-LINC00583 |
| rs7018700 | 9 | 13971940 | *LINC00583* | T | A | 0.126 | 0.74 (0.5-1) | 0.045 | 5'-26 Kb-LINC00583/3'-14 Kb-PES1P2 |
| rs11198062 | 10 | 117659457 | *EMX2* | A | C | 0.189 | 0.94 (0.7-1.2) | 0.584 | 5'-87 Kb-AC005871.1/3'-76 Kb-AL139121.1 |
| rs80089797 | 11 | 2096447 | *IGF2* | C | T | 0.241 | 0.92 (0.7-1.1) | 0.436 | 5'-95 Kb-H19/3'-33 Kb-IGF2 |
| **rs163168** | 11 | 2803115 | *CDKN1C* | C | T | 0.412 | 0.71 (0.6-0.9) | 0.001 | non-coding intronic |
| **rs450208** | 11 | 2910751 | *SLC22A18* | T | G | 0.294 | 0.84 (0.7-1) | 0.091 | intronic,3downstream |
| rs12823525 | 12 | 95536905 | *USP44* | C | G | 0.216 | 0.85 (0.7-1.1) | 0.184 | intronic |
| rs889512 | 16 | 75208114 | *CTRB2* | C | G | 0.056 | 0.81 (0.5-1.2) | 0.332 | 5'-1 Kb-CTRB2/3'-11 Kb- CTRB1 |
| **rs689** | 11 | 2160994 | *INS* | T | A | 0.188 | 1.32 (1.1-1.5) | 0.00046 | splice region variant |
| rs9282541 | 9 | 104858554 | *ABCA1* | G | A | 0.114 | 0.99 (0.7-1.3) | 0.971 | **R230C** |
| rs4977038 | 8 | 142534540 | *ADGRB1* | T | C | 0.117 | 0.92 (0.7-1.2) | 0.584 | non-coding intronic |
| rs9701796 | 1 | 18859635 | *TAS1R2* | C | G | 0.17 | 0.94 (0.7-1.2) | 0.655 | **S9C** |
| **rs6577691** | 8 | 135961229 | *KHDRBS3* | T | G | 0.149 | 0.63 (0.5-0.8) | 0.002 | 5'-219 Kb-RNU1-35P/3'-86 Kb-AC079098.1 |
| rs28449777 | 8 | 142540000 | *ARC* | A | G | 0.137 | 0.84 (0.6-1.1) | 0.221 | intronic, non-coding |
| rs641959 | 9 | 133258308 | *OBP2B* | C | A | 0.422 | 0.87 (1-1.4) | 0.149 | non-coding intronic |
| rs7077032 | 10 | 45064532 | *C10orf25* | A | G | 0.252 | 0.94 (0.8-1.2) | 0.598 | non-coding intronic |
| rs16910359 | 11 | 12017672 | *DKK3* | G | A | 0.434 | 0.99 (0.8-1.2) | 0.944 | 5'-8 Kb-DKK3/3'-13 Kb- LINC02547 |
| rs1139971 | 11 | 44618718 | *CD82* | G | A | 0.364 | 1.1 (0.9-1.3) | 0.346 | **I216V/I241V** |
| rs4237808 | 12 | 53700013 | *CALCOCO1* | T | C | 0.329 | 0.99 (0.8-1.2) | 0.955 | 5'-23 Kb-ATP5G2/3'-9 Kb- CALCOCO1 |
| rs1147440 | 14 | 66003647 | *CTD-2014B16* | A | G | 0.29 | 1.01 (0.8-1.2) | 0.942 | non-coding intronic |
| rs17426432 | 15 | 75838049 | *UBE2Q2* | A | T | 0.28 | 0.98 (0.8-1.2) | 0.818 | 5'-45 Kb-DNM1P49/3'-5 Kb-UBE2Q2 |
| rs477421 | 15 | 101622450 | *TM2D3* | G | A | 0.15 | 0.96 (0.7-1.2) | 0.751 | non-coding intronic |
| **rs4984636** | 16 | 1202441 | *CACNA1H* | T | C | 0.134 | 0.7 (0.5-0.9) | 0.017 | **V651A/V664A** |
| rs9933126 | 16 | 20300870 | *GP2* | G | T | 0.372 | 0.93 (0.8-1.1) | 0.468 | 5'-62 Kb-SNRPEP3/3'-9 Kb- GP2 |
| rs7406402 | 17 | 31418963 | *RAB11FIP4* | A | G | 0.055 | 1.45 (1-2.1) | 0.053 | intronic |
| rs2025804 | 1 | 65480438 | *LEPR* | A | G | 0.442 | 1.02 (0.8-1.2) | 0.857 | intronic |
| rs5082 | 1 | 161223893 | *APOA2* | A | G | 0.149 | 0.83 (0.6-1.1) | 0.164 | 5'-0 Kb-APOA2/3'-2 Kb- TOMM40L |
| rs5400 | 3 | 171014511 | *SLC2A2* | G | A | 0.102 | 0.78 (0.6-1.1) | 0.127 | **T110I** |
| **rs1799883** | 4 | 119320747 | *FABP2* | C | T | 0.231 | 1.15 (0.9-1.4) | 0.197 | **T55A/T55S/T55P** |
| rs4994 | 8 | 37966280 | *ADRB3* | A | G | 0.249 | 1.15 (0.9-1.4) | 0.190 | **W64R** |
| rs1800588 | 15 | 58431476 | *LIPC* | T | C | 0.327 | 1.07 (0.8-1.1) | 0.439 | 5upstream, intronic |
| rs2287019 | 19 | 45698914 | *GIPR* | C | T | 0.063 | 0.97 (0.7-1.4) | 0.873 | 3downstream, intronic |
| rs1042714 | 5 | 148826910 | *ADRB2* | C | G | 0.138 | 0.79 (0.6-1) | 0.096 | **E27Q** |
| **rs1800795** | 7 | 22727026 | *IL6* | G | C | 0.113 | 0.66 (0.5-0.9) | 0.011 | 5upstream, intronic |
| rs4343 | 17 | 63488670 | *ACE* | A | G | 0.346 | 0.89 (0.7-1.1) | 0.254 | coding syn |
| rs4961 | 4 | 2904980 | *ADD1* | G | T | 0.211 | 0.99 (0.8-1.2) | 0.901 | **G460W** |
| **rs2070744** | 7 | 150992991 | *NOS3* | T | C | 0.154 | 0.72 (0.5-0.9) | 0.019 | intronic |
| rs11926707 | 3 | 46884049 | *KIF9* | C | T | 0.212 | 0.94 (0.7-1.2) | 0.597 | intronic |
| rs7978610 | 12 | 123984025 | *ZNF664* | G | C | 0.213 | 1.03 (0.8-1.3) | 0.829 | intronic |
| **rs1005752** | 15 | 77525786 | *HMG20A* | A | C | 0.388 | 1.21 (1-1.5) | 0.044 | regulatory_region_variant |
| rs2258238 | 12 | 65827280 | *HMGA2* | A | T | 0.072 | 0.79 (0.5-1.2) | 0.233 | regulatory_region_variant |
| rs3931020 | 1 | 74769633 | *TYW3/CRYZ* | C | T | 0.147 | 0.82 (0.6-1.1) | 0.166 | 5'-3 Kb-TYW3/3'-194 Kb- AC133865.1 |
| rs3745368 | 19 | 7670411 | *RETN* | G | A | 0.02 | 0.56 (0.3-1.2) | 0.150 | 3utr,3downstream |
| rs1862513 | 19 | 7668907 | *RETN* | C | G | 0.124 | 0.84 (0.6-1.1) | 0.250 | 5'-2 Kb-RPS27AP19/3'-0 Kb-RETN |
| **Selected from the OMIM database: https://www.omim.org/** | | | | | | | | | |
| **rs1801278** | 2 | 226795828 | *IRS1* | C | T | 0.034 | 0.55 (0.3-1) | 0.053 | **G972R** |
| rs1044498 | 6 | 131851228 | *ENPP1* | A | C | 0.191 | 0.99 (0.8-1.3) | 0.931 | **K121Q** |
| **rs1799999** | 7 | 113878379 | *PPP1R3A* | C | A | 0.263 | 1.26 (1-1.5) | 0.028 | **D905Y** |
| rs1805097 | 13 | 109782884 | IRS2 | T | C | 0.466 | 1.01 (0.8-1.2) | 0.952 | **G1057A** |
| **Selected from articles that included a Mexican population: see Table legend** | | | | | | | | | |
| **rs10261386^a^** | 7 | 120711855 | *CPED1* | T | C | 0.38 | 0.85 (1-1.4) | 0.103 | intronic |
| rs10823559^b^ | 10 | 72288302 | *PALD1* | C | C | 0.472 | 0.96 (0.8-1.2) | 0.650 | intronic |
| rs2975760^c^ | 2 | 240591746 | *CAPN10* | T | C | 0.064 | 0.75 (0.5-1.1) | 0.164 | intronic,3downstream |
| **rs7607759^C^** | 2 | 240591746 | *CAPN10* | A | G | 0.066 | 0.73 (0.5-1.1) | 0.131 | **T504A** |
| rs1800629^d^ | 6 | 31575254 | *TNF-𝛼* | G | A | 0.046 | 1.05 (0.7-1.6) | 0.820 | 5'-1 Kb-LTA/3'-0 Kb-TNF |
| rs361525^e^ | 6 | 31575324 | *TNF-𝛼* | G | A | 0.034 | 1.3 (0.8-2.1) | 0.281 | 5'-1 Kb-LTA/3'-0 Kb-TNF |
| rs2794521^f^ | 1 | 159715306 | *CRP* | T | C | 0.176 | 0.91 (0.7-1.2) | 0.437 | 5'-1 Kb-CRP/3'-44 Kb- AL445528.1 |
| rs909253^e^ | 6 | 31572536 | *LTA* | A | G | 0.366 | 0.96 (0.8-1.2) | 0.665 | non-coding intronic |
| rs1800972^g^ | 8 | 6877901 | *DEFB1* | G | C | 0.372 | 1.12 (0.9-1.4) | 0.232 | 5utr |
| **Selected from a previously published paper from our group: Sci Rep. 2019; 9(1): 2748** | | | | | | | | | |
| **rs149483638** | 11 | 2140300 | *IGF2* | C | T | 0.001 | 1.25 (1.1-1.4) | 0.0022 | Intron variant |
| **rs2237897** | 11 | 2837316 | *KCNQ1* | C | T | 0.051 | 1.44 (1.3-1.6) | 0.000000048 | Intron variant |
| **rs483353044** | 12 | 120999288 | *HNF1A* | G | A | 0 | 2 (0.8-5) | 1.3E-01 | Missense variant |
| **rs4458523** | 4 | 6288259 | *WFS1* | T | G | 0.385 | 1.07 (0.9-1.2) | 3.5E-01 | Intron variant |
| **rs3802177** | 8 | 117172786 | *SLC30A8* | G | A | 0.295 | 1.15 (1-1.3) | 0.044 | UTR variant |
| **rs4402960** | 3 | 185793899 | *IGF2BP2* | G | C | 0.315 | 1.28 (1.1-1.5) | 0.0031 | Intron variant |
| **rs7903146** | 10 | 112998590 | *TCF7L2* | C | G | [0.286](about:blank#frequency_tab) | 1.5 (1.3-1.8) | 0.0000080 | Intron variant |
| **rs10811661** | 9 | 22134095 | *CDKN2A* | T | A | [0.169](about:blank#frequency_tab) | 1.07 (0.7-1.5) | 0.731 | Single nucleotide variation |
| **rs75493593** | 17 | 7041768 | *SLC16A11* | G | C | 0.006 | 1.31 (1.2-1.5) | 0.000028 | Missense variant |

Abbreviations: Chr, chromosome; MAF, major allele frequency; MEX, Mexico; OR, odds ratio; SNP, single nuclear polymorphism; T2D, type 2 diabetes.

^a^Data Brief. 2020; 28: 104866; ^b^Nutrients. 2018; 10(9): 1175; ^c^Am J Hum Genet. 2004; 74(2): 208–222; ^d^PeerJ. 2016; 4: e2090; ^e^Int J Chron Obstruct Pulmon Dis. 2018; 13: 627–637; ^f^Int J Environ Res Public Health. 2016; 13(1): 103; ^g^Mol Genet Genomic Med. 2019; 7(1): e00509.

The SNP alleles in bold were associated with T2D in the whole sample or in the stratified sample by sex and age of T2D presentation.
**Table S2.** Groups of genes and their function related to T2D

| **Group** | **Locus** | **SNP** | **GENE NAME** | **Principle Function** | **Reference** |
| --- | --- | --- | --- | --- | --- |
| **Group G1: Genes involved in production of insulin located in chr 11p15.5** | | | | | |
| G1 | *INS-IGF2* | rs149483638 | Insulin-Like Growth Factor 2 Read-Through Product | Promotes the survival and proliferation of pancreatic β cells | Mercader et al., 2017 |
|  | *INS* | rs689 | Insulin | Insulin production | Johannessen et al., 2016 |
|  | *KCNQ1* | rs2237897 | Potassium Voltage-Gated Channel Subfamily Q Member 1 | A potassium channel involved in pancreatic β-cell mass | Zhang et al., 2015; Yu et al., 2015; Rattanatham et al., 2021 |
|  | *KCNQ1* | rs163168 | Potassium Voltage-Gated Channel Subfamily Q Member 1 | Pancreatic β-cell mass | Tan et al., 2009 |
|  | *SLC22A18* | rs450208 | Solute carrier family 22 member 18 | Glucose and lipid transport | Kitsiou-Tzeli et al., 2017 |
| **Group G2: Genes involved in production of insulin located in other genomic positions** | | | | | |
| G2 | *IGF2BP2* | rs4402960 | Insulin Like Growth Factor 2 MRNA Binding Protein 2 | Regulates translation of IGF2 gene involved in the survival and proliferation of pancreatic beta cells | Liu et al., 2020; Lasram et al., 2015 |
|  | *TCF7L2* | rs7903146 | Transcription Factor 7 Like 2 | Transcription factor involved in the production and secretion of insulin | Potasso et al., 2020; Barabash et al., 2020 |
|  | *CDKN2A/B* | rs10811661 | Cyclin Dependent Kinase Inhibitor 2B | Participates in mass regulation of the beta cells of the pancreas | Wang et al., 2020 |
|  | *SLC30A8* | rs3802177 | Solute carrier family 30 member 8 | Pancreatic beta cell-specific zinc transporter involved in insulin secretion | Ding et al., 2018; Fukunaka et al., 2018 |
|  | *HNF1A* | rs483353044 | HNF1 Homeobox A | Transcription factor involved in the production and secretion of insulin | Misra et al., 2020 |
|  | *WFS1* | rs4458523 | Wolframin ER Transmembrane Glycoprotein | Survival and function of pancreatic cells β | Langenberg et al., 2014 |
|  | *HMG20A* | rs1005752 | High Mobility Group 20A | Chromatin regulator involved in insulin secretion and maturation of pancreatic cells β | Huang et al., 2020; Mellado et al., 2018 |
| **Group G3: Genes involved in peripheral resistance to insulin** | | | | | |
| G3 | *SLC16A11* | rs75493593 | Solute Carrier Family 16 Member 11 | Lipid transport in the liver | Traurig et al., 2016; Hidalgo et al., 2019 |
|  | *IRS1* | rs1801278 | Insulin receptor substrate 1 | Insulin receptor signaling | Dabiri et al., 2019; Albegali et al., 2019 |
|  | *FABP2* | rs1799883 | Fatty acid binding protein 2 | Lipid transport in the intestine | Raza et al., 2017 |
|  | *CAPN10* | rs7607759 | Calpain 10 | Calcium-dependent cysteine protease, may play a role in insulin-stimulated glucose uptake | Song et al., 2004 |
|  | *PPP1R3A* | rs1799999 | Protein phosphatase 1 regulatory subunit 3A | Regulation of glycogen synthesis, encodes a regulatory subunit of a phosphatase [PP1] involved in glycogen synthesis | Sokhi et al., 2016 |
|  | *PTPRD* | rs10511567 | Protein tyrosine phosphatase, receptor type D | Regulation in insulin signaling | Parrillo et al., 2020 |
| **Group G4: Genes involved in inflammation and other functions** | | | | | |
| G4 | *IL6* | rs1800795 | Interleukin 6 | Inflammation and insulin resistance | Plataki et al., 2018; Ururahy et al., 2015 |
|  | *NOS3* | rs2070744 | Nitric oxide synthase 3 | Inflammation, vascular complications, and kidney disease | Abdullah et al., 2021 |
|  | *CPED1* | rs10261386 | Cadherin like and PC-esterase domain containing 1 | Associated with bone mineralization; peripheral vascular disease in T2D; WHR | Chesi et al., 2015 |
|  | *KHDRBS3* | rs6577691 | KH RNA binding domain containing, signal transduction associated 3 | Participates in the regulation of "splicing"; albuminuria and kidney disease in T2D | Gao et al., 2015 |
|  | *CACNA1H* | rs4984636 | Calcium voltage-gated channel subunit alpha1 H | Hyperglycemia, peripheral neuropathy, systolic pressure | Lehtinen et al., 2011 |

**Table S3.** Participant demographic and clinical characteristics (N = 2020)

| **Variable** | **Females** | | **Males** | | **Both sexes** | |
| --- | --- | --- | --- | --- | --- | --- |
|  | (n = 1089) | | (n = 931) | | (n = 2020) | |
|  | **Control** | **Cases** | **Control** | **Cases** | **Control** | **Cases** |
|  | (n = 543) | (n = 546) | (n = 465) | (n = 466) | (n = 1008) | (n = 1012) |
| Continuous variables: means ± SD (n) | | | | | | |
| Age (years) | 59.7 ± 11.1 (543) | 55.2 ± 12 (546) | 58.6 ± 11.4 (465) | 55.7 ± 11.4 (466) | 59.1 ± 11.3 (1008) | 55.5 ± 11.7 (1012) |
| BMIadj (kg/m^2^) | 28 ± 5 (543) | 31.4 ± 5.6 (546) | 26.9 ± 4.1 (465) | 30.5 ± 5 (466) | 27.5 ± 4.6 (1008) | 31 ± 5.3 (1012) |
| Waist (cm) | 93.4 ± 11.4 (357) | 97.5 ± 11.5 (426) | 93.4 ± 10.5 (310) | 98.6 ± 12.6 (281) | 93.4 ± 11 (667) | 97.9 ± 12 (707) |
| Hip (cm) | 103.8 ± 11.2 (336) | 106.5 ± 11.9 (424) | 98.9 ± 7.9 (298) | 101.7 ± 10.8 (277) | 101.5 ± 10.1 (634) | 104.6 ± 11.7 (701) |
| WHR | 0.9 ± 0.1 (336) | 0.92 ± 0.1 (424) | 0.94 ± 0.1 (298) | 0.97 ± 0.1 (277) | 0.92 ± 0.1 (634) | 0.94 ± 0.1 (701) |
| Age at diabetes diagnosis (years) |  | 45.6 ± 10.5 (546) |  | 46.1 ± 10.9 (466) |  | 45.8 ± 10.7 (1012) |
| Years with the disease |  | 9.5 ± 8.5 (546) |  | 9.6 ± 9 (466) |  | 9.5 ± 8.7 (1012) |
| Parental diabetes history: % (n) | | | | | | |
| None | 63.5 (273) | 37.6 (180) | 71.4 (260) | 41.7 (174) | 67.1 (533) | 39.5 (354) |
| Mother | 19.5 (84) | 30.1 (144) | 14.6 (53) | 26.4 (110) | 17.3 (137) | 28.3 (254) |
| Father | 11.4 (49) | 13.6 (65) | 9.6 (35) | 14.9 (62) | 10.6 (84) | 14.2 (127) |
| Both parents | 5.6 (24) | 18.8 (90) | 4.4 (16) | 17 (71) | 5 (40) | 18 (161) |
| Total | 100 (430) | 100 (479) | 100 (364) | 100 (417) | 100 (794) | 100 (896) |
| Smoking: % (n) | | | | | | |
| No | 87.9 (515) | 88.5 (45) | 71.4 (445) | 69.5 (57) | 80.2 (960) | 79.8 (102) |
| Yes | 12.1 (1) | 11.5 (479) | 28.6 (2) | 30.5 (388) | 19.8 (3) | 20.2 (867) |
| Total | 100 (519) | 100 (541) | 100 (448) | 100 (462) | 100 (967) | 100 (1003) |
| T2D treatment: % (n) | | | | | | |
| No |  | 8.6 (45) |  | 12.8 (57) |  | 10.5 (102) |
| Yes |  | 91.4 (479) |  | 87.2 (388) |  | 89.5 (867) |
| Total |  | 100 (524) |  | 100 (445) |  | 100 (969) |

Abbreviations: BMI, body mass index; SD, standard deviation.

BMIadj shows the BMI adjusted as described previously (reference Sci Rep. 019;9(1):2748); waist and hip circumferences shown are those measured at enrollment.

For the differences of means between groups, the P values were assessed using a t-test.

For the differences in the frequency distribution between groups, the P values were assessed using the chi-square test.

**Table S4.** Allelic frequency of 23 SNPs for the total population and stratified by age of T2D diagnosis (4040 chromosomes)

| **Group** | **Locus** | **RA** | **Allelic Frequency: % (n)** | | | | | | |
| --- | --- | --- | --- | --- | --- | --- | --- | --- | --- |
|  |  |  | **Control**  **(n = 2016)** | **All** | | **T2D diagnosis ≤ 45 years** | | **T2D diagnosis ≥ 46 years** | |
|  |  |  |  | **Cases**  **(n = 2014)** | **Chi square** | **Cases  (n = 1050)** | **Chi square** | **Cases**  **(n = 964)** | **Chi square** |
| **G1** | ***INS-IGF2* (rs149483638)** | 0 | 28.1 (563) | 23.8 (479) |  | 24.6 (255) |  | 23.2 (223) |  |
|  |  | 1 | 71.9 (1443) | 76.2 (1531) | 2.20E-03^a^ | 75.4 (783) | 3.90E-02 ^a^ | 76.8 (739) | 4.76E-03 ^a^ |
|  | ***INS* (rs689)** | 0 | 81.2 (1605) | 76.7 (1526) |  | 76.7 (779) |  | 76.7 (739) |  |
|  |  | 1 | 18.8 (371) | 23.3 (464) | 4.52E-04 ^a^ | 23.3 (237) | 3.39E-03 ^a^ | 23.3 (225) | 3.85E-03 ^a^ |
|  | ***KCNQ1* (rs2237897)** | 0 | 39.2 (787) | 31 (623) |  | 29 (301) |  | 33.4 (321) |  |
|  |  | 1 | 60.8 (1219) | 69 (1387) | 4.55E-08 ^a^ | 71 (737) | 2.33E-08 ^a^ | 66.6 (641) | 1.99E-03 ^a^ |
|  | ***CDKN1C* (rs163168)** | 0 | 43.6 (877) | 35.8 (723) |  | 33.3 (348) |  | 38.8 (373) |  |
|  |  | 1 | 56.4 (1133) | 64.2 (1295) | 4.17E-07 ^a^ | 66.7 (698) | 2.92E-08 ^a^ | 61.2 (589) | 1.21E-02 ^a^ |
|  | ***SLC22A18* (rs450208)** | 0 | 31.1 (625) | 24.7 (498) |  | 24.7 (259) |  | 24.7 (238) |  |
|  |  | 1 | 68.9 (1387) | 75.3 (1522) | 5.63E-06 ^a^ | 75.3 (789) | 2.35E-04 ^a^ | 75.3 (724) | 3.79E-04 ^a^ |
| **G2** | ***IGF2BP2* (rs4402960)** | 0 | 83.7 (1679) | 80.1 (1610) |  | 79.2 (822) |  | 81.1 (780) |  |
|  |  | 1 | 16.3 (327) | 19.9 (400) | 3.06E-03 ^a^ | 20.8 (216) | 2.07E-03 ^a^ | 18.9 (182) | 7.66E-02 ^a^ |
|  | ***TCF7L2* (rs7903146)** | 0 | 87.9 (1763) | 82.9 (1666) |  | 82.1 (852) |  | 83.8 (806) |  |
|  |  | 1 | 12.1 (243) | 17.1 (344) | 7.28E-06 ^a^ | 17.9 (186) | 1.28E-05 ^a^ | 16.2 (156) | 2.16E-03 ^a^ |
|  | ***CDKN2A* (rs10811661)** | 0 | 9.6 (193) | 8.7 (174) |  | 8.4 (87) |  | 9 (87) |  |
|  |  | 1 | 90.4 (1813) | 91.3 (1836) | 2.89E-01 | 91.6 (951) | 2.62E-01 | 91 (875) | 6.14E-01 |
|  | ***SLC30A8* (rs3802177)** | 0 | 72.8 (1460) | 69.9 (1405) |  | 66.3 (688) |  | 73.7 (709) |  |
|  |  | 1 | 27.2 (546) | 30.1 (605) | 4.35E-02 ^a^ | 33.7 (350) | 1.91E-04 ^a^ | 26.3 (253) | 5.97E-01 |
|  | ***HNF1A* (rs483353044** | 0 | 99.7 (1999) | 99.3 (1996) |  | 99.3 (1031) |  | 99.3 (955) |  |
|  |  | 1 | 0.3 (7) | 0.7 (14) | 1.27E-01 | 0.7 (7) | 2.08E-01 | 0.7 (7) | 1.59E-01 |
|  | ***WFS1* (rs4458523)** | 0 | 23.3 (468) | 22.1 (444) |  | 20.6 (214) |  | 23.5 (226) |  |
|  |  | 1 | 76.7 (1538) | 77.9 (1566) | 3.48E-01 | 79.4 (824) | 8.87E-02 ^a^ | 76.5 (736) | 9.22E-01 |
|  | ***HMG20A* (rs1005752)** | 0 | 58.1 (1168) | 59.5 (1200) |  | 57.9 (606) |  | 61.4 (589) |  |
|  |  | 1 | 41.9 (844) | 40.5 (816) | 3.43E-01 | 42.1 (440) | 9.51E-01 | 38.6 (371) | 8.68E-02 ^a^ |
| **G3** | ***SLC16A11* (rs75493593)** | 0 | 62.6 (1256) | 56.1 (1128) |  | 53.9 (560) |  | 58.4 (562) |  |
|  |  | 1 | 37.4 (750) | 43.9 (882) | 2.81E-05 ^a^ | 46.1 (478) | 3.87E-06 ^a^ | 41.6 (400) | 2.82E-02 ^a^ |
|  | ***IRS1* (rs1801278)** | 0 | 97.3 (1957) | 97.3 (1966) |  | 98 (1027) |  | 96.6 (929) |  |
|  |  | 1 | 2.7 (55) | 2.7 (54) | 9.06E-01 | 2 (21) | 2.18E-01 | 3.4 (33) | 2.94E-01 |
|  | ***FABP2* (rs1799883)** | 0 | 75.9 (1528) | 76.1 (1538) |  | 75.3 (789) |  | 77.2 (743) |  |
|  |  | 1 | 24.1 (484) | 23.9 (482) | 8.85E-01 | 24.7 (259) | 6.87E-01 | 22.8 (219) | 4.38E-01 |
|  | ***CAPN10* (rs7607759)** | 0 | 94.7 (1903) | 94.5 (1908) |  | 95.3 (999) |  | 93.7 (901) |  |
|  |  | 1 | 5.3 (107) | 5.5 (112) | 7.57E-01 | 4.7 (49) | 4.40E-01 | 6.3 (61) | 2.61E-01 |
|  | ***PPP1R3A* (rs1799999)** | 0 | 70.9 (1428) | 69.2 (1396) |  | 69.1 (723) |  | 69 (664) |  |
|  |  | 1 | 29.1 (586) | 30.8 (622) | 2.32E-01 | 30.9 (323) | 3.06E-01 | 31 (298) | 2.94E-01 |
|  | ***PTPRD* (rs10511567)** | 0 | 72.2 (1455) | 71.8 (1452) |  | 71.9 (755) |  | 71.7 (690) |  |
|  |  | 1 | 27.8 (559) | 28.2 (570) | 7.59E-01 | 28.1 (295) | 8.42E-01 | 28.3 (272) | 7.68E-01 |
| **G4** | ***IL6* (rs1800795)** | 0 | 92.1 (1855) | 92 (1859) |  | 92.9 (974) |  | 91 (875) |  |
|  |  | 1 | 7.9 (159) | 8 (161) | 9.29E-01 | 7.1 (74) | 4.09E-01 | 9 (87) | 2.87E-01 |
|  | ***NOS3* (rs2070744)** | 0 | 85.5 (1721) | 87.5 (1768) |  | 88.4 (926) |  | 86.6 (833) |  |
|  |  | 1 | 14.5 (293) | 12.5 (252) | 5.41E-02 ^a^ | 11.6 (122) | 2.58E-02 ^a^ | 13.4 (129) | 4.05E-01 |
|  | ***CPED1* (rs10261386)** | 0 | 65.3 (1314) | 64.4 (1301) |  | 65 (681) |  | 63.8 (614) |  |
|  |  | 1 | 34.7 (698) | 35.6 (719) | 5.49E-01 | 35 (367) | 8.57E-01 | 36.2 (348) | 4.28E-01 |
|  | ***KHDRBS3* (rs6577691)** | 0 | 87.7 (1764) | 89.5 (1807) |  | 89.7 (940) |  | 89.3 (859) |  |
|  |  | 1 | 12.3 (248) | 10.5 (213) | 7.55E-02 ^a^ | 10.3 (108) | 9.81E-02 ^a^ | 10.7 (103) | 2.00E-01 |
|  | ***CACNA1H* (rs4984636)** | 0 | 88.7 (1786) | 90.4 (1828) |  | 89.4 (939) |  | 91.5 (880) |  |
|  |  | 1 | 11.3 (228) | 9.6 (194) | 7.31E-02 ^a^ | 10.6 (111) | 5.30E-01 | 8.5 (82) | 1.95E-02 ^a^ |

Abbreviations: RA, risk allele; SNP, single nuclear polymorphism; T2D, type 2 diabetes.

^a^P < 0.1 (control versus case).

**Table S5.** Allelic frequency of 23 SNPs stratified by sex (4040 chromosomes)

| **Group** | **Locus** | **RA** | **Allelic Frequency** | | | | | |
| --- | --- | --- | --- | --- | --- | --- | --- | --- |
|  |  |  | **Females** | | | **Males** | | |
|  |  |  | **Controls**  **(n = 1086)** | **Cases**  **(n = 1092)** | **Chi square** | **Controls**  **(n = 930)** | **Cases**  **(n = 932)** | **Chi square** |
| **G1** | ***INS-IGF2* (rs149483638)** | 0 | 26.6 (288) | 25.1 (271) |  | 29.8 (275) | 22.4 (208) |  |
|  |  | 1 | 73.4 (794) | 74.9 (809) | 4.2E-01 | 70.2 (649) | 77.6 (722) | 2.9E-04 ^a^ |
|  | ***INS* (rs689)** | 0 | 80.2 (855) | 77.9 (829) |  | 82.4 (750) | 75.3 (697) |  |
|  |  | 1 | 19.8 (211) | 22.1 (235) | 1.9E-01 | 17.6 (160) | 24.7 (229) | 1.8E-04 ^a^ |
|  | ***KCNQ1* (rs2237897)** | 0 | 36 (389) | 31.9 (344) |  | 43.1 (398) | 30 (279) |  |
|  |  | 1 | 64 (693) | 68.1 (736) | 4.4E-02 ^a^ | 56.9 (526) | 70 (651) | 5.0E-09 ^a^ |
|  | ***CDKN1C* (rs163168)** | 0 | 41.4 (447) | 37 (403) |  | 46.2 (430) | 34.5 (320) |  |
|  |  | 1 | 58.6 (633) | 63 (687) | 3.5E-02 ^a^ | 53.8 (500) | 65.5 (608) | 2.4E-07 ^a^ |
|  | ***SLC22A18* (rs450208)** | 0 | 30.1 (326) | 23.1 (252) |  | 32.2 (299) | 26.5 (246) |  |
|  |  | 1 | 69.9 (756) | 76.9 (838) | 2.2E-04 ^a^ | 67.8 (631) | 73.5 (684) | 6.9E-03 ^a^ |
| **G2** | ***IGF2BP2* (rs4402960)** | 0 | 81.6 (883) | 79.8 (862) |  | 86.1 (796) | 80.4 (748) |  |
|  |  | 1 | 18.4 (199) | 20.2 (218) | 2.9E-01 | 13.9 (128) | 19.6 (182) | 9.7E-04 ^a^ |
|  | ***TCF7L2* (rs7903146)** | 0 | 85.9 (929) | 83.6 (903) |  | 90.3 (834) | 82 (763) |  |
|  |  | 1 | 14.1 (153) | 16.4 (177) | 1.5E-01 | 9.7 (90) | 18 (167) | 3.1E-07 ^a^ |
|  | ***CDKN2A* (rs10811661)** | 0 | 9.8 (106) | 8 (86) |  | 9.4 (87) | 9.5 (88) |  |
|  |  | 1 | 90.2 (976) | 92 (994) | 1.3E-01 | 90.6 (837) | 90.5 (842) | 9.7E-01 |
|  | ***SLC30A8* (rs3802177)** | 0 | 72.7 (787) | 70.4 (760) |  | 72.8 (673) | 69.4 (645) |  |
|  |  | 1 | 27.3 (295) | 29.6 (320) | 2.2E-01 | 27.2 (251) | 30.6 (285) | 9.8E-02 ^a^ |
|  | ***HNF1A* (rs483353044** | 0 | 99.7 (1079) | 99.4 (1073) |  | 99.6 (920) | 99.2 (923) |  |
|  |  | 1 | 0.3 (3) | 0.6 (7) | 2.0E-01 | 0.4 (4) | 0.8 (7) | 3.7E-01 |
|  | ***WFS1* (rs4458523)** | 0 | 23.5 (254) | 23 (248) |  | 23.2 (214) | 21.1 (196) |  |
|  |  | 1 | 76.5 (828) | 77 (832) | 7.8E-01 | 76.8 (710) | 78.9 (734) | 2.8E-01 |
|  | ***HMG20A* (rs1005752)** | 0 | 57.4 (621) | 59.2 (643) |  | 58.8 (547) | 59.9 (557) |  |
|  |  | 1 | 42.6 (461) | 40.8 (443) | 3.9E-01 | 41.2 (383) | 40.1 (373) | 6.4E-01 |
| **G3** | ***SLC16A11* (rs75493593)** | 0 | 62.5 (676) | 54.9 (593) |  | 62.8 (580) | 57.5 (535) |  |
|  |  | 1 | 37.5 (406) | 45.1 (487) | 3.5E-04 ^a^ | 37.2 (344) | 42.5 (395) | 2.1E-02 ^a^ |
|  | ***IRS1* (rs1801278)** | 0 | 97 (1050) | 97.4 (1062) |  | 97.5 (907) | 97.2 (904) |  |
|  |  | 1 | 3 (32) | 2.6 (28) | 5.8E-01 | 2.5 (23) | 2.8 (26) | 6.6E-01 |
|  | ***FABP2* (rs1799883)** | 0 | 76.1 (823) | 77.6 (846) |  | 75.8 (705) | 74.4 (692) |  |
|  |  | 1 | 23.9 (259) | 22.4 (244) | 3.9E-01 | 24.2 (225) | 25.6 (238) | 4.9E-01 |
|  | ***CAPN10* (rs7607759)** | 0 | 93.9 (1014) | 94 (1025) |  | 95.6 (889) | 94.9 (883) |  |
|  |  | 1 | 6.1 (66) | 6 (65) | 8.9E-01 | 4.4 (41) | 5.1 (47) | 5.1E-01 |
|  | ***PPP1R3A* (rs1799999)** | 0 | 72.5 (786) | 67.1 (730) |  | 69 (642) | 71.6 (666) |  |
|  |  | 1 | 27.5 (298) | 32.9 (358) | 6.0E-03 ^a^ | 31 (288) | 28.4 (264) | 2.2E-01 |
|  | ***PTPRD* (rs10511567)** | 0 | 74 (802) | 69.9 (762) |  | 70.2 (653) | 74 (690) |  |
|  |  | 1 | 26 (282) | 30.1 (328) | 3.4E-02 ^a^ | 29.8 (277) | 26 (242) | 6.6E-02 ^a^ |
| **G4** | ***IL6* (rs1800795)** | 0 | 91.2 (989) | 92.8 (1012) |  | 93.1 (866) | 91.1 (847) |  |
|  |  | 1 | 8.8 (95) | 7.2 (78) | 1.7E-01 | 6.9 (64) | 8.9 (83) | 1.0E-01 ^a^ |
|  | ***NOS3* (rs2070744)** | 0 | 84.8 (919) | 87.8 (957) |  | 86.2 (802) | 87.2 (811) |  |
|  |  | 1 | 15.2 (165) | 12.2 (133) | 4.1E-02 ^a^ | 13.8 (128) | 12.8 (119) | 5.4E-01 |
|  | ***CPED1* (rs10261386)** | 0 | 64.2 (695) | 65.5 (714) |  | 66.6 (619) | 63.1 (587) |  |
|  |  | 1 | 35.8 (387) | 34.5 (376) | 5.3E-01 | 33.4 (311) | 36.9 (343) | 1.2E-01 |
|  | ***KHDRBS3* (rs6577691)** | 0 | 88 (952) | 88.4 (964) |  | 87.3 (812) | 90.6 (843) |  |
|  |  | 1 | 12 (130) | 11.6 (126) | 7.4E-01 | 12.7 (118) | 9.4 (87) | 2.2E-02 ^a^ |
|  | ***CACNA1H* (rs4984636)** | 0 | 88.3 (957) | 91.3 (995) |  | 89.1 (829) | 89.4 (833) |  |
|  |  | 1 | 11.7 (127) | 8.7 (95) | 2.1E-02 ^a^ | 10.9 (101) | 10.6 (99) | 8.7E-01 |

Abbreviations: RA, risk allele; SNP, single nucleotide polymorphism; T2D, type 2 diabetes.

^a^P < 0.1 (control versus case).

**Table S6.** Allelic frequency of 23 SNPs stratified by sex and age of T2D diagnosis (4040 chromosomes)

| **Group** | **Locus** | **RA** | **Allelic Frequency** | | | | | | | | | |
| --- | --- | --- | --- | --- | --- | --- | --- | --- | --- | --- | --- | --- |
|  |  |  | **Controls** | | **Cases T2D diagnosis ≤45 years** | | | | **Cases T2D diagnosis ≥46 years** | | | |
|  |  |  | **Females**  **(n = 1086)** | **Males**  **(n = 930)** | **Females**  **(n = 576)** | **Chi square** | **Males**  **(n = 474)** | **Chi square** | **Females**  **(n = 510)** | **Chi square** | **Males**  **(n = 454)** | **Chi square** |
| **G1** | ***INS-IGF2* (rs149483638)** | 0 | 26.6 (288) | 29.8 (275) | 25.4 (144) |  | 23.5 (111) |  | 24.8 (126) |  | 21.4 (97) |  |
|  |  | 1 | 73.4 (794) | 70.2 (649) | 74.6 (422) | 6.1E-01 | 76.5 (361) | 1.4E-02 ^a^ | 75.2 (382) | 4.4E-01 | 78.6 (357) | 9.7E-04 ^a^ |
|  | ***INS* (rs689)** | 0 | 80.2 (855) | 82.4 (750) | 78.8 (432) |  | 74.1 (347) |  | 76.7 (391) |  | 76.7 (348) |  |
|  |  | 1 | 19.8 (211) | 17.6 (160) | 21.2 (116) | 5.2E-01 | 25.9 (121) | 3.1E-04 ^a^ | 23.3 (119) | 1.1E-01 ^a^ | 23.3 (106) | 1.1E-02 ^a^ |
|  | ***KCNQ1* (rs2237897)** | 0 | 36 (389) | 43.1 (398) | 30 (170) |  | 27.8 (131) |  | 34.1 (173) |  | 32.6 (148) |  |
|  |  | 1 | 64 (693) | 56.9 (526) | 70 (396) | 1.6E-02 ^a^ | 72.2 (341) | 2.4E-08 ^a^ | 65.9 (335) | 4.6E-01 | 67.4 (306) | 1.9E-04 ^a^ |
|  | ***CDKN1C* (rs163168)** | 0 | 41.4 (447) | 46.2 (430) | 34.2 (197) |  | 32.1 (151) |  | 40.2 (204) |  | 37.2 (169) |  |
|  |  | 1 | 58.6 (633) | 53.8 (500) | 65.8 (379) | 4.3E-03 ^a^ | 67.9 (319) | 4.2E-07 ^a^ | 59.8 (304) | 6.4E-01 | 62.8 (285) | 1.5E-03 ^a^ |
|  | ***SLC22A18* (rs450208)** | 0 | 30.1 (326) | 32.2 (299) | 22.7 (131) |  | 27.1 (128) |  | 23.8 (121) |  | 25.8 (117) |  |
|  |  | 1 | 69.9 (756) | 67.8 (631) | 77.3 (445) | 1.4E-03 ^a^ | 72.9 (344) | 5.3E-02 ^a^ | 76.2 (387) | 9.1E-03 ^a^ | 74.2 (337) | 1.5E-02 ^a^ |
| **G2** | ***IGF2BP2* (rs4402960)** | 0 | 81.6 (883) | 86.1 (796) | 79.3 (449) |  | 79 (373) |  | 80.3 (408) |  | 81.9 (372) |  |
|  |  | 1 | 18.4 (199) | 13.9 (128) | 20.7 (117) | 2.6E-01 | 21 (99) | 6.5E-04 ^a^ | 19.7 (100) | 5.4E-01 | 18.1 (82) | 4.1E-02 ^a^ |
|  | ***TCF7L2* (rs7903146)** | 0 | 85.9 (929) | 90.3 (834) | 83.2 (471) |  | 80.7 (381) |  | 84.1 (427) |  | 83.5 (379) |  |
|  |  | 1 | 14.1 (153) | 9.7 (90) | 16.8 (95) | 1.5E-01 | 19.3 (91) | 5.2E-07 ^a^ | 15.9 (81) | 3.4E-01 | 16.5 (75) | 2.7E-04 ^a^ |
|  | ***CDKN2A* (rs10811661)** | 0 | 9.8 (106) | 9.4 (87) | 6.9 (39) |  | 10.2 (48) |  | 9.3 (47) |  | 8.8 (40) |  |
|  |  | 1 | 90.2 (976) | 90.6 (837) | 93.1 (527) | 4.8E-02 ^a^ | 89.8 (424) | 6.5E-01 | 90.7 (461) | 7.3E-01 | 91.2 (414) | 7.2E-01 |
|  | ***SLC30A8* (rs3802177)** | 0 | 72.7 (787) | 72.8 (673) | 67.1 (380) |  | 65.3 (308) |  | 73.8 (375) |  | 73.6 (334) |  |
|  |  | 1 | 27.3 (295) | 27.2 (251) | 32.9 (186) | 1.8E-02 ^a^ | 34.7 (164) | 3.4E-03 ^a^ | 26.2 (133) | 6.5E-01 | 26.4 (120) | 7.7E-01 |
|  | ***HNF1A* (rs483353044** | 0 | 99.7 (1079) | 99.6 (920) | 99.5 (563) |  | 99.2 (468) |  | 99.2 (504) |  | 99.3 (451) |  |
|  |  | 1 | 0.3 (3) | 0.4 (4) | 0.5 (3) | 4.2E-01 | 0.8 (4) | 3.3E-01 | 0.8 (4) | 1.5E-01 | 0.7 (3) | 5.8E-01 |
|  | ***WFS1* (rs4458523)** | 0 | 23.5 (254) | 23.2 (214) | 22.1 (125) |  | 18.9 (89) |  | 23.6 (120) |  | 23.3 (106) |  |
|  |  | 1 | 76.5 (828) | 76.8 (710) | 77.9 (441) | 5.2E-01 | 81.1 (383) | 6.5E-02 ^a^ | 76.4 (388) | 9.5E-01 | 76.7 (348) | 9.4E-01 |
|  | ***HMG20A* (rs1005752)** | 0 | 57.4 (621) | 58.8 (547) | 58 (333) |  | 57.8 (273) |  | 60.9 (308) |  | 61.9 (281) |  |
|  |  | 1 | 42.6 (461) | 41.2 (383) | 42 (241) | 8.1E-01 | 42.2 (199) | 7.3E-01 | 39.1 (198) | 1.9E-01 | 38.1 (173) | 2.7E-01 |
| **G3** | ***SLC16A11* (rs75493593)** | 0 | 62.5 (676) | 62.8 (580) | 52.5 (297) |  | 55.7 (263) |  | 57.3 (291) |  | 59.7 (271) |  |
|  |  | 1 | 37.5 (406) | 37.2 (344) | 47.5 (269) | 8.8E-05 ^a^ | 44.3 (209) | 1.1E-02 ^a^ | 42.7 (217) | 4.8E-02 ^a^ | 40.3 (183) | 2.7E-01 |
|  | ***IRS1* (rs1801278)** | 0 | 97 (1050) | 97.5 (907) | 98.3 (566) |  | 97.7 (461) |  | 96.5 (490) |  | 96.7 (439) |  |
|  |  | 1 | 3 (32) | 2.5 (23) | 1.7 (10) | 1.3E-01 | 2.3 (11) | 8.7E-01 | 3.5 (18) | 5.3E-01 | 3.3 (15) | 3.7E-01 |
|  | ***FABP2* (rs1799883)** | 0 | 76.1 (823) | 75.8 (705) | 78.1 (450) |  | 71.8 (339) |  | 77.4 (393) |  | 77.1 (350) |  |
|  |  | 1 | 23.9 (259) | 24.2 (225) | 21.9 (126) | 3.4E-01 | 28.2 (133) | 1.1E-01 ^a^ | 22.6 (115) | 5.7E-01 | 22.9 (104) | 6.0E-01 |
|  | ***CAPN10* (rs7607759)** | 0 | 93.9 (1014) | 95.6 (889) | 95.3 (549) |  | 95.3 (450) |  | 92.9 (472) |  | 94.5 (429) |  |
|  |  | 1 | 6.1 (66) | 4.4 (41) | 4.7 (27) | 2.3E-01 | 4.7 (22) | 8.3E-01 | 7.1 (36) | 4.6E-01 | 5.5 (25) | 3.7E-01 |
|  | ***PPP1R3A* (rs1799999)** | 0 | 72.5 (786) | 69 (642) | 66 (379) |  | 72.9 (344) |  | 67.9 (345) |  | 70.3 (319) |  |
|  |  | 1 | 27.5 (298) | 31 (288) | 34 (195) | 6.0E-03 ^a^ | 27.1 (128) | 1.4E-01 | 32.1 (163) | 5.9E-02 ^a^ | 29.7 (135) | 6.4E-01 |
|  | ***PTPRD* (rs10511567)** | 0 | 74 (802) | 70.2 (653) | 68.9 (397) |  | 75.5 (358) |  | 71.1 (361) |  | 72.5 (329) |  |
|  |  | 1 | 26 (282) | 29.8 (277) | 31.1 (179) | 2.8E-02 ^a^ | 24.5 (116) | 3.6E-02 ^a^ | 28.9 (147) | 2.2E-01 | 27.5 (125) | 3.9E-01 |
| **G4** | ***IL6* (rs1800795)** | 0 | 91.2 (989) | 93.1 (866) | 92.9 (535) |  | 93 (439) |  | 92.7 (471) |  | 89 (404) |  |
|  |  | 1 | 8.8 (95) | 6.9 (64) | 7.1 (41) | 2.4E-01 | 7 (33) | 9.4E-01 | 7.3 (37) | 3.2E-01 | 11 (50) | 8.7E-03 ^a^ |
|  | ***NOS3* (rs2070744)** | 0 | 84.8 (919) | 86.2 (802) | 88.7 (511) |  | 87.9 (415) |  | 86.8 (441) |  | 86.3 (392) |  |
|  |  | 1 | 15.2 (165) | 13.8 (128) | 11.3 (65) | 2.7E-02 ^a^ | 12.1 (57) | 3.8E-01 | 13.2 (67) | 2.8E-01 | 13.7 (62) | 9.6E-01 |
|  | ***CPED1* (rs10261386)** | 0 | 64.2 (695) | 66.6 (619) | 65.5 (377) |  | 64.4 (304) |  | 65.6 (333) |  | 61.9 (281) |  |
|  |  | 1 | 35.8 (387) | 33.4 (311) | 34.5 (199) | 6.2E-01 | 35.6 (168) | 4.2E-01 | 34.4 (175) | 6.1E-01 | 38.1 (173) | 8.8E-02 ^a^ |
|  | ***KHDRBS3* (rs6577691)** | 0 | 88 (952) | 87.3 (812) | 89.1 (513) |  | 90.5 (427) |  | 87.8 (446) |  | 91 (413) |  |
|  |  | 1 | 12 (130) | 12.7 (118) | 10.9 (63) | 5.1E-01 | 9.5 (45) | 8.2E-02 ^a^ | 12.2 (62) | 9.1E-01 | 9 (41) | 4.5E-02 ^a^ |
|  | ***CACNA1H* (rs4984636)** | 0 | 88.3 (957) | 89.1 (829) | 89.4 (515) |  | 89.5 (424) |  | 93.3 (474) |  | 89.4 (406) |  |
|  |  | 1 | 11.7 (127) | 10.9 (101) | 10.6 (61) | 4.9E-01 | 10.5 (50) | 8.6E-01 | 6.7 (34) | 1.9E-03 ^a^ | 10.6 (48) | 8.7E-01 |

Abbreviations: RA, risk allele; SNP, single nucleotide polymorphism; T2D, type 2 diabetes.

^a^P < 0.1 (control versus case).

**Table S7.** Allelic association of 23 SNPs with T2D in the whole sample and stratified by sex (4040 chromosomes)

| **Group** | **Locus (SNP)** | **Univariate Logistic Regression Models** | | | | | | | | | |
| --- | --- | --- | --- | --- | --- | --- | --- | --- | --- | --- | --- |
|  |  | **T2D diagnosis ≤45 years** | | | **T2D diagnosis ≥46 years** | | | **Females** | | **Males** | |
|  |  | **OR (95% CI)** | **p-Wald** | **SI** | **OR (95% CI)** | **p-Wald** | **SI** | **OR (95% CI)** | **p-Wald** | **OR (95% CI)** | **p-Wald** |
| **G1** | INS-IGF2 (rs149483638) | 1.2 (1-1.4) | 3.9E-02 ^a^ | >0.05 | 1.29 (1.1-1.5) | 4.8E-03 ^a^ | 5.7E-02 ^a^ | 1.08 (0.9-1.3) | 4.2E-01 | 1.47 (1.2-1.8) | 3.0E-04 ^a^ |
|  | INS (rs689) | 1.32 (1.1-1.6) | 3.4E-03 ^a^ | 3.1E-02 ^a^ | 1.32 (1.1-1.6) | 3.9E-03 ^a^ | >0.05 | 1.15 (0.9-1.4) | 1.9E-01 | 1.54 (1.2-1.9) | 1.9E-04 ^a^ |
|  | KCNQ1 (rs2237897) | 1.58 (1.3-1.9) | 2.6E-08 ^a^ | 1.3E-02 ^a^ | 1.29 (1.1-1.5) | 2.0E-03 ^a^ | 2.7E-0 2 ^a^ | 1.2 (1-1.4) | 4.4E-02 ^a^ | 1.77 (1.5-2.1) | 5.8E-09 ^a^ |
|  | CDKN1C (rs163168) | 1.55 (1.3-1.8) | 3.2E-08 ^a^ | 6.9E-02 ^a^ | 1.22 (1-1.4) | 1.2E-02 ^a^ | 4.6E-02 ^a^ | 1.2 (1-1.4) | 3.5E-02 ^a^ | 1.63 (1.4-2) | 2.6E-07 ^a^ |
|  | SLC22A18 (rs450208) | 1.37 (1.2-1.6) | 2.4E-04 ^a^ | >0.05 | 1.37 (1.2-1.6) | 3.9E-04 ^a^ | >0.05 | 1.43 (1.2-1.7) | 2.3E-04 ^a^ | 1.32 (1.1-1.6) | 7.0E-03 ^a^ |
| **G2** | IGF2BP2 (rs4402960) | 1.35 (1.1-1.6) | 2.1E-03 ^a^ | 7.1E-02 ^a^ | 1.2 (1-1.5) | 7.7E-02 ^a^ | >0.05 | 1.12 (0.9-1.4) | 2.9E-01 | 1.51 (1.2-1.9) | 1.0E-03 ^a^ |
|  | TCF7L2 (rs7903146) | 1.58 (1.3-1.9) | 1.4E-05 ^a^ | 5.9E-03 ^a^ | 1.4 (1.1-1.7) | 2.2E-03 ^a^ | 3.9E-02 ^a^ | 1.19 (0.9-1.5) | 1.5E-01 | 2.03 (1.5-2.7) | 4.4E-07 ^a^ |
|  | CDKN2A (rs10811661) | 1.16 (0.9-1.5) | 2.6E-01 | 8.4E-02 ^a^ | 1.07 (0.8-1.4) | 6.1E-01 | >0.05 | 1.26 (0.9-1.7) | 1.3E-01 | 0.99 (0.7-1.4) | 9.7E-01 |
|  | SLC30A8 (rs3802177) | 1.36 (1.2-1.6) | 2.0E-04 ^a^ | >0.05 | 0.95 (0.8-1.1) | 6.0E-01 | >0.05 | 1.12 (0.9-1.4) | 2.2E-01 | 1.18 (1-1.4) | 9.9E-02 ^a^ |
|  | HNF1A (rs483353044) | 1.94 (0.7-5.5) | 2.2E-01 | >0.05 | 2.09 (0.7-6) | 1.7E-01 | >0.05 | 2.35 (0.6-9.1) | 2.2E-01 | 1.74 (0.5-6) | 3.8E-01 |
|  | WFS1 (rs4458523) | 1.17 (1-1.4) | 8.9E-02 ^a^ | >0.05 | 0.99 (0.8-1.2) | 9.2E-01 | >0.05 | 1.03 (0.8-1.3) | 7.8E-01 | 1.13 (0.9-1.4) | 2.8E-01 |
|  | HMG20A (rs1005752) | 1 (0.9-1.2) | 9.5E-01 | >0.05 | 0.87 (0.7-1) | 8.7E-02 ^a^ | >0.05 | 0.93 (0.8-1.1) | 3.9E-01 | 0.96 (0.8-1.2) | 6.4E-01 |
| **G3** | SLC16A11 (rs75493593) | 1.43 (1.2-1.7) | 4.0E-06 ^a^ | >0.05 | 1.19 (1-1.4) | 2.8E-02 ^a^ | >0.05 | 1.37 (1.2-1.6) | 3.6E-04 ^a^ | 1.24 (1-1.5) | 2.1E-02 ^a^ |
|  | IRS1 (rs1801278) | 0.73 (0.4-1.2) | 2.2E-01 | >0.05 | 1.26 (0.8-2) | 3.0E-01 | >0.05 | 0.87 (0.5-1.4) | 5.8E-01 | 1.13 (0.6-2) | 6.6E-01 |
|  | FABP2 (rs1799883) | 1.04 (0.9-1.2) | 6.9E-01 | 6.9E-02 ^a^ | 0.93 (0.8-1.1) | 4.4E-01 | >0.05 | 0.92 (0.8-1.1) | 3.9E-01 | 1.08 (0.9-1.3) | 4.9E-01 |
|  | CAPN10 (rs7607759) | 0.87 (0.6-1.2) | 4.4E-01 | >0.05 | 1.2 (0.9-1.7) | 2.6E-01 | >0.05 | 0.97 (0.7-1.4) | 8.9E-01 | 1.15 (0.8-1.8) | 5.1E-01 |
|  | PPP1R3A (rs1799999) | 1.09 (0.9-1.3) | 3.1E-01 | 3.3E-03 ^a^ | 1.09 (0.9-1.3) | 2.9E-01 | >0.05 | 1.29 (1.1-1.6) | 6.1E-03 ^a^ | 0.88 (0.7-1.1) | 2.2E-01 |
|  | PTPRD (rs10511567) | 1.02 (0.9-1.2) | 8.4E-01 | 2.5E-03 ^a^ | 1.03 (0.9-1.2) | 7.7E-01 | >0.05 | 1.22 (1-1.5) | 3.5E-02 ^a^ | 0.83 (0.7-1) | 6.6E-02 ^a^ |
| **G4** | IL6 (rs1800795) | 0.89 (0.7-1.2) | 4.1E-01 | >0.05 | 1.16 (0.9-1.5) | 2.9E-01 | 1.1E-02 ^a^ | 0.8 (0.6-1.1) | 1.7E-01 | 1.33 (0.9-1.9) | 1.0E-01 ^a^ |
|  | NOS3 (rs2070744) | 0.77 (0.6-1) | 2.6E-02 ^a^ | >0.05 | 0.91 (0.7-1.1) | 4.1E-01 | >0.05 | 0.77 (0.6-1) | 4.1E-02 ^a^ | 0.92 (0.7-1.2) | 5.4E-01 |
|  | CPED1 (rs10261386) | 1.01 (0.9-1.2) | 8.6E-01 | >0.05 | 1.07 (0.9-1.3) | 4.3E-01 | >0.05 | 0.95 (0.8-1.1) | 5.3E-01 | 1.16 (1-1.4) | 1.2E-01 |
|  | KHDRBS3 (rs6577691) | 0.82 (0.6-1) | 9.8E-02 ^a^ | >0.05 | 0.85 (0.7-1.1) | 2.0E-01 | >0.05 | 0.96 (0.7-1.2) | 7.4E-01 | 0.71 (0.5-1) | 2.2E-02 ^a^ |
|  | CACNA1H (rs4984636) | 0.93 (0.7-1.2) | 5.3E-01 | >0.05 | 0.73 (0.6-1) | 2.0E-02 ^a^ | 3.2E-02 ^a^ | 0.72 (0.5-1) | 2.1E-02 ^a^ | 0.98 (0.7-1.3) | 8.7E-01 |
| **No. SNPs** | | **12** | |  | **10** | |  | **8** | | **12** | |
| **Sum of univariate R^2^** | | **0.0737** | |  | **0.0332** | |  | **0.035** | | **0.107** | |

Abbreviations: CI, confidence interval; OR, odds ratio; SI, sex interaction; SNP, single polymorphonuclear polymorphism; T2D, type 2 diabetes.

^a^P < 0.1.

**Table S8.** Analysis of Hardy–Weinberg equilibrium in the control population (n = 1008)

| **Locus** | **Number of RA** | **Genotypic Frequency: % (n)** | | |
| --- | --- | --- | --- | --- |
|  |  | **Observed** | **Expected** | **Chi square** |
| **SLC16A11 (rs75493593)** | 0 | 40.9 (410) | 39.2 (393) | 0.270 |
|  | 1 | 43.5 (436) | 46.8 (470) |  |
|  | 2 | 15.7 (157) | 14 (140) |  |
| **INS-IGF2 (rs149483638)** | 0 | 10.4 (104) | 7.9 (79) | 0.026 |
|  | 1 | 35.4 (355) | 40.4 (405) |  |
|  | 2 | 54.2 (544) | 51.7 (519) |  |
| **HNF1A (rs483353044** | 0 | 99.3 (996) | 99.3 (996) | 1.000 |
|  | 1 | 0.7 (7) | 0.7 (7) |  |
| **WFS1 (rs4458523)** | 0 | 6.3 (63) | 5.4 (55) | 0.604 |
|  | 1 | 34.1 (342) | 35.8 (359) |  |
|  | 2 | 59.6 (598) | 58.8 (590) |  |
| **IGF2BP2 (rs4402960)** | 0 | 70.1 (703) | 70.1 (703) | 0.998 |
|  | 1 | 27.2 (273) | 27.3 (274) |  |
|  | 2 | 2.7 (27) | 2.7 (27) |  |
| **INS (rs689)** | 0 | 66.9 (661) | 66 (652) | 0.440 |
|  | 1 | 28.6 (283) | 30.5 (301) |  |
|  | 2 | 4.5 (44) | 3.5 (35) |  |
| **KCNQ1 (rs2237897)** | 0 | 16.8 (169) | 15.4 (154) | 0.386 |
|  | 1 | 44.8 (449) | 47.7 (478) |  |
|  | 2 | 38.4 (385) | 36.9 (370) |  |
| **TCF7L2 (rs7903146)** | 0 | 77.7 (779) | 77.2 (775) | 0.714 |
|  | 1 | 20.4 (205) | 21.3 (214) |  |
|  | 2 | 1.9 (19) | 1.5 (15) |  |
| **CDKN2A (rs10811661)** | 0 | 1.5 (15) | 0.9 (9) | 0.391 |
|  | 1 | 16.3 (163) | 17.4 (174) |  |
|  | 2 | 82.3 (825) | 81.7 (819) |  |
| **IRS1 (rs1801278)** | 0 | 94.8 (954) | 94.6 (952) | 0.560 |
|  | 1 | 4.9 (49) | 5.3 (53) |  |
|  | 2 | 0.3 (3) | 0.1 (1) |  |
| **FABP2 (rs1799883)** | 0 | 58.3 (587) | 57.7 (580) | 0.701 |
|  | 1 | 35.2 (354) | 36.5 (368) |  |
|  | 2 | 6.5 (65) | 5.8 (58) |  |
| **CAPN10 (rs7607759)** | 0 | 89.4 (898) | 89.6 (901) | 0.204 |
|  | 1 | 10.6 (107) | 10.1 (101) |  |
|  | 2 | 0 (0) | 0.3 (3) |  |
| **PPP1R3A (rs1799999)** | 0 | 50.7 (511) | 50.3 (506) | 0.876 |
|  | 1 | 40.3 (406) | 41.3 (415) |  |
|  | 2 | 8.9 (90) | 8.5 (85) |  |
| **SLC30A8 (rs3802177)** | 0 | 53 (532) | 53 (531) | 0.991 |
|  | 1 | 39.5 (396) | 39.6 (397) |  |
|  | 2 | 7.5 (75) | 7.4 (74) |  |
| **PTPRD (rs10511567)** | 0 | 53.6 (540) | 52.2 (526) | 0.299 |
|  | 1 | 37.2 (375) | 40.1 (404) |  |
|  | 2 | 9.1 (92) | 7.7 (78) |  |
| **CDKN1C (rs163168)** | 0 | 20.2 (203) | 19 (191) | 0.567 |
|  | 1 | 46.9 (471) | 49.2 (494) |  |
|  | 2 | 32.9 (331) | 31.8 (319) |  |
| **SLC22A18 (rs450208)** | 0 | 10.3 (104) | 9.6 (97) | 0.769 |
|  | 1 | 41.5 (417) | 42.8 (431) |  |
|  | 2 | 48.2 (485) | 47.5 (478) |  |
| **IL6 (rs1800795)** | 0 | 85.7 (863) | 84.8 (854) | 0.084 |
|  | 1 | 12.8 (129) | 14.5 (146) |  |
|  | 2 | 1.5 (15) | 0.6 (6) |  |
| **HMG20A (rs1005752)** | 0 | 34.3 (345) | 33.7 (339) | 0.860 |
|  | 1 | 47.5 (478) | 48.7 (490) |  |
|  | 2 | 18.2 (183) | 17.6 (177) |  |
| **NOS3 (rs2070744)** | 0 | 74 (745) | 73 (735) | 0.254 |
|  | 1 | 22.9 (231) | 24.9 (250) |  |
|  | 2 | 3.1 (31) | 2.1 (21) |  |
| **CPED1 (rs10261386)** | 0 | 43.3 (436) | 42.7 (429) | 0.790 |
|  | 1 | 43.9 (442) | 45.3 (456) |  |
|  | 2 | 12.7 (128) | 12 (121) |  |
| **KHDRBS3 (rs6577691)** | 0 | 77.5 (780) | 76.9 (773) | 0.415 |
|  | 1 | 20.3 (204) | 21.6 (217) |  |
|  | 2 | 2.2 (22) | 1.5 (15) |  |
| **CACNA1H (rs4984636)** | 0 | 78.6 (792) | 78.6 (792) | 1.000 |
|  | 1 | 20.1 (202) | 20.1 (202) |  |
|  | 2 | 1.3 (13) | 1.3 (13) |  |

Abbreviation: RA, risk allele.

**Table S9.** Genotypic frequency of 23 SNPs (N = 2020)

| **Group** | **Locus** | **RA** | **Genotypic Frequency: % (n)** | | | | | | | | | | | |
| --- | --- | --- | --- | --- | --- | --- | --- | --- | --- | --- | --- | --- | --- | --- |
|  |  |  | **T2D diagnosis any age** | | | | **T2D diagnosis ≤ 45 years** | | | | **T2D diagnosis ≥ 46 years** | | | |
|  |  |  | **Controls** | **Cases** | **Chi square** | **Linear Chi square** | **Controls** | **Cases** | **Chi square** | **Linear Chi square** | **Controls** | **Cases** | **Chi square** | **Linear Chi square** |
| **G1** | **INS-IGF2 (rs149483638)** | 0 | 10.4 (104) | 5.9 (59) |  |  | 10.4 (104) | 6.6 (34) |  |  | 10.4 (104) | 5.2 (25) |  |  |
|  |  | 1 | 35.4 (355) | 35.9 (361) | 9.30E-04 ^a^ | 3.12E-03 ^a^ | 35.4 (355) | 36 (187) | 4.59E-02 ^a^ | 4.85E-02 ^a^ | 35.4 (355) | 36 (173) | 3.58E-03 | 6.78E-03 |
|  |  | 2 | 54.2 (544) | 58.2 (585) |  |  | 54.2 (544) | 57.4 (298) |  |  | 54.2 (544) | 58.8 (283) |  |  |
|  | **INS (rs689)** | 0 | 66.9 (661) | 58.9 (586) |  |  | 66.9 (661) | 58.9 (299) |  |  | 66.9 (661) | 58.9 (284) |  |  |
|  |  | 1 | 28.6 (283) | 35.6 (354) | 1.10E-03 ^a^ | 5.63E-04 ^a^ | 28.6 (283) | 35.6 (181) | 8.90E-03 ^a^ | 4.12E-03 ^a^ | 28.6 (283) | 35.5 (171) | 1.11E-02 | 4.70E-03 |
|  |  | 2 | 4.5 (44) | 5.5 (55) |  |  | 4.5 (44) | 5.5 (28) |  |  | 4.5 (44) | 5.6 (27) |  |  |
|  | **KCNQ1 (rs2237897)** | 0 | 16.8 (169) | 11.1 (112) |  |  | 16.8 (169) | 9.1 (47) |  |  | 16.8 (169) | 13.5 (65) |  |  |
|  |  | 1 | 44.8 (449) | 39.7 (399) | 8.21E-07 ^a^ | 1.31E-07 ^a^ | 44.8 (449) | 39.9 (207) | 4.14E-07 ^a^ | 6.00E-08 ^a^ | 44.8 (449) | 39.7 (191) | 7.47E-03 | 2.92E-03 |
|  |  | 2 | 38.4 (385) | 49.2 (494) |  |  | 38.4 (385) | 51.1 (265) |  |  | 38.4 (385) | 46.8 (225) |  |  |
|  | **CDKN1C (rs163168)** | 0 | 20.2 (203) | 13.4 (135) |  |  | 20.2 (203) | 11.1 (58) |  |  | 20.2 (203) | 16 (77) |  |  |
|  |  | 1 | 46.9 (471) | 44.9 (453) | 4.13E-06 ^a^ | 7.16E-07 ^a^ | 46.9 (471) | 44.4 (232) | 3.46E-07 ^a^ | 5.61E-08 ^a^ | 46.9 (471) | 45.5 (219) | 4.93E-02 | 1.42E-02 |
|  |  | 2 | 32.9 (331) | 41.7 (421) |  |  | 32.9 (331) | 44.6 (233) |  |  | 32.9 (331) | 38.5 (185) |  |  |
|  | **SLC22A18 (rs450208)** | 0 | 10.3 (104) | 5.5 (56) |  |  | 10.3 (104) | 5 (26) |  |  | 10.3 (104) | 6.2 (30) |  |  |
|  |  | 1 | 41.5 (417) | 38.2 (386) | 1.56E-05 ^a^ | 6.22E-06 ^a^ | 41.5 (417) | 39.5 (207) | 3.93E-04 ^a^ | 2.49E-04 ^a^ | 41.5 (417) | 37 (178) | 2.09E-03 | 4.59E-04 |
|  |  | 2 | 48.2 (485) | 56.2 (568) |  |  | 48.2 (485) | 55.5 (291) |  |  | 48.2 (485) | 56.8 (273) |  |  |
| **G2** | **IGF2BP2 (rs4402960)** | 0 | 70.1 (703) | 64.5 (648) |  |  | 70.1 (703) | 63 (327) |  |  | 70.1 (703) | 66.1 (318) |  |  |
|  |  | 1 | 27.2 (273) | 31.2 (314) | 1.25E-02 ^a^ | 3.28E-03 ^a^ | 27.2 (273) | 32.4 (168) | 8.74E-03 ^a^ | 2.20E-03 ^a^ | 27.2 (273) | 29.9 (144) | 1.96E-01 | 7.83E-02 |
|  |  | 2 | 2.7 (27) | 4.3 (43) |  |  | 2.7 (27) | 4.6 (24) |  |  | 2.7 (27) | 4 (19) |  |  |
|  | **TCF7L2 (rs7903146)** | 0 | 77.7 (779) | 68.1 (684) |  |  | 77.7 (779) | 66.5 (345) |  |  | 77.7 (779) | 69.9 (336) |  |  |
|  |  | 1 | 20.4 (205) | 29.7 (298) | 6.99E-06 ^a^ | 7.05E-06 ^a^ | 20.4 (205) | 31.2 (162) | 1.26E-05 ^a^ | 1.34E-05 ^a^ | 20.4 (205) | 27.9 (134) | 4.69E-03 | 2.38E-03 |
|  |  | 2 | 1.9 (19) | 2.3 (23) |  |  | 1.9 (19) | 2.3 (12) |  |  | 1.9 (19) | 2.3 (11) |  |  |
|  | **CDKN2A (rs10811661)** | 0 | 1.5 (15) | 1.1 (11) |  |  | 1.5 (15) | 1.3 (7) |  |  | 1.5 (15) | 0.8 (4) |  |  |
|  |  | 1 | 16.3 (163) | 15.1 (152) | 5.57E-01 | 3.02E-01 | 16.3 (163) | 14.1 (73) | 5.15E-01 | 2.79E-01 | 16.3 (163) | 16.4 (79) | 5.67E-01 | 6.22E-01 |
|  |  | 2 | 82.3 (825) | 83.8 (842) |  |  | 82.3 (825) | 84.6 (439) |  |  | 82.3 (825) | 82.7 (398) |  |  |
|  | **SLC30A8 (rs3802177)** | 0 | 53 (532) | 50.4 (507) |  |  | 53 (532) | 47.8 (248) |  |  | 53 (532) | 53.2 (256) |  |  |
|  |  | 1 | 39.5 (396) | 38.9 (391) | 4.38E-02 ^a^ | 4.80E-02 ^a^ | 39.5 (396) | 37 (192) | 1.20E-05 ^a^ | 3.12E-04 ^a^ | 39.5 (396) | 41 (197) | 4.81E-01 | 5.95E-01 |
|  |  | 2 | 7.5 (75) | 10.6 (107) |  |  | 7.5 (75) | 15.2 (79) |  |  | 7.5 (75) | 5.8 (28) |  |  |
|  | **HNF1A (rs483353044** | 0 | 99.3 (996) | 98.6 (991) |  |  | 99.3 (996) | 98.7 (512) |  |  | 99.3 (996) | 98.5 (474) |  |  |
|  |  | 1 | 0.7 (7) | 1.4 (14) | 1.26E-01 |  | 0.7 (7) | 1.3 (7) | 2.07E-01 |  | 0.7 (7) | 1.5 (7) | 1.58E-01 |  |
|  | **WFS1 (rs4458523)** | 0 | 6.3 (63) | 5.5 (55) |  |  | 6.3 (63) | 4.2 (22) |  |  | 6.3 (63) | 6.7 (32) |  |  |
|  |  | 1 | 34.1 (342) | 33.2 (334) | 6.37E-01 | 3.58E-01 | 34.1 (342) | 32.8 (170) | 1.84E-01 | 9.40E-02 ^a^ | 34.1 (342) | 33.7 (162) | 9.57E-01 | 9.24E-01 |
|  |  | 2 | 59.6 (598) | 61.3 (616) |  |  | 59.6 (598) | 63 (327) |  |  | 59.6 (598) | 59.7 (287) |  |  |
|  | **HMG20A (rs1005752)** | 0 | 34.3 (345) | 35.9 (362) |  |  | 34.3 (345) | 35 (183) |  |  | 34.3 (345) | 36.9 (177) |  |  |
|  |  | 1 | 47.5 (478) | 47.2 (476) | 6.41E-01 | 3.48E-01 | 47.5 (478) | 45.9 (240) | 8.18E-01 | 9.51E-01 | 47.5 (478) | 49 (235) | 1.44E-01 | 8.81E-02 |
|  |  | 2 | 18.2 (183) | 16.9 (170) |  |  | 18.2 (183) | 19.1 (100) |  |  | 18.2 (183) | 14.2 (68) |  |  |
| **G3** | **SLC16A11 (rs75493593)** | 0 | 40.9 (410) | 32 (322) |  |  | 40.9 (410) | 30.6 (159) |  |  | 40.9 (410) | 33.5 (161) |  |  |
|  |  | 1 | 43.5 (436) | 48.2 (484) | 1.21E-04 ^a^ | 4.39E-05 ^a^ | 43.5 (436) | 46.6 (242) | 4.87E-05 ^a^ | 8.45E-06 ^a^ | 43.5 (436) | 49.9 (240) | 2.05E-02 | 3.15E-02 |
|  |  | 2 | 15.7 (157) | 19.8 (199) |  |  | 15.7 (157) | 22.7 (118) |  |  | 15.7 (157) | 16.6 (80) |  |  |
|  | **IRS1 (rs1801278)** | 0 | 94.8 (954) | 94.9 (958) |  |  | 94.8 (954) | 96.2 (504) |  |  | 94.8 (954) | 93.3 (449) |  |  |
|  |  | 1 | 4.9 (49) | 5 (50) | 9.00E-01 | 9.09E-01 | 4.9 (49) | 3.6 (19) | 4.92E-01 | 2.37E-01 | 4.9 (49) | 6.4 (31) | 4.33E-01 | 3.09E-01 |
|  |  | 2 | 0.3 (3) | 0.2 (2) |  |  | 0.3 (3) | 0.2 (1) |  |  | 0.3 (3) | 0.2 (1) |  |  |
|  | **FABP2 (rs1799883)** | 0 | 58.3 (587) | 58 (586) |  |  | 58.3 (587) | 57.4 (301) |  |  | 58.3 (587) | 58.8 (283) |  |  |
|  |  | 1 | 35.2 (354) | 36.2 (366) | 7.44E-01 | 8.86E-01 | 35.2 (354) | 35.7 (187) | 9.23E-01 | 6.93E-01 | 35.2 (354) | 36.8 (177) | 2.57E-01 | 4.41E-01 |
|  |  | 2 | 6.5 (65) | 5.7 (58) |  |  | 6.5 (65) | 6.9 (36) |  |  | 6.5 (65) | 4.4 (21) |  |  |
|  | **CAPN10 (rs7607759)** | 0 | 89.4 (898) | 89.2 (901) |  |  | 89.4 (898) | 91 (477) |  |  | 89.4 (898) | 87.5 (421) |  |  |
|  |  | 1 | 10.6 (107) | 10.5 (106) | 2.23E-01 | 7.53E-01 | 10.6 (107) | 8.6 (45) | 6.68E-02 ^a^ | 4.34E-01 | 10.6 (107) | 12.3 (59) | 2.26E-01 | 2.50E-01 |
|  |  | 2 | 0 (0) | 0.3 (3) |  |  | 0 (0) | 0.4 (2) |  |  | 0 (0) | 0.2 (1) |  |  |
|  | **PPP1R3A (rs1799999)** | 0 | 50.7 (511) | 48.1 (485) |  |  | 50.7 (511) | 48.6 (254) |  |  | 50.7 (511) | 47.2 (227) |  |  |
|  |  | 1 | 40.3 (406) | 42.2 (426) | 4.73E-01 | 2.35E-01 | 40.3 (406) | 41.1 (215) | 5.82E-01 | 3.13E-01 | 40.3 (406) | 43.7 (210) | 4.23E-01 | 2.96E-01 |
|  |  | 2 | 8.9 (90) | 9.7 (98) |  |  | 8.9 (90) | 10.3 (54) |  |  | 8.9 (90) | 9.1 (44) |  |  |
|  | **PTPRD (rs10511567)** | 0 | 53.6 (540) | 51.5 (521) |  |  | 53.6 (540) | 51 (268) |  |  | 53.6 (540) | 52 (250) |  |  |
|  |  | 1 | 37.2 (375) | 40.6 (410) | 2.55E-01 | 7.63E-01 | 37.2 (375) | 41.7 (219) | 1.59E-01 | 8.45E-01 | 37.2 (375) | 39.5 (190) | 6.90E-01 | 7.74E-01 |
|  |  | 2 | 9.1 (92) | 7.9 (80) |  |  | 9.1 (92) | 7.2 (38) |  |  | 9.1 (92) | 8.5 (41) |  |  |
| **G4** | **IL6 (rs1800795)** | 0 | 85.7 (863) | 84.8 (856) |  |  | 85.7 (863) | 86.3 (452) |  |  | 85.7 (863) | 83 (399) |  |  |
|  |  | 1 | 12.8 (129) | 14.6 (147) | 1.28E-01 | 9.31E-01 | 12.8 (129) | 13.4 (70) | 1.42E-01 | 4.26E-01 | 12.8 (129) | 16 (77) | 2.03E-01 | 3.07E-01 |
|  |  | 2 | 1.5 (15) | 0.7 (7) |  |  | 1.5 (15) | 0.4 (2) |  |  | 1.5 (15) | 1 (5) |  |  |
|  | **NOS3 (rs2070744)** | 0 | 74 (745) | 77.4 (782) |  |  | 74 (745) | 79 (414) |  |  | 74 (745) | 75.7 (364) |  |  |
|  |  | 1 | 22.9 (231) | 20.2 (204) | 1.77E-01 | 6.36E-02 ^a^ | 22.9 (231) | 18.7 (98) | 9.11E-02 ^a^ | 3.22E-02 ^a^ | 22.9 (231) | 21.8 (105) | 7.09E-01 | 4.21E-01 |
|  |  | 2 | 3.1 (31) | 2.4 (24) |  |  | 3.1 (31) | 2.3 (12) |  |  | 3.1 (31) | 2.5 (12) |  |  |
|  | **CPED1 (rs10261386)** | 0 | 43.3 (436) | 41.6 (420) |  |  | 43.3 (436) | 41.4 (217) |  |  | 43.3 (436) | 42 (202) |  |  |
|  |  | 1 | 43.9 (442) | 45.6 (461) | 7.07E-01 | 5.52E-01 | 43.9 (442) | 47.1 (247) | 4.63E-01 | 8.57E-01 | 43.9 (442) | 43.7 (210) | 6.74E-01 | 4.37E-01 |
|  |  | 2 | 12.7 (128) | 12.8 (129) |  |  | 12.7 (128) | 11.5 (60) |  |  | 12.7 (128) | 14.3 (69) |  |  |
|  | **KHDRBS3 (rs6577691)** | 0 | 77.5 (780) | 80.6 (814) |  |  | 77.5 (780) | 81.5 (427) |  |  | 77.5 (780) | 79.8 (384) |  |  |
|  |  | 1 | 20.3 (204) | 17.7 (179) | 2.24E-01 | 8.46E-02 ^a^ | 20.3 (204) | 16.4 (86) | 1.82E-01 | 1.11E-01 | 20.3 (204) | 18.9 (91) | 3.59E-01 | 2.11E-01 |
|  |  | 2 | 2.2 (22) | 1.7 (17) |  |  | 2.2 (22) | 2.1 (11) |  |  | 2.2 (22) | 1.2 (6) |  |  |
|  | **CACNA1H (rs4984636)** | 0 | 78.6 (792) | 81.6 (825) |  |  | 78.6 (792) | 80 (420) |  |  | 78.6 (792) | 83.4 (401) |  |  |
|  |  | 1 | 20.1 (202) | 17.6 (178) | 1.85E-01 | 7.24E-02 ^a^ | 20.1 (202) | 18.9 (99) | 8.21E-01 | 5.31E-01 | 20.1 (202) | 16.2 (78) | 5.15E-02 | 1.90E-02 |
|  |  | 2 | 1.3 (13) | 0.8 (8) |  |  | 1.3 (13) | 1.1 (6) |  |  | 1.3 (13) | 0.4 (2) |  |  |

Abbreviations: RA, risk allele; SNP, single nucleotide polymorphism; T2D, type 2 diabetes.

^a^Markers that entered the multivariate model.

**Table S10.** Genotypic frequency of 23 SNPs stratified by sex (N = 2020)

| **Group** | **Locus** | **RA** | **Genotypic frequency** | | | | | | | |
| --- | --- | --- | --- | --- | --- | --- | --- | --- | --- | --- |
|  |  |  | **Females** | | | | **Males** | | | |
|  |  |  | **Controls  (n = 543)** | **Cases  (n = 543)** | **Chi square** | **Linear chi square** | **Controls  (n = 465)** | **Cases  (n = 464)** | **Chi square** | **Linear chi square** |
| **G1** | **INS-IGF2 (rs149483638)** | 0 | 9.4 (51) | 7 (38) |  |  | 11.5 (53) | 4.5 (21) |  |  |
|  |  | 1 | 34.4 (186) | 36.1 (195) | 3.46E-01 | 4.36E-01 | 36.6 (169) | 35.7 (166) | 2.43E-04 ^a^ | 4.34E-04 ^a^ |
|  |  | 2 | 56.2 (304) | 56.9 (307) |  |  | 51.9 (240) | 59.8 (278) |  |  |
|  | **INS (rs689)** | 0 | 65.5 (349) | 61.5 (327) |  |  | 68.6 (312) | 55.9 (259) |  |  |
|  |  | 1 | 29.5 (157) | 32.9 (175) | 3.97E-01 | 2.06E-01 | 27.7 (126) | 38.7 (179) | 4.13E-04 ^a^ | 1.87E-04 ^a^ |
|  |  | 2 | 5.1 (27) | 5.6 (30) |  |  | 3.7 (17) | 5.4 (25) |  |  |
|  | **KCNQ1 (rs2237897)** | 0 | 13.7 (74) | 10.9 (59) |  |  | 20.6 (95) | 11.4 (53) |  | |
|  |  | 1 | 44.5 (241) | 41.9 (226) | 1.41E-01 ^b^ | 4.80E-02 ^a^ | 45 (208) | 37.2 (173) | 1.67E-07 ^a^ | 3.08E-08 ^a^ |
|  |  | 2 | 41.8 (226) | 47.2 (255) |  |  | 34.4 (159) | 51.4 (239) |  |  |
|  | **CDKN1C (rs163168)** | 0 | 17.4 (94) | 13.4 (73) |  |  | 23.4 (109) | 13.4 (62) |  |  |
|  |  | 1 | 48 (259) | 47.2 (257) | 1.01E-01 ^a^ | 3.54E-02 ^a^ | 45.6 (212) | 42.2 (196) | 4.72E-06 ^a^ | 7.50E-07 ^a^ |
|  |  | 2 | 34.6 (187) | 39.4 (215) |  |  | 31 (144) | 44.4 (206) |  |  |
|  | **SLC22A18 (rs450208)** | 0 | 10.4 (56) | 4.4 (24) |  |  | 10.3 (48) | 6.9 (32) |  |  |
|  |  | 1 | 39.6 (214) | 37.4 (204) | 2.46E-04 ^a^ | 2.44E-04 ^a^ | 43.7 (203) | 39.1 (182) | 2.61E-02 ^a^ | 6.99E-03 ^a^ |
|  |  | 2 | 50.1 (271) | 58.2 (317) |  |  | 46 (214) | 54 (251) |  |  |
| **G2** | **IGF2BP2 (rs4402960)** | 0 | 66.4 (359) | 64.1 (346) |  |  | 74.5 (344) | 64.9 (302) |  |  |
|  |  | 1 | 30.5 (165) | 31.5 (170) | 4.70E-01 | 2.92E-01 | 23.4 (108) | 31 (144) | 4.85E-03 ^a^ | 1.12E-03 ^a^ |
|  |  | 2 | 3.1 (17) | 4.4 (24) |  |  | 2.2 (10) | 4.1 (19) |  |  |
|  | **TCF7L2 (rs7903146)** | 0 | 74.1 (401) | 69.6 (376) |  |  | 81.8 (378) | 66.2 (308) |  |  |
|  |  | 1 | 23.5 (127) | 28 (151) | 2.37E-01 | 1.47E-01 | 16.9 (78) | 31.6 (147) | 4.36E-07 ^a^ | 2.48E-07 ^a^ |
|  |  | 2 | 2.4 (13) | 2.4 (13) |  |  | 1.3 (6) | 2.2 (10) |  |  |
|  | **CDKN2A (rs10811661)** | 0 | 1.8 (10) | 0.9 (5) |  |  | 1.1 (5) | 1.3 (6) |  |  |
|  |  | 1 | 15.9 (86) | 14.1 (76) | 2.87E-01 | 1.48E-01 | 16.7 (77) | 16.3 (76) | 9.51E-01 | 9.73E-01 |
|  |  | 2 | 82.3 (445) | 85 (459) |  |  | 82.3 (380) | 82.4 (383) |  |  |
|  | **SLC30A8 (rs3802177)** | 0 | 54 (292) | 50.4 (272) |  |  | 51.9 (240) | 50.5 (235) |  |  |
|  |  | 1 | 37.5 (203) | 40 (216) | 4.77E-01 | 2.34E-01 | 41.8 (193) | 37.6 (175) | 1.13E-02 ^a^ | 1.04E-01 ^a^ |
|  |  | 2 | 8.5 (46) | 9.6 (52) |  |  | 6.3 (29) | 11.8 (55) |  |  |
|  | **HNF1A (rs483353044)** | 0 | 99.4 (538) | 98.7 (533) |  |  | 99.1 (458) | 98.5 (458) |  |  |
|  |  | 1 | 0.6 (3) | 1.3 (7) | 2.03E-01 ^b^ | 2.03E-01 ^b^ | 0.9 (4) | 1.5 (7) | 3.69E-01 | 3.69E-01 |
|  | **WFS1 (rs4458523)** | 0 | 6.3 (34) | 6.1 (33) |  |  | 6.3 (29) | 4.7 (22) |  |  |
|  |  | 1 | 34.4 (186) | 33.7 (182) | 9.60E-01 | 7.83E-01 | 33.8 (156) | 32.7 (152) | 5.10E-01 | 2.88E-01 |
|  |  | 2 | 59.3 (321) | 60.2 (325) |  |  | 60 (277) | 62.6 (291) |  |  |
|  | **HMG20A (rs1005752)** | 0 | 34.4 (186) | 35.4 (192) |  |  | 34.2 (159) | 36.6 (170) |  |  |
|  |  | 1 | 46 (249) | 47.7 (259) | 5.28E-01 | 4.00E-01 | 49.2 (229) | 46.7 (217) | 7.06E-01 | 6.38E-01 |
|  |  | 2 | 19.6 (106) | 16.9 (92) |  |  | 16.6 (77) | 16.8 (78) |  |  |
| **G3** | **SLC16A11 (rs75493593)** | 0 | 41.2 (223) | 30.2 (163) |  |  | 40.5 (187) | 34.2 (159) |  |  |
|  |  | 1 | 42.5 (230) | 49.4 (267) | 7.02E-04 ^a^ | 4.95E-04 ^a^ | 44.6 (206) | 46.7 (217) | 7.91E-02 ^a^ | 2.44E-02 ^a^ |
|  |  | 2 | 16.3 (88) | 20.4 (110) |  |  | 14.9 (69) | 19.1 (89) |  |  |
|  | **IRS1 (rs1801278)** | 0 | 94.5 (511) | 94.9 (517) |  |  | 95.3 (443) | 94.8 (441) |  |  |
|  |  | 1 | 5.2 (28) | 5.1 (28) | 3.64E-01 | 5.88E-01 | 4.5 (21) | 4.7 (22) | 8.35E-01 | 6.79E-01 |
|  |  | 2 | 0.4 (2) | 0 (0) |  |  | 0.2 (1) | 0.4 (2) |  |  |
|  | **FABP2 (rs1799883)** | 0 | 58.2 (315) | 60 (327) |  |  | 58.5 (272) | 55.7 (259) |  |  |
|  |  | 1 | 35.7 (193) | 35.2 (192) | 5.94E-01 | 3.92E-01 | 34.6 (161) | 37.4 (174) | 6.63E-01 | 4.94E-01 |
|  |  | 2 | 6.1 (33) | 4.8 (26) |  |  | 6.9 (32) | 6.9 (32) |  |  |
|  | **CAPN10 (rs7607759)** | 0 | 87.8 (474) | 88.4 (482) |  |  | 91.2 (424) | 90.1 (419) |  |  |
|  |  | 1 | 12.2 (66) | 11.2 (61) | 3.26E-01 | 8.83E-01 | 8.8 (41) | 9.7 (45) | 5.45E-01 | 5.07E-01 |
|  |  | 2 | 0 (0) | 0.4 (2) |  |  | 0 (0) | 0.2 (1) |  |  |
|  | **PPP1R3A (rs1799999)** | 0 | 53.3 (289) | 46.5 (253) |  |  | 47.7 (222) | 49.9 (232) |  |  |
|  |  | 1 | 38.4 (208) | 41.2 (224) | 2.60E-02 ^a^ | 7.54E-03 ^a^ | 42.6 (198) | 43.4 (202) | 2.42E-01 | 2.16E-01 |
|  |  | 2 | 8.3 (45) | 12.3 (67) |  |  | 9.7 (45) | 6.7 (31) |  |  |
|  | **PTPRD (rs10511567)** | 0 | 57 (309) | 48.1 (262) |  |  | 49.7 (231) | 55.6 (259) |  |  |
|  |  | 1 | 33.9 (184) | 43.7 (238) | 4.21E-03 ^a^ | 3.80E-02 ^a^ | 41.1 (191) | 36.9 (172) | 1.81E-01 ^b^ | 7.04E-02 ^a^ |
|  |  | 2 | 9 (49) | 8.3 (45) |  |  | 9.2 (43) | 7.5 (35) |  |  |
| **G4** | **IRS1 (rs1801278)** | 0 | 94.5 (511) | 94.9 (517) |  |  | 95.3 (443) | 94.8 (441) |  |  |
|  |  | 1 | 5.2 (28) | 5.1 (28) | 3.64E-01 | 5.88E-01 | 4.5 (21) | 4.7 (22) | 8.35E-01 | 6.79E-01 |
|  |  | 2 | 0.4 (2) | 0 (0) |  |  | 0.2 (1) | 0.4 (2) |  |  |
|  | **FABP2 (rs1799883)** | 0 | 58.2 (315) | 60 (327) |  |  | 58.5 (272) | 55.7 (259) |  |  |
|  |  | 1 | 35.7 (193) | 35.2 (192) | 5.94E-01 | 3.92E-01 | 34.6 (161) | 37.4 (174) | 6.63E-01 | 4.94E-01 |
|  |  | 2 | 6.1 (33) | 4.8 (26) |  |  | 6.9 (32) | 6.9 (32) |  |  |
|  | **CAPN10 (rs7607759)** | 0 | 87.8 (474) | 88.4 (482) |  |  | 91.2 (424) | 90.1 (419) |  |  |
|  |  | 1 | 12.2 (66) | 11.2 (61) | 3.26E-01 | 8.83E-01 | 8.8 (41) | 9.7 (45) | 5.45E-01 | 5.07E-01 |
|  |  | 2 | 0 (0) | 0.4 (2) |  |  | 0 (0) | 0.2 (1) |  |  |
|  | **PPP1R3A (rs1799999)** | 0 | 53.3 (289) | 46.5 (253) |  |  | 47.7 (222) | 49.9 (232) |  |  |
|  |  | 1 | 38.4 (208) | 41.2 (224) | 2.60E-02 ^a^ | 7.54E-03 ^a^ | 42.6 (198) | 43.4 (202) | 2.42E-01 | 2.16E-01 |
|  |  | 2 | 8.3 (45) | 12.3 (67) |  |  | 9.7 (45) | 6.7 (31) |  |  |
|  | **PTPRD (rs10511567)** | 0 | 57 (309) | 48.1 (262) |  |  | 49.7 (231) | 55.6 (259) |  |  |
|  |  | 1 | 33.9 (184) | 43.7 (238) | 4.21E-03 ^a^ | 3.80E-02 ^a^ | 41.1 (191) | 36.9 (172) | 1.81E-01 ^b^ | 7.04E-02 ^a^ |
|  |  | 2 | 9 (49) | 8.3 (45) |  |  | 9.2 (43) | 7.5 (35) |  |  |
|  | **IL6 (rs1800795)** | 0 | 84.1 (456) | 86.4 (471) |  |  | 87.5 (407) | 82.8 (385) |  |  |
|  |  | 1 | 14.2 (77) | 12.8 (70) | 2.88E-01 | 1.82E-01 | 11.2 (52) | 16.6 (77) | 3.96E-02 ^a^ | 1.11E-01 ^b^ |
|  |  | 2 | 1.7 (9) | 0.7 (4) |  |  | 1.3 (6) | 0.6 (3) |  |  |
|  | **NOS3 (rs2070744)** | 0 | 73.1 (396) | 78.3 (427) |  |  | 75.1 (349) | 76.3 (355) |  |  |
|  |  | 1 | 23.4 (127) | 18.9 (103) | 1.27E-01 ^b^ | 5.17E-02 ^a^ | 22.4 (104) | 21.7 (101) | 7.70E-01 | 5.47E-01 |
|  |  | 2 | 3.5 (19) | 2.8 (15) |  |  | 2.6 (12) | 1.9 (9) |  |  |
|  | **CPED1 (rs10261386)** | 0 | 41.4 (224) | 43.3 (236) |  |  | 45.6 (212) | 39.6 (184) |  |  |
|  |  | 1 | 45.7 (247) | 44.4 (242) | 8.13E-01 | 5.37E-01 | 41.9 (195) | 47.1 (219) | 1.73E-01 ^b^ | 1.25E-01 ^b^ |
|  |  | 2 | 12.9 (70) | 12.3 (67) |  |  | 12.5 (58) | 13.3 (62) |  |  |
|  | **KHDRBS3 (rs6577691)** | 0 | 78.6 (425) | 79.1 (431) |  |  | 76.3 (355) | 82.4 (383) |  |  |
|  |  | 1 | 18.9 (102) | 18.7 (102) | 9.13E-01 | 7.54E-01 | 21.9 (102) | 16.6 (77) | 7.26E-02 ^a^ | 2.30E-02 ^a^ |
|  |  | 2 | 2.6 (14) | 2.2 (12) |  |  | 1.7 (8) | 1.1 (5) |  |  |
|  | **CACNA1H (rs4984636)** | 0 | 77.9 (422) | 82.9 (452) |  |  | 79.6 (370) | 80 (373) |  |  |
|  |  | 1 | 20.8 (113) | 16.7 (91) | 4.57E-02 ^a^ | 1.95E-02 ^a^ | 19.1 (89) | 18.7 (87) | 9.83E-01 | 8.69E-01 |
|  |  | 2 | 1.3 (7) | 0.4 (2) |  |  | 1.3 (6) | 1.3 (6) |  |  |

Abbreviations: RA, risk allele; SNP, single nucleotide polymorphism; T2D, type 2 diabetes.

^a^P < 0.1 and ^b^P < 0.2 were selected for the multivariate model.

**Table S11.** Genotypic frequency of 23 SNPs stratified by sex and age of T2D diagnosis (N = 2020)

| **Group** | **Locus** | **RA** | **Genotypic frequency** | | | | | | | | | | | | | | | | | |
| --- | --- | --- | --- | --- | --- | --- | --- | --- | --- | --- | --- | --- | --- | --- | --- | --- | --- | --- | --- | --- |
|  |  |  | **Controls** | | **T2D diagnosis ≤45 years** | | | | | | | | **T2D diagnosis ≥ 46 years** | | | | | | | |
|  |  |  | **Females** | **Males** | **Females** | | | | **Males** | | | | **Females** | | | | **Males** | | | |
|  |  |  | **Control  (n = 543)** | **Control  (n = 465)** | **Cases  (n = 288)** | **Chi square** | **Linear chi square** | **Chi square alleles comparison** | **Cases  (n = 237)** | **Chi square** | **Linear chi square** | **Chi square alleles comparison** | **Cases  (n = 255)** | **Chi square** | **Linear chi square** | **Chi square alleles comparison** | **Cases  (n = 227)** | **Chi square** | **Linear chi square** | **Chi square alleles comparison** |
| **G1** | **INS-IGF2 (rs149483638)** | 0 | 9.4 (51) | 11.5 (53) | 7.4 (21) |  |  |  | 5.5 (13) |  |  |  | 6.7 (17) |  |  |  | 3.5 (8) |  |  |  |
|  |  | 1 | 34.4 (186) | 36.6 (169) | 36 (102) | 6.04E-01 | 6.23E-01 | 6.1E-01 | 36 (85) | 2.85E-02 ^a^ | 1.82E-02 ^a^ | 1.4E-02 ^a^ | 36.2 (92) | 4.27E-01 | 4.62E-01 | 4.4E-01 | 35.7 (81) | 1.46E-03 ^a^ | 1.50E-03 ^a^ | 9.7E-04 ^a^ |
|  |  | 2 | 56.2 (304) | 51.9 (240) | 56.5 (160) |  |  |  | 58.5 (138) |  |  |  | 57.1 (145) |  |  |  | 60.8 (138) |  |  |  |
|  | **INS (rs689)** | 0 | 65.5 (349) | 68.6 (312) | 62 (170) |  |  |  | 55.1 (129) |  |  |  | 60.4 (154) |  |  |  | 57.3 (130) |  |  |  |
|  |  | 1 | 29.5 (157) | 27.7 (126) | 33.6 (92) | 4.71E-01 | 5.25E-01 | 5.2E-01 | 38 (89) | 1.69E-03 ^a^ | 4.02E-04 ^a^ | 3.1E-04 ^a^ | 32.5 (83) | 2.95E-01 | 1.20E-01 ^b^ | 1.1E-01 ^b^ | 38.8 (88) | 1.15E-02 ^a^ | 1.14E-02 ^a^ | 1.1E-02 ^a^ |
|  |  | 2 | 5.1 (27) | 3.7 (17) | 4.4 (12) |  |  |  | 6.8 (16) |  |  |  | 7.1 (18) |  |  |  | 4 (9) |  |  |  |
|  | **KCNQ1 (rs2237897)** | 0 | 13.7 (74) | 20.6 (95) | 9.9 (28) |  |  |  | 8.1 (19) |  |  |  | 12.2 (31) |  |  |  | 15 (34) |  |  |  |
|  |  | 1 | 44.5 (241) | 45 (208) | 40.3 (114) | 5.97E-02 ^a^ | 1.82E-02 ^a^ | 1.6E-02 ^a^ | 39.4 (93) | 5.65E-07 ^a^ | 8.44E-08 ^a^ | 2.4E-08 ^a^ | 43.7 (111) | 7.67E-01 | 4.68E-01 | 4.6E-01 | 35.2 (80) | 5.21E-04 ^a^ | 4.35E-04 ^a^ | 1.9E-04 ^a^ |
|  |  | 2 | 41.8 (226) | 34.4 (159) | 49.8 (141) |  |  |  | 52.5 (124) |  |  |  | 44.1 (112) |  |  |  | 49.8 (113) |  |  |  |
|  | **CDKN1C (rs163168)** | 0 | 17.4 (94) | 23.4 (109) | 11.8 (34) |  |  |  | 10.2 (24) |  |  |  | 15.4 (39) |  |  |  | 16.7 (38) |  |  |  |
|  |  | 1 | 48 (259) | 45.6 (212) | 44.8 (129) | 1.78E-02 ^a^ | 4.57E-03 ^a^ | 4.3E-03 ^a^ | 43.8 (103) | 5.48E-06 ^a^ | 1.06E-06 ^a^ | 4.2E-07 ^a^ | 49.6 (126) | 7.64E-01 | 6.41E-01 | 6.4E-01 | 41 (93) | 8.22E-03 ^a^ | 2.50E-03 ^a^ | 1.5E-03 ^a^ |
|  |  | 2 | 34.6 (187) | 31 (144) | 43.4 (125) |  |  |  | 46 (108) |  |  |  | 35 (89) |  |  |  | 42.3 (96) |  |  |  |
|  | **SLC22A18 (rs450208)** | 0 | 10.4 (56) | 10.3 (48) | 3.5 (10) |  |  |  | 6.8 (16) |  |  |  | 5.5 (14) |  |  |  | 7 (16) |  |  |  |
|  |  | 1 | 39.6 (214) | 43.7 (203) | 38.5 (111) | 1.21E-03 ^a^ | 1.50E-03 ^a^ | 1.4E-03 ^a^ | 40.7 (96) | 1.45E-01 ^b^ | 5.24E-02 ^a^ | 5.3E-02 ^a^ | 36.6 (93) | 3.06E-02 ^a^ | 1.07E-02 ^a^ | 9.1E-03 ^a^ | 37.4 (85) | 5.04E-02 ^a^ | 1.57E-02 ^a^ | 1.5E-02 ^a^ |
|  |  | 2 | 50.1 (271) | 46 (214) | 58 (167) |  |  |  | 52.5 (124) |  |  |  | 57.9 (147) |  |  |  | 55.5 (126) |  |  |  |
| **G2** | **IGF2BP2 (rs4402960)** | 0 | 66.4 (359) | 74.5 (344) | 63.3 (179) |  |  |  | 62.7 (148) |  |  |  | 65 (165) |  |  |  | 67.4 (153) |  |  |  |
|  |  | 1 | 30.5 (165) | 23.4 (108) | 32.2 (91) | 4.69E-01 | 2.64E-01 | 2.6E-01 | 32.6 (77) | 3.45E-03 ^a^ | 7.68E-04 ^a^ | 6.5E-04 ^a^ | 30.7 (78) | 6.88E-01 | 5.38E-01 | 5.4E-01 | 29.1 (66) | 1.30E-01 ^b^ | 4.35E-02 ^a^ | 4.1E-02 ^a^ |
|  |  | 2 | 3.1 (17) | 2.2 (10) | 4.6 (13) |  |  |  | 4.7 (11) |  |  |  | 4.3 (11) |  |  |  | 3.5 (8) |  |  |  |
|  | **TCF7L2 (rs7903146)** | 0 | 74.1 (401) | 81.8 (378) | 67.8 (192) |  |  |  | 64.8 (153) |  |  |  | 71.7 (182) |  |  |  | 67.8 (154) |  |  |  |
|  |  | 1 | 23.5 (127) | 16.9 (78) | 30.7 (87) | 5.85E-02 ^a^ | 1.51E-01 ^b^ | 1.5E-01 ^b^ | 31.8 (75) | 3.47E-06 ^a^ | 7.61E-07 ^a^ | 5.2E-07 ^a^ | 24.8 (63) | 5.82E-01 | 3.55E-01 | 3.4E-01 | 31.3 (71) | 8.86E-05 ^a^ | 2.25E-04 ^a^ | 2.7E-04 ^a^ |
|  |  | 2 | 2.4 (13) | 1.3 (6) | 1.4 (4) |  |  |  | 3.4 (8) |  |  |  | 3.5 (9) |  |  |  | 0.9 (2) |  |  |  |
|  | **CDKN2A (rs10811661)** | 0 | 1.8 (10) | 1.1 (5) | 0.7 (2) |  |  |  | 2.1 (5) |  |  |  | 1.2 (3) |  |  |  | 0.4 (1) |  |  |  |
|  |  | 1 | 15.9 (86) | 16.7 (77) | 12.4 (35) | 1.55E-01 ^b^ | 5.78E-02 ^a^ | 4.8E-02 ^a^ | 16.1 (38) | 5.47E-01 | 6.61E-01 | 6.5E-01 | 16.1 (41) | 7.86E-01 | 7.41E-01 | 7.3E-01 | 16.7 (38) | 6.95E-01 | 7.16E-01 | 7.2E-01 |
|  |  | 2 | 82.3 (445) | 82.3 (380) | 86.9 (246) |  |  |  | 81.8 (193) |  |  |  | 82.7 (210) |  |  |  | 82.8 (188) |  |  |  |
|  | **SLC30A8 (rs3802177)** | 0 | 54 (292) | 51.9 (240) | 48.4 (137) |  |  |  | 47 (111) |  |  |  | 52.4 (133) |  |  |  | 54.2 (123) |  |  |  |
|  |  | 1 | 37.5 (203) | 41.8 (193) | 37.5 (106) | 3.42E-02 ^a^ | 2.33E-02 ^a^ | 1.8E-02 ^a^ | 36.4 (86) | 8.69E-05 ^a^ | 4.13E-03 ^a^ | 3.4E-03 ^a^ | 42.9 (109) | 9.32E-02 ^a^ | 6.50E-01 | 6.5E-01 | 38.8 (88) | 7.33E-01 | 7.69E-01 | 7.7E-01 |
|  |  | 2 | 8.5 (46) | 6.3 (29) | 14.1 (40) |  |  |  | 16.5 (39) |  |  |  | 4.7 (12) |  |  |  | 7 (16) |  |  |  |
|  | **HNF1A (rs483353044)** | 0 | 99.4 (538) | 99.1 (458) | 98.9 (280) |  |  |  | 98.3 (232) |  |  |  | 98.4 (250) |  |  |  | 98.7 (224) |  |  |  |
|  |  | 1 | 0.6 (3) | 0.9 (4) | 1.1 (3) | 4.18E-01 | 4.18E-01 | 4.2E-01 | 1.7 (4) | 3.30E-01 | 3.31E-01 | 3.3E-01 | 1.6 (4) | 1.51E-01 ^b^ | 1.51E-01 ^b^ | 1.5E-01 ^b^ | 1.3 (3) | 5.75E-01 | 5.75E-01 | 5.8E-01 |
|  | **WFS1 (rs4458523)** | 0 | 6.3 (34) | 6.3 (29) | 4.9 (14) |  |  |  | 3.4 (8) |  |  |  | 7.1 (18) |  |  |  | 6.2 (14) |  |  |  |
|  |  | 1 | 34.4 (186) | 33.8 (156) | 34.3 (97) | 7.28E-01 | 5.31E-01 | 5.2E-01 | 30.9 (73) | 1.60E-01 ^b^ | 6.98E-02 ^a^ | 6.5E-02 ^a^ | 33.1 (84) | 8.76E-01 | 9.50E-01 | 9.5E-01 | 34.4 (78) | 9.88E-01 | 9.40E-01 | 9.4E-01 |
|  |  | 2 | 59.3 (321) | 60 (277) | 60.8 (172) |  |  |  | 65.7 (155) |  |  |  | 59.8 (152) |  |  |  | 59.5 (135) |  |  |  |
|  | **HMG20A (rs1005752)** | 0 | 34.4 (186) | 34.2 (159) | 34.8 (100) |  |  |  | 35.2 (83) |  |  |  | 36 (91) |  |  |  | 37.9 (86) |  |  |  |
|  |  | 1 | 46 (249) | 49.2 (229) | 46.3 (133) | 9.63E-01 | 8.13E-01 | 8.1E-01 | 45.3 (107) | 5.20E-01 | 7.27E-01 | 7.3E-01 | 49.8 (126) | 1.81E-01 ^b^ | 1.97E-01 ^b^ | 1.9E-01 ^b^ | 48 (109) | 5.43E-01 | 2.69E-01 | 2.7E-01 |
|  |  | 2 | 19.6 (106) | 16.6 (77) | 18.8 (54) |  |  |  | 19.5 (46) |  |  |  | 14.2 (36) |  |  |  | 14.1 (32) |  |  |  |
| **G3** | **SLC16A11 (rs75493593)** | 0 | 41.2 (223) | 40.5 (187) | 26.9 (76) |  |  |  | 35.2 (83) |  |  |  | 33.5 (85) |  |  |  | 33.5 (76) |  |  |  |
|  |  | 1 | 42.5 (230) | 44.6 (206) | 51.2 (145) | 2.12E-04 ^a^ | 1.40E-04 ^a^ | 8.8E-05 ^a^ | 41.1 (97) | 1.56E-02 ^a^ | 1.49E-02 ^a^ | 1.1E-02 ^a^ | 47.6 (121) | 1.10E-01 ^b^ | 5.64E-02 ^a^ | 4.8E-02 ^a^ | 52.4 (119) | 1.35E-01 ^b^ | 2.70E-01 | 2.7E-01 |
|  |  | 2 | 16.3 (88) | 14.9 (69) | 21.9 (62) |  |  |  | 23.7 (56) |  |  |  | 18.9 (48) |  |  |  | 14.1 (32) |  |  |  |
|  | **IRS1 (rs1801278)** | 0 | 94.5 (511) | 95.3 (443) | 96.5 (278) |  |  |  | 95.8 (226) |  |  |  | 92.9 (236) |  |  |  | 93.8 (213) |  |  |  |
|  |  | 1 | 5.2 (28) | 4.5 (21) | 3.5 (10) | 3.10E-01 | 1.46E-01 ^b^ | 1.3E-01 ^b^ | 3.8 (9) | 8.09E-01 | 8.76E-01 | 8.7E-01 | 7.1 (18) | 3.55E-01 | 5.43E-01 | 5.3E-01 | 5.7 (13) | 6.85E-01 | 3.93E-01 | 3.7E-01 |
|  |  | 2 | 0.4 (2) | 0.2 (1) | 0 (0) |  |  |  | 0.4 (1) |  |  |  | 0 (0) |  |  |  | 0.4 (1) |  |  |  |
|  | **FABP2 (rs1799883)** | 0 | 58.2 (315) | 58.5 (272) | 61.5 (177) |  |  |  | 52.5 (124) |  |  |  | 58.7 (149) |  |  |  | 59 (134) |  |  |  |
|  |  | 1 | 35.7 (193) | 34.6 (161) | 33.3 (96) | 6.43E-01 | 3.49E-01 | 3.4E-01 | 38.6 (91) | 2.87E-01 | 1.16E-01 ^b^ | 1.1E-01 ^b^ | 37.4 (95) | 4.40E-01 | 5.68E-01 | 5.7E-01 | 36.1 (82) | 5.70E-01 | 6.04E-01 | 6.0E-01 |
|  |  | 2 | 6.1 (33) | 6.9 (32) | 5.2 (15) |  |  |  | 8.9 (21) |  |  |  | 3.9 (10) |  |  |  | 4.8 (11) |  |  |  |
|  | **CAPN10 (rs7607759)** | 0 | 87.8 (474) | 91.2 (424) | 91.3 (263) |  |  |  | 90.7 (214) |  |  |  | 85.8 (218) |  |  |  | 89.4 (203) |  |  |  |
|  |  | 1 | 12.2 (66) | 8.8 (41) | 8 (23) | 2.80E-02 ^a^ | 8.25E-01 | 2.3E-01 | 9.3 (22) | 8.26E-01 | 6.68E-02 | 8.3E-01 | 14.2 (36) | 4.43E-01 | 4.44E-01 | 4.6E-01 | 10.1 (23) | 3.03E-01 | 2.26E-01 | 3.7E-01 |
|  |  | 2 | 0 (0) | 0 (0) | 0.7 (2) |  |  |  | 0 (0) |  |  |  | 0 (0) |  |  |  | 0.4 (1) |  |  |  |
|  | **PPP1R3A (rs1799999)** | 0 | 53.3 (289) | 47.7 (222) | 46 (132) |  |  |  | 51.7 (122) |  |  |  | 46.5 (118) |  |  |  | 48 (109) |  |  |  |
|  |  | 1 | 38.4 (208) | 42.6 (198) | 40.1 (115) | 1.89E-02 ^a^ | 7.89E-03 ^a^ | 6.0E-03 ^a^ | 42.4 (100) | 2.11E-01 | 1.33E-01 ^b^ | 1.4E-01 ^b^ | 42.9 (109) | 1.72E-01 ^b^ | 6.38E-02 ^a^ | 5.9E-02 ^a^ | 44.5 (101) | 6.23E-01 | 6.38E-01 | 6.4E-01 |
|  |  | 2 | 8.3 (45) | 9.7 (45) | 13.9 (40) |  |  |  | 5.9 (14) |  |  |  | 10.6 (27) |  |  |  | 7.5 (17) |  |  |  |
|  | **PTPRD (rs10511567)** | 0 | 57 (309) | 49.7 (231) | 45.5 (131) |  |  |  | 57.8 (137) |  |  |  | 50.8 (129) |  |  |  | 53.3 (121) |  |  |  |
|  |  | 1 | 33.9 (184) | 41.1 (191) | 46.9 (135) | 1.28E-03 ^a^ | 3.19E-02 ^a^ | 2.8E-02 ^a^ | 35.4 (84) | 1.10E-01 ^b^ | 3.88E-02 ^a^ | 3.6E-02 ^a^ | 40.6 (103) | 1.89E-01 ^b^ | 2.40E-01 | 2.2E-01 | 38.3 (87) | 6.66E-01 | 3.93E-01 | 3.9E-01 |
|  |  | 2 | 9 (49) | 9.2 (43) | 7.6 (22) |  |  |  | 6.8 (16) |  |  |  | 8.7 (22) |  |  |  | 8.4 (19) |  |  |  |
| **G4** | **IL6 (rs1800795)** | 0 | 84.1 (456) | 87.5 (407) | 86.1 (248) |  |  |  | 86.4 (204) |  |  |  | 86.6 (220) |  |  |  | 78.9 (179) |  |  |  |
|  |  | 1 | 14.2 (77) | 11.2 (52) | 13.5 (39) | 2.42E-01 | 2.61E-01 | 2.4E-01 | 13.1 (31) | 4.27E-01 | 9.41E-01 | 9.4E-01 | 12.2 (31) | 6.38E-01 | 3.43E-01 | 3.2E-01 | 20.3 (46) | 5.36E-03 | 1.10E-02 | 8.7E-03 ^a^ |
|  |  | 2 | 1.7 (9) | 1.3 (6) | 0.3 (1) |  |  |  | 0.4 (1) |  |  |  | 1.2 (3) |  |  |  | 0.9 (2) |  |  |  |
|  | **NOS3 (rs2070744)** | 0 | 73.1 (396) | 75.1 (349) | 80.6 (232) |  |  |  | 77.1 (182) |  |  |  | 76 (193) |  |  |  | 75.3 (171) |  |  |  |
|  |  | 1 | 23.4 (127) | 22.4 (104) | 16.3 (47) | 4.94E-02 ^a^ | 3.70E-02 ^a^ | 2.7E-02 ^a^ | 21.6 (51) | 5.01E-01 | 3.86E-01 | 3.8E-01 | 21.7 (55) | 5.60E-01 | 3.03E-01 | 2.8E-01 | 22 (50) | 9.94E-01 | 9.58E-01 | 9.6E-01 |
|  |  | 2 | 3.5 (19) | 2.6 (12) | 3.1 (9) |  |  |  | 1.3 (3) |  |  |  | 2.4 (6) |  |  |  | 2.6 (6) |  |  |  |
|  | **CPED1 (rs10261386)** | 0 | 41.4 (224) | 45.6 (212) | 41.7 (120) |  |  |  | 41.1 (97) |  |  |  | 45.3 (115) |  |  |  | 38.3 (87) |  |  |  |
|  |  | 1 | 45.7 (247) | 41.9 (195) | 47.6 (137) | 6.44E-01 | 6.19E-01 | 6.2E-01 | 46.6 (110) | 4.71E-01 | 4.30E-01 | 4.2E-01 | 40.6 (103) | 4.01E-01 | 6.15E-01 | 6.1E-01 | 47.1 (107) | 1.92E-01 ^b^ | 9.42E-02 ^a^ | 8.8E-02 ^a^ |
|  |  | 2 | 12.9 (70) | 12.5 (58) | 10.8 (31) |  |  |  | 12.3 (29) |  |  |  | 14.2 (36) |  |  |  | 14.5 (33) |  |  |  |
|  | **KHDRBS3 (rs6577691)** | 0 | 78.6 (425) | 76.3 (355) | 80.6 (232) |  |  |  | 82.6 (195) |  |  |  | 77.6 (197) |  |  |  | 82.4 (187) |  |  |  |
|  |  | 1 | 18.9 (102) | 21.9 (102) | 17 (49) | 7.94E-01 | 5.38E-01 | 5.1E-01 | 15.7 (37) | 1.44E-01 ^b^ | 8.72E-02 ^a^ | 8.2E-02 ^a^ | 20.5 (52) | 7.66E-01 | 9.17E-01 | 9.1E-01 | 17.2 (39) | 1.15E-01 ^b^ | 4.51E-02 ^a^ | 4.5E-02 ^a^ |
|  |  | 2 | 2.6 (14) | 1.7 (8) | 2.4 (7) |  |  |  | 1.7 (4) |  |  |  | 2 (5) |  |  |  | 0.4 (1) |  |  |  |
|  | **CACNA1H (rs4984636)** | 0 | 77.9 (422) | 79.6 (370) | 79.5 (229) |  |  |  | 80.6 (191) |  |  |  | 86.6 (220) |  |  |  | 79.7 (181) |  |  |  |
|  |  | 1 | 20.8 (113) | 19.1 (89) | 19.8 (57) | 6.75E-01 | 4.87E-01 | 4.9E-01 | 17.7 (42) | 8.34E-01 | 8.61E-01 | 8.6E-01 | 13.4 (34) | 6.28E-03 ^a^ | 1.80E-03 ^a^ | 1.9E-03 ^a^ | 19.4 (44) | 8.93E-01 | 8.71E-01 | 8.7E-01 |
|  |  | 2 | 1.3 (7) | 1.3 (6) | 0.7 (2) |  |  |  | 1.7 (4) |  |  |  | 0 (0) |  |  |  | 0.9 (2) |  |  |  |

Abbreviations: RA, risk allele; SNP, single nucleotide polymorphism; T2D, type 2 diabetes.

^a^P < 0.1 and ^b^P < 0.2 were selected for the multivariate model.

**Table S12.** Association of 23 SNP genotypes with T2D stratified by sex and age of T2D diagnosis (N = 2020)

| **Group** | **Gene (SNP)***  **(No. RA)** | **Univariate Logistic Regression Models** | | | | | | | | | | | |
| --- | --- | --- | --- | --- | --- | --- | --- | --- | --- | --- | --- | --- | --- |
|  |  | **T2D diagnosis ≤45 years** | | | | | | **T2D diagnosis ≥46 years** | | | | | |
|  |  | **Females** | | | **Males** | | | **Females** | | | **Males** | | |
|  |  | **OR (95% CI)** | **p-Wald** | **R^2^** | **OR (95% CI)** | **p-Wald** | **R^2^** | **OR (95% CI)** | **p-Wald** | **R^2^** | **OR (95% CI)** | **p-Wald** | **R^2^** |
| **G1** | **INS_IGF2 (1)** | 1.33 (0.8-2.3) | 3.2E-01 | 0.002 | 2.05 (1.1-4) ^c^ | 3.3E-02 ^a^ | 0.015 | 1.48 (0.8-2.7) | 2.0E-01 | 0.003 | 3.18 (1.4-7) ^c^ | 4.1E-03 ^a^ | 0.030 |
|  | **INS_IGF2 (2)** | 1.28 (0.7-2.2) | 3.8E-01 |  | 2.34 (1.2-4.5) | 9.3E-03 ^a^ |  | 1.43 (0.8-2.6) | 2.3E-01 |  | 3.81 (1.8-8.2) | 6.9E-04 ^a^ |  |
|  | **INS(1)** | 1.2 (0.9-1.6) | 2.5E-01 | 0.003 | 1.71 (1.2-2.4) ^c^ | 2.0E-03 ^a^ | 0.025 | 1.2 (0.9-1.7) | 2.8E-01 | 0.004 | 1.68 (1.2-2.4) ^c^ | 3.0E-03 ^a^ | 0.018 |
|  | **INS(2)** | 0.91 (0.5-1.8) | 8.0E-01 |  | 2.28 (1.1-4.6) | 2.4E-02 ^a^ |  | 1.51 (0.8-2.8) | 2.0E-01 |  | 1.27 (0.6-2.9) | 5.7E-01 |  |
|  | **KCNQ1(1)** | 1.25 (0.8-2) | 3.7E-01 | 0.009 | 2.24 (1.3-3.9) ^c^ | 4.1E-03 ^a^ | 0.059 | 1.1 (0.7-1.8) | 7.0E-01 | 0.001 | 1.07 (0.7-1.7) ^c^ | 7.6E-01 | 0.030 |
|  | **KCNQ1(2)** | 1.65 (1-2.7) | 4.2E-02 ^a^ |  | 3.9 (2.3-6.7) | 1.0E-06 ^a^ |  | 1.18 (0.7-1.9) | 4.9E-01 |  | 1.99 (1.3-3.1) | 3.5E-03 ^a^ |  |
|  | **CDKN1C (1)** | 1.38 (0.9-2.1) ^c^ | 1.6E-01 | 0.013 | 2.21 (1.3-3.6) ^c^ | 1.9E-03 ^a^ | 0.050 | 1.17 (0.8-1.8) | 4.7E-01 | 0.001 | 1.26 (0.8-2) ^c^ | 3.1E-01 | 0.019 |
|  | **CDKN1C (2)** | 1.85 (1.2-2.9) | 7.9E-03 ^a^ |  | 3.41 (2.1-5.7) | 2.2E-06 ^a^ |  | 1.15 (0.7-1.8) | 5.5E-01 |  | 1.91 (1.2-3) | 4.8E-03 ^a^ |  |
|  | **SLC22A18 (1)** | 2.9 (1.4-5.9) ^c^ | 3.3E-03 ^a^ | 0.025 | 1.42 (0.8-2.6) | 2.7E-01 | 0.008 | 1.74 (0.9-3.3) ^c^ | 8.8E-02 ^a^ | **0.013** | 1.26 (0.7-2.3) | 4.7E-01 | 0.012 |
|  | **SLC22A18 (2)** | 3.45 (1.7-6.9) | 5.2E-04 ^a^ |  | 1.74 (0.9-3.2) | 7.4E-02 ^a^ |  | 2.17 (1.2-4) | 1.4E-02 ^a^ |  | 1.77 (1-3.2) | 6.6E-02 ^a^ |  |
| **G2** | **IGF2BP2(1)** | 1.11 (0.8-1.5) | 5.3E-01 | 0.002 | 1.66 (1.2-2.4) ^c^ | 4.7E-03 ^a^ | 0.022 | 1.03 (0.7-1.4) | 8.7E-01 | 0.001 | 1.37 (1-2) | 8.4E-02 ^a^ | 0.008 |
|  | **IGF2BP2(2)** | 1.53 (0.7-3.2) | 2.6E-01 |  | 2.56 (1.1-6.2) | 3.6E-02 ^a^ |  | 1.41 (0.6-3.1) | 3.9E-01 |  | 1.8 (0.7-4.6) | 2.3E-01 |  |
|  | **TCF7L2(1)** | 1.43 (1-2) ^c^ | 2.9E-02 ^a^ | 0.009 | 2.38 (1.6-3.4) ^c^ | 4.2E-06 ^a^ | 0.047 | 1.09 (0.8-1.5) | 6.2E-01 | 0.002 | 2.23 (1.5-3.2) ^c^ | 2.3E-05 ^a^ | 0.036 |
|  | **TCF7L2(2)** | 0.64 (0.2-2) | 4.4E-01 |  | 3.29 (1.1-9.7) | 3.0E-02 ^a^ |  | 1.53 (0.6-3.6) | 3.4E-01 |  | 0.82 (0.2-4.1) | 8.1E-01 |  |
|  | **CDKN2A(1)** | 2.03 (0.4-9.8) ^c^ | 3.7E-01 | 0.007 | 0.49 (0.1-1.8) | 2.9E-01 | 0.001 | 1.59 (0.4-6.1) | 5.0E-01 | 0.001 | 2.47 (0.3-21.9) | 4.2E-01 | 0.002 |
|  | **CDKN2A(2)** | 2.76 (0.6-12.7) | 1.9E-01 ^b^ |  | 0.51 (0.1-1.8) | 2.9E-01 |  | 1.57 (0.4-5.8) | 4.9E-01 |  | 2.47 (0.3-21.3) | 4.1E-01 |  |
|  | **SLC30A8 (1)** | 1.11 (0.8-1.5) ^c^ | 5.0E-01 | 0.011 | 0.96 (0.7-1.4) ^c^ | 8.3E-01 | 0.035 | 1.18 (0.9-1.6) ^c^ | 3.0E-01 | 0.009 | 0.89 (0.6-1.2) | 4.9E-01 | 0.001 |
|  | **SLC30A8 (2)** | 1.85 (1.2-3) | 1.0E-02 ^a^ |  | 2.91 (1.7-4.9) | 8.1E-05 ^a^ |  | 0.57 (0.3-1.1) | 1.0E-01 ^a^ |  | 1.08 (0.6-2.1) | 8.2E-01 |  |
|  | **HNF1A(1)** | 1.92 (0.4-9.6) | 4.3E-01 | 0.001 | 1.97 (0.5-8) | 3.4E-01 | 0.002 | 2.87 (0.6-12.9) ^c^ | 1.7E-01 ^b^ | 0.003 | 1.53 (0.3-6.9) | 5.8E-01 | 0.001 |
|  | **WFS1(1)** | 1.27 (0.6-2.5) | 4.9E-01 | 0.001 | 1.7 (0.7-3.9) ^c^ | 2.1E-01 | 0.008 | 0.85 (0.5-1.6) | 6.2E-01 | 0.000 | 1.04 (0.5-2.1) | 9.2E-01 | 0.000 |
|  | **WFS1(2)** | 1.3 (0.7-2.5) | 4.3E-01 |  | 2.03 (0.9-4.5) | 8.6E-02 ^a^ |  | 0.89 (0.5-1.6) | 7.2E-01 |  | 1.01 (0.5-2) | 9.8E-01 |  |
|  | **HMG20A(1)** | 0.99 (0.7-1.4) | 9.7E-01 | 0.000 | 0.9 (0.6-1.3) | 5.4E-01 | 0.003 | 1.03 (0.7-1.4) | 8.4E-01 | 0.006 | 0.88 (0.6-1.2) | 4.7E-01 | 0.002 |
|  | **HMG20A(2)** | 0.95 (0.6-1.4) | 8.0E-01 |  | 1.14 (0.7-1.8) | 5.6E-01 |  | 0.69 (0.4-1.1) ^c^ | 1.1E-01 ^b^ |  | 0.77 (0.5-1.3) | 2.9E-01 |  |
| **G3** | **SLC16A11(1)** | 1.85 (1.3-2.6) ^c^ | 3.0E-04 ^a^ | 0.029 | 1.06 (0.7-1.5) ^c^ | 7.4E-01 | 0.016 | 1.38 (1-1.9) ^c^ | 5.8E-02 ^a^ | 0.008 | 1.42 (1-2) ^c^ | 4.9E-02 ^a^ | 0.008 |
|  | **SLC16A11(2)** | 2.07 (1.4-3.1) | 6.3E-04 ^a^ |  | 1.83 (1.2-2.8) | 6.8E-03 ^a^ |  | 1.43 (0.9-2.2) | 1.0E-01 |  | 1.14 (0.7-1.9) | 6.0E-01 |  |
|  | **IRS1(1)** | 0.66 (0.3-1.4) | 2.6E-01 | 0.005 | 0.84 (0.4-1.9) | 6.7E-01 | 0.047 | 1.39 (0.8-2.6) ^c^ | 2.9E-01 | 0.005 | 1.29 (0.6-2.6) | 4.9E-01 | 0.001 |
|  | **IRS1(2)** |  |  |  | 1.96 (0.1-31.5) | 6.3E-01 |  |  |  |  | 2.08 (0.1-33.4) | 6.1E-01 |  |
|  | **FABP2(1)** | 0.89 (0.7-1.2) | 4.4E-01 | 0.001 | 1.24 (0.9-1.7) ^c^ | 2.1E-01 ^b^ | 0.005 | 1.04 (0.8-1.4) | 8.0E-01 | 0.003 | 1.03 (0.7-1.4) | 8.5E-01 | 0.002 |
|  | **FABP2(2)** | 0.81 (0.4-1.5) | 5.1E-01 |  | 1.44 (0.8-2.6) | 2.3E-01 |  | 0.64 (0.3-1.3) | 2.3E-01 |  | 0.7 (0.3-1.4) | 3.2E-01 |  |
|  | **CAPN10(1)** | 0.63 (0.4-1) ^c^ | 6.7E-02 ^a^ | 0.013 | 1.06 (0.6-1.8) | 8.3E-01 | 0.000 | 1.19 (0.8-1.8) | 4.4E-01 | 0.001 | 1.17 (0.7-2) | 5.6E-01 | 0.005 |
|  | **PPP1R3A(1)** | 1.21 (0.9-1.6) ^c^ | 2.2E-01 | 0.013 | 0.92 (0.7-1.3) | 6.1E-01 | 0.006 | 1.28 (0.9-1.8) | 1.2E-01 ^b^ | 0.006 | 1.04 (0.7-1.4) | 8.2E-01 | 0.002 |
|  | **PPP1R3A(2)** | 1.95 (1.2-3.1) | 5.8E-03 ^a^ |  | 0.57 (0.3-1.1) | 8.1E-02 ^a^ |  | 1.47 (0.9-2.5) | 1.5E-01 ^b^ |  | 0.77 (0.4-1.4) | 3.9E-01 |  |
|  | **PTPRD(1)** | 1.73 (1.3-2.3) ^c^ | 3.7E-04 ^a^ | 0.022 | 0.74 (0.5-1) | 7.8E-02 ^a^ | 0.009 | 1.34 (1-1.8) ^c^ | 7.0E-02 ^a^ | 0.006 | 0.87 (0.6-1.2) | 4.1E-01 | 0.002 |
|  | **PTPRD(2)** | 1.06 (0.6-1.8) | 8.4E-01 |  | 0.63 (0.3-1.2) | 1.4E-01 |  | 1.08 (0.6-1.9) | 7.9E-01 |  | 0.84 (0.5-1.5) | 5.7E-01 |  |
| **G4** | **IL6(1)** | 0.93 (0.6-1.4) | 7.4E-01 | 0.006 | 1.19 (0.7-1.9) | 4.7E-01 | 0.004 | 0.83 (0.5-1.3) | 4.3E-01 | 0.002 | 2.01 (1.3-3.1) ^c^ | 1.6E-03 ^a^ | 0.020 |
|  | **IL6(2)** | 0.2 (0-1.6) | 1.3E-01 ^b^ |  | 0.33 (0-2.8) | 3.1E-01 |  | 0.69 (0.2-2.6) | 5.8E-01 |  | 0.76 (0.2-3.8) | 7.4E-01 |  |
|  | **NOS3(1)** | 0.63 (0.4-0.9) ^c^ | 1.5E-02 ^a^ | 0.010 | 0.94 (0.6-1.4) | 7.5E-01 | 0.003 | 0.89 (0.6-1.3) | 5.2E-01 | 0.002 | 0.98 (0.7-1.4) | 9.2E-01 | 0.000 |
|  | **NOS3(2)** | 0.81 (0.4-1.8) | 6.1E-01 |  | 0.48 (0.1-1.7) | 2.6E-01 |  | 0.65 (0.3-1.6) | 3.6E-01 |  | 1.02 (0.4-2.8) | 9.7E-01 |  |
|  | **CPED1(1)** | 1.04 (0.8-1.4) | 8.2E-01 | 0.001 | 1.23 (0.9-1.7) | 2.2E-01 | 0.003 | 0.81 (0.6-1.1) | 2.0E-01 | 0.003 | 1.34 (0.9-1.9) | 9.7E-02 ^a^ | 0.007 |
|  | **CPED1(2)** | 0.83 (0.5-1.3) | 4.3E-01 |  | 1.09 (0.7-1.8) | 7.3E-01 |  | 1 (0.6-1.6) | 9.9E-01 |  | 1.39 (0.8-2.3) | 2.0E-01 |  |
|  | **KHDRBS3(1)** | 0.88 (0.6-1.3) | 5.1E-01 | 0.001 | 0.66 (0.4-1) ^c^ | 5.0E-02 ^a^ | 0.008 | 1.1 (0.8-1.6) | 6.2E-01 | 0.001 | 0.73 (0.5-1.1) ^c^ | 1.2E-01 ^b^ | 0.010 |
|  | **KHDRBS3(2)** | 0.92 (0.4-2.3) | 8.5E-01 |  | 0.91 (0.3-3.1) | 8.8E-01 |  | 0.77 (0.3-2.2) | 6.2E-01 |  | 0.24 (0-1.9) | 1.8E-01 ^b^ |  |
|  | **CACNA1H(1)** | 0.93 (0.7-1.3) | 6.9E-01 | 0.001 | 0.91 (0.6-1.4) | 6.7E-01 | 0.001 | 0.58 (0.4-0.9) ^c^ | 9.7E-03 ^a^ | 0.022 | 1.01 (0.7-1.5) | 9.6E-01 | 0.000 |
|  | **CACNA1H(2)** | 0.53 (0.1-2.6) | 4.3E-01 |  | 1.29 (0.4-4.6) | 6.9E-01 |  |  |  |  | 0.68 (0.1-3.4) | 6.4E-01 |  |
| **No. SNPs** | | 10 | | | 13 | | | 5 | | | 10 | | |
| **ΣR^2^** | | 0.155 | | | 0.306 | | | 0.057 | | | 0.226 | | |

Abbreviations: SNP, single nucleotide polymorphism; T2D, type 2 diabetes.

^a^P < 0.1; ^b^P < 0.2; ^c^Remained in the multivariate model

**Table S13.** Analysis of linkage disequilibrium of five SNPs located in Chr 11p15.5 region (insulin and KCNQ1 genes region) in the control group (n = 1008)

| **SNP** | **Alleles** | **Frequency** | | | | **Disequilibrium Linkage** | | |
| --- | --- | --- | --- | --- | --- | --- | --- | --- |
|  |  | **Observed/Expected** | | **Observed C+T** | ***P* value** | **D'** | | **r^2^** |
| **INS-IGF2 (rs149483638) vs.** | | **C** | **T** |  |  | **C** | **T** |  |
| **INS (rs689)** | T | 1134/1140 | 267/260 | 1401 | >0.05 | -0.0608 | 0.0608 | 0.0003 |
|  | A | 458/451 | 97/103 | 555 | >0.05 | 0.0608 | -0.0608 | 0.0003 |
| **KCNQ1 (rs2237897)** | T | 603/551 | 798/850 | 1402 | <0.00001 | 0.2392 | -0.0267 | 0.0147 |
|  | C | 166/218 | 389/337 | 555 | <0.00001 | -0.0267 | 0.2392 | 0.0147 |
| **CDKN1C (rs163168)** | C | 885/784 | 516/617 | 1401 | <0.00001 | 0.3267 | -0.3267 | 0.0537 |
|  | T | 209/310 | 346/245 | 555 | <0.00001 | -0.3267 | 0.3267 | 0.0537 |
| **SLC22A18 (rs450208)** | T | 1063/962 | 338/439 | 1401 | <0.00001 | 0.2652 | -0.2652 | 0.061 |
|  | G | 280/381 | 275/174 | 555 | <0.00001 | -0.2652 | 0.2652 | 0.061 |
| **INS (rs689) vs.** | | **A** | **T** |  |  | **A** | **T** |  |
| **KCNQ1 (rs2237897)** | T | 690/626 | 902/966 | 1592 | <0.00001 | 0.448 | -0.448 | 0.0297 |
|  | C | 79/143 | 285/221 | 364 | <0.00001 | -0.448 | -0.448 | 0.0297 |
| **CDKN1C (rs163168)** | C | 957/890 | 635/701 | 1592 | <0.00001 | 0.3271 | -0.3271 | 0.031 |
|  | T | 137/204 | 227/160 | 364 | <0.00001 | -0.3271 | 0.3271 | 0.031 |
| **SLC22A18 (rs450208)** | T | 1144/1093 | 448/499 | 1592 | <0.00001 | 0.2038 | -0.2038 | 0.0208 |
|  | G | 199/250 | 165/114 | 364 | <0.00001 | -0.2038 | 0.2038 | 0.0208 |
| **KCNQ1 (rs2237897) vs.** | | **C** | **T** |  |  | **C** | **T** |  |
| **CDKN1C (rs163168)** | C | 410/430 | 359/339 | 769 | >0.05 | -0.0467 | 0.0467 | 0.0018 |
|  | T | 684/664 | 503/523 | 1187 | >0.05 | 0.0467 | -0.0467 | 0.0018 |
| **SLC22A18 (rs450208)** | T | 563/528 | 206/241 | 769 | <0.001 | 0.1452 | -0.1452 | 0.0062 |
|  | G | 780/815 | 407/372 | 1187 | <0.001 | -0.1452 | 0.1452 | 0.0062 |
| **CDKN1C (rs163168) vs.** | | **C** | **T** |  |  | **C** | **T** |  |
| **SLC22A18 (rs450208)** | T | 954/751 | 140/343 | 1094 | <0.00001 | 0.5917 | -0.5917 | 0.2028 |
|  | G | 389/592 | 473/270 | 862 | <0.00001 | -0.5917 | 0.5917 | 0.2028 |

Abbreviations: D', Tajima statistic; r2, statistic for linkage disequilibrium; SNP, single nucleotide polymorphism; T2D, type 2 diabetes.

P value calculated with chi square.

**Table S14.** Genotype multivariate logistic regression models with 23 SNPs stratified by sex and age of T2D diagnosis (N = 2020)

| **Group** | **SNPs** | **RA** | **OR (95% CI)** | **p-Wald** |
| --- | --- | --- | --- | --- |
| **Females T2D diagnosis ≤45 years** | | | **controls, n = 416/cases, n = 241** | |
| G1 | KCNQ1 (rs163168) | 1 | 1.15 (0.7-1.9) | 5.7E-01 |
|  |  | 2 | 1.56 (0.9-2.6) | 8.3E-02 |
|  | SLC22A18 (rs450208) | 1 | 2.94 (1.3-6.4) | 6.7E-03 |
|  |  | 2 | 3.43 (1.6-7.5) | 1.8E-03 |
| G2 | TCF7L2 (rs7903146) | 1 | 1.6 (1.1-2.3) | 8.8E-03 |
|  |  | 2 | 0.68 (0.2-2.2) | 5.1E-01 |
|  | CDKN2A (rs10811661) | 1 | 2.28 (0.4-12) | 3.3E-01 |
|  |  | 2 | 2.78 (0.6-14) | 2.1E-01 |
|  | SLC30A8 (rs3802177) | 1 | 0.91 (0.6-1.3) | 5.7E-01 |
|  |  | 2 | 1.48 (0.9-2.5) | 1.4E-01 |
| G3 | SLC16A11 (rs75493593) | 1 | 2.02 (1.4-2.9) | 1.3E-04 |
|  |  | 2 | 1.91 (1.2-3) | 4.8E-03 |
|  | CAPN10 (rs7607759) | 1 | 0.53 (0.3-0.9) | 2.4E-02 |
|  | PPP1R3A (rs1799999) | 1 | 1.05 (0.8-1.5) | 7.7E-01 |
|  |  | 2 | 1.91 (1.1-3.2) | 1.4E-02 |
|  | PTPRD (rs10511567) | 1 | 1.73 (1.2-2.4) | 1.1E-03 |
|  |  | 2 | 0.85 (0.5-1.6) | 6.1E-01 |
|  | NOS3 (rs2070744) | 1 | 0.67 (0.4-1) | 5.8E-02 |
|  |  | 2 | 0.98 (0.4-2.4) | 9.6E-01 |
| **Males T2D diagnosis ≤45 years** | | | **controls, n = 356/cases, n = 209** | |
| G1 | INS-IGF2 (rs149483638)/INS (rs689) | 1 | 2.65 (1.2-5.9) | 1.6E-02 |
|  |  | 2 | 2.51 (1.2-5.4) | 1.8E-02 |
|  |  | 3 | 3.6 (1.6-8.1) | 1.9E-03 |
|  |  | 4 | 3.69 (1.3-10.5) | 1.4E-02 |
|  | KCNQ1 (rs2237897/rs163168) | 1 | 2.53 (0.9-7.1) | 7.8E-02 |
|  |  | 2 | 3.7 (1.9-7.3) | 1.5E-04 |
|  |  | 3 | 8.32 (3.6-19.1) | 5.9E-07 |
|  |  | 4 | 5.83 (2.9-11.5) | 4.1E-07 |
| G2 | IGF2BP2 (rs4402960) | 1 | 1.59 (1.1-2.4) | 2.2E-02 |
|  |  | 2 | 2.11 (0.8-5.9) | 1.5E-01 |
|  | TCF7L2 (rs7903146) | 1 | 2.22 (1.5-3.4) | 2.4E-04 |
|  |  | 2 | 3.37 (1-11.9) | 5.8E-02 |
|  | SLC30A8 (rs3802177) | 1 | 0.89 (0.6-1.3) | 5.5E-01 |
|  |  | 2 | 3.38 (1.8-6.2) | 7.8E-05 |
|  | WFS1 (rs4458523) | 1 | 2.33 (0.9-5.9) | 7.3E-02 |
|  |  | 2 | 3.13 (1.3-7.7) | 1.3E-02 |
| G3 | SLC16A11 (rs75493593) | 1 | 1.13 (0.8-1.7) | 5.7E-01 |
|  |  | 2 | 3.01 (1.8-5) | 2.1E-05 |
|  | FABP2 (rs1799883) | 1 | 1.42 (1-2.1) | 7.2E-02 |
|  |  | 2 | 1.52 (0.8-3) | 2.2E-01 |
| G4 | KHDRBS3 (rs6577691) | 1 | 0.48 (0.3-0.8) | 3.1E-03 |
|  |  | 2 | 0.89 (0.2-3.3) | 8.7E-01 |
| **Females T2D diagnosis ≥46 years** | | | **controls, n = 416/cases, n = 216** | |
| G1 | SLC22A18 (rs450208) | 1 | 1.93 (1-3.7) | 4.9E-02 |
|  |  | 2 | 2.55 (1.3-4.8) | 4.2E-03 |
| G2 | SLC30A8 (rs3802177) | 1 | 1.14 (0.8-1.6) | 4.3E-01 |
|  |  | 2 | 0.59 (0.3-1.2) | 1.3E-01 |
|  | HNF1A (rs483353044) | 1 | 3.29 (0.7-15.6) | 1.3E-01 |
|  | HMG20A (rs1005752) | 1 | 0.96 (0.7-1.4) | 8.3E-01 |
|  |  | 2 | 0.66 (0.4-1.1) | 7.9E-02 |
| G3 | SLC16A11 (rs75493593) | 1 | 1.4 (1-2) | 5.8E-02 |
|  |  | 2 | 1.25 (0.8-1.9) | 3.3E-01 |
|  | IRS1 (rs1801278) | 1 | 1.4 (0.7-2.6) | 3.0E-01 |
|  |  | 2 | 0 (0-0) | 1.0E+00 |
|  | PTPRD (rs10511567) | 1 | 1.44 (1-2) | 3.0E-02 |
|  |  | 2 | 1.16 (0.7-2) | 6.0E-01 |
| G4 | CACNA1H (rs4984636) | 1 | 0.58 (0.4-0.9) | 1.3E-02 |
|  |  | 2 | 0 (0-0) | 1.0E+00 |
| **Males T2D diagnosis ≥46 years** | | | **controls, n = 356/cases, n = 200** | |
| G1 | INS-IGF2 (rs149483638)/INS (rs689) | 1 | 2.64 (1.1-6.1) | 2.3E-02 |
|  |  | 2 | 3.09 (1.4-6.9) | 5.9E-03 |
|  |  | 3 | 4.66 (2-10.8) | 3.2E-04 |
|  |  | 4 | 2.15 (0.7-7.1) | 2.1E-01 |
|  | KCNQ1 (rs2237897/rs163168) | 1 | 0.9 (0.3-2.4) | 8.4E-01 |
|  |  | 2 | 1.1 (0.7-1.8) | 7.1E-01 |
|  |  | 3 | 1.67 (0.8-3.4) | 1.5E-01 |
|  |  | 4 | 1.75 (1-2.9) | 3.3E-02 |
| G2 | TCF7L2 (rs7903146) | 1 | 2 (1.3-3) | 7.3E-04 |
|  |  | 2 | 0.91 (0.2-4.7) | 9.1E-01 |
| G3 | SLC16A11 (rs75493593) | 1 | 1.54 (1.1-2.2) | 2.1E-02 |
|  |  | 2 | 1.37 (0.8-2.3) | 2.4E-01 |
| G4 | IL6 (rs1800795) | 1 | 1.62 (1-2.6) | 4.4E-02 |
|  |  | 2 | 1.24 (0.2-6.6) | 8.0E-01 |
|  | KHDRBS3 (rs6577691) | 1 | 0.66 (0.4-1) | 6.1E-02 |
|  |  | 2 | 0.2 (0-1.7) | 1.4E-01 |

G1: Insulin production (Chr 11p15.5 region); G2: Insulin production (Other genomic regions); G3: Insulin resistance; G4: Inflammation and other functions

Abbreviations: CI, confidence interval; OR, odds ratio; RA, risk allele; SNP, single nucleotide polymorphism; T2D, type 2 diabetes.
